# Cost-effectiveness of human papillomavirus self-sampling in the Swedish cervical screening program

**DOI:** 10.1101/2025.02.12.25322120

**Authors:** Ellinor Östensson, Christer Borgfeldt, Kine Pedersen, Kristina Hellman, Stephen Sy, Jiayao Lei, Emily A. Burger, Mark S. Clements

**Author notes:** Corresponding author: Mark Clements. Equal contributions (Christer Borgfeldt, Kine Pedersen). Ministry of Social Affairs (Socialdepartmentet), Stockholm, Sweden.

## Abstract

**Background:** In Sweden, the cervical cancer screening programme is based on primary human papillomavirus (HPV) testing with either clinician-collected cervical sampling or home-based vaginal self-sampling. We assessed the effectiveness and cost-effectiveness of primary HPV clinician-collected sampling and primary HPV self-collected sampling for unvaccinated cohorts of Swedish women.

**Methods:** A model-based analysis was performed to project long-term costs and quality-adjusted life-years (QALYs). Screening strategies included no screening, 18 clinician-collected strategies and 36 self-sampling strategies, with variations in the screening frequency, start age and follow-up management. We estimated incremental cost-effectiveness ratios benchmarked against willingness-to-pay (WTP) thresholds of €50,000 and €100,000 per QALY gained.

**Results:** Compared with the 2022 recommendations (with primary clinician-collected HPV testing from ages 23 with 5-yearly screening to age 50 and 7-yearly screening through to age 64), self-sampling at the same intensity would lead to similar effectiveness and a 36% reduction in costs. Among the clinician-collected sampling strategies, the optimal strategies involved primary HPV testing from age 25 with 10-yearly screening with extended genotyping (at €50,000 per QALY gained), or 7- and 10-yearly screening for €100,000. Across all strategies, the optimal strategies included primary self-sampling from age 25 with direct referral to colposcopy for HPV-16/18/45 with 7- and 10-yearly screening at €50,000 per QALY gained, and 5- and 7-yearly screening for €100,000 per QALY gained. These results were sensitive to the assumed accuracy of self-sampling compared with clinician-collected sampling.

**Conclusions:** Transitioning from clinician-collected cervical sampling to vaginal HPV self-sampling is likely to be cost-effective for unvaccinated women in Sweden.

**Highlights:**

- For an analysis restricted to clinician-collected sampling, the 2022 Swedish population-based recommendations (primary HPV testing from age 23 with 5-yearly screening to age 50 and 7-yearly screening between ages 50 and 64 years) were on the cost-efficiency frontier, however longer screening intervals were optimal.
- For all of the considered strategies, the optimal strategies were primary vaginal self-sampling from age 25 years with direct referral to colposcopy for HPV 16, 18 and 45 with seven- and ten-yearly screening at €50,000 per QALY gained, and with five- and seven-yearly screening at €100,000 per QALY gained.

## Introduction

To increase the effectiveness of organised cervical cancer screening programs, primary human papillomavirus (HPV) testing has been introduced in several countries due to its better performance compared with cytology [1, 2]. In Sweden, cervical cancer screening recommendations from 2015 included primary screening using cytology below age 30 years followed by HPV testing for abnormal cytology, and primary HPV testing from age 30 years. Swedish recommendations from 2022 included primary HPV testing with either clinician-collected endocervical sampling or vaginal self-collection (i.e., self-sampling) between ages 23 and 70 years with 5-yearly screening before age 50 years and 7-yearly screening between ages 50 and 64 years. Vaginal self-sampling HPV analyses have shown similar sensitivity to detect high grade intraepithelial cervical lesions (HSIL, which includes cancer, cervical intraepithelial neoplasia (CIN) 3 and most CIN 2) in the organised screening setting as clinician-collected cervical HPV analyses [3, 4].

The risk of disease progression from HPV infection to lesions depends on the HPV genotype. Thirteen HPV genotypes have been associated with the highest risk of cervical cancer, of which HPV 16, 18 and 45 cause the majority of cases [5-7]. In contemporary screening programs, HPV diagnostics may use a *partial* genotyping result for HPV 16 and 18 and/or 45, with the remaining 11 high-risk HPV genotypes given as a pooled result. *Extended genotyping* requires assays that report a minimum of 6 individual genotypes, and *full genotyping* requires the assay to report all high-risk genotypes individually [8]. In certain regions (i.e. Stockholm Region), extended genotyping has been implemented since 2022. A recent systematic review showed that extended HPV genotyping to the other 11 genotypes is a useful risk stratification method in cervical cancer screening [9].

In addition to offering self-sampling as an alternative to clinician-collected screening, home-based self-sampling can improve screening compliance [10]. In addition to the similar clinical accuracy, previous studies offering self-sampling as a choice showed a high acceptance rate and higher participation rates among under-screened women [3, 4, 11-13]. During the COVID-19 pandemic, HPV self-sampling was used in five regions in Sweden as the primary screening method. Post-pandemic, many health regions in Sweden continued with primary self-sampling which supported the formal introduction of self-sampling in the 2022 guidelines. However, no Swedish studies have evaluated the clinical effectiveness with self-sampling algorithms including extended genotyping, or offered self-sampling to all women within the screening age; moreover, few studies have projected the cost-effectiveness of self-sampling in primary screening within population-based screening [14-16]. Although the 2022 recommendations included partial genotyping and the option of self-sampling, no formal cost-effectiveness analysis in Sweden was performed despite cost-effectiveness being a key decision criterion for priority-setting in Sweden. As Swedish guidelines have not been adapted to reflect the lower cervical cancer risk for HPV vaccinated women, our analysis focuses on the large group of unvaccinated women. Consequently, we aim to evaluate the cost-effectiveness of screening with clinician-collected cervical sampling or vaginal self-sampling within the Swedish organised screening program for unvaccinated women.

## Material and methods

### Analytic overview

We conducted a model-based cost-effectiveness analysis to evaluate the lifetime costs and quality-adjusted life-years (QALYs) for alternative cervical cancer screening strategies in unvaccinated women. We used a well-validated individual-based simulation model [17] to compare cervical screening strategies that included both primary clinician-collected sampling and primary self-sampling. We evaluated outcomes of a cohort of 3 million females from age 9 years over their lifetime. Model outcomes included the discounted lifetime costs per woman, quality-adjusted life expectancy (QALE) per woman, the lifetime risk of detected cervical cancer per 100 000 women, the expected number of colposcopies over a lifetime, and the proportion of cancers detected at a local stage.

### Screening strategies and assumptions

We compared a total of 55 cervical screening strategies. The candidate strategies included the 2015 and 2022 Swedish cervical cancer screening recommendations together with strategies that were selected based on discussions with key experts (Table 1). In addition to a no screening strategy, the simulated screening strategies varied by: primary screening test using either clinician-collected HPV samples (including strategies that involved switching from primary cytology to primary HPV testing at age 30 years) or primary HPV self-sampling (including strategies that involved extended genotyping or direct referral to colposcopy following a positive test indicating HPV 16, 18 or 45); age to start screening (ages 23 or 25 years); and age-stratified screening intervals split before and from age 50 years (with combinations of 3-7y, 5-7y, 7-7y, 5-10y, 7-10y and 10-10y) (Figure 1 and Appendix D). For the base case analysis, we assumed full compliance with screening, follow-up, colposcopy referrals and treatment for precancerous lesions to prevent biasing findings towards over-screening [18]. We further assumed equal diagnostic accuracy of the HPV analysis for the clinician-collected sampling and self-sampling.

**Table 1.** Description of cervical clinician-collected (CC) and vaginal self-sampling (SS) screening strategies.

| Label | Description | Start age (years) | Screening intervals before and from age 50 years | Number of lifetime screens | Stop age (years) |
| --- | --- | --- | --- | --- | --- |
| CC-HPV(s) | Cytology before age 30 and switch (s) to HPV testing from age 30 y with analysis for types 16/18 | 23, 25 | 3-7y, 5-7y, 5-10y | 12, 11, 8 | 70 |
| CC-HPV | HPV testing with analysis for types 16/18/45 | 23, 25 | 3-7y, 5-7y, 7-7y, 5-10y, 7-10y, 10-10y | 12, 11, 8, 6, 5 | 70 |
| SS-HPV | Self-sampling with HPV analysis for types 16/18 | 23, 25 | 3-7y, 5-7y, 7-7y, 5-10y, 7-10y, 10-10y | 12, 11, 8, 6, 5 | 70 |
| SS-HPV(r) | Self-sampling with HPV analysis for type 16/18/45 with direct referral to colposcopy at positive result. Repeat self-sampling at positive test result for other types. | 23, 25 | 3-7y, 5-7y, 7-7y, 5-10y, 7-10y, 10-10y | 12, 11, 8, 6, 5 | 70 |
| SS-HPV EG | Self-sampling with HPV analysis for extended genotypes of 16/18/45/31/33/52/58. At age 64 years, HPV tests for additional types of 35/39/51/56/59/66/68 are included. | 23, 25 | 3-7y, 5-7y, 7-7y, 5-10y, 7-10, 10-10y | 12, 11, 8, 6, 5 | 70 |

**Figure 1.**
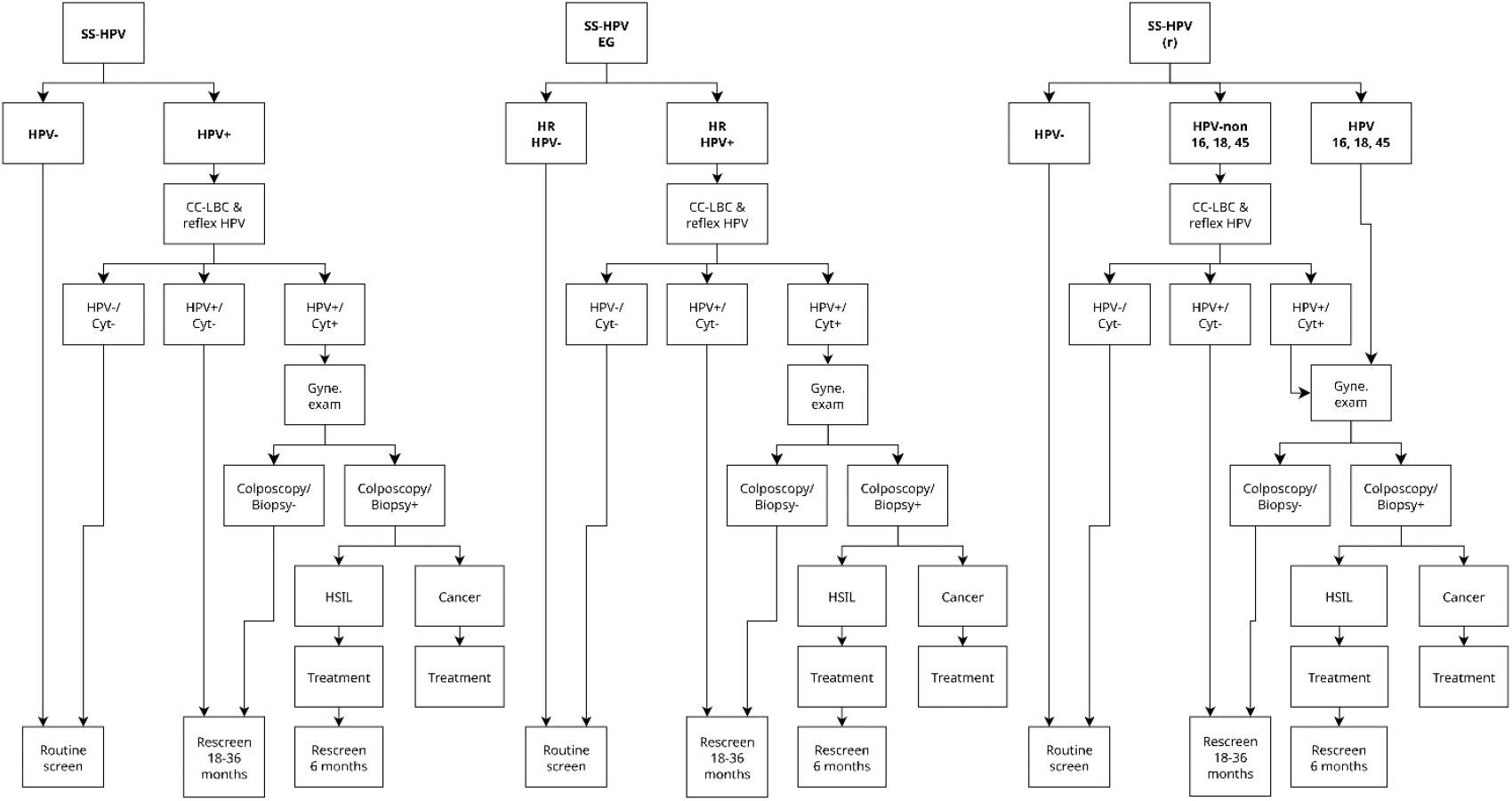
Candidate self-sampling screening algorithms. Note: Details for clinician-collected testing and follow-up management given abnormal test results are presented in Appendix D. Note that the SS-HPV and SS-HPV EG strategies only vary by the HPV test. Abbreviations: HPV=Human papillomavirus; SS=Self-sampling; EG=Extended genotyping; (r)=Direct referral to colposcopy for HPV 16/18/45; Cyt=Cytology; HSIL=High-grade squamous intraepithelial lesions; CC=Clinician-collected; LBC=Liquid-based cytology.

### Simulation model

The simulation model [17] had previously been adjusted to reflect HPV and cervical cancer epidemiology in Sweden [15]. At monthly intervals, women transition between health states, including; healthy; HPV infection (by individual HPV types 16, 18, 31, 33, 45, 52, 58, pooled other high-risk HPV types and pooled low-risk types); cervical precancerous lesions CIN2 or CIN3 by HPV type; squamous cell carcinoma by stage (local, regional, and distant cervical cancer); and death. The model does not include rare cervical cancer types. To align the natural history model (based on CIN) with Swedish guidelines (based on HSIL), most CIN2-3 are classified as HSIL in Sweden. A likelihood-based calibration approach was used to identify 50 good-fitting parameter sets. There were 55 strategies and 50 parameter sets, of which 13 (0.5%) simulations were missing due to output failures. We used 100 multiple imputations to impute the outcomes associated with the missing strategy-parameter set combination (see Appendix E). All model outcomes were reported as means across the imputations and across the parameter sets [15].

### Cost-effectiveness analysis

Following international and 2020 Swedish health economic recommendations [19-21], we used an extended health care perspective and discounted costs and health benefits by 3% per year. We also included a health care perspective using direct medical costs, which reflected a recent update to the Swedish recommendations. We calculated the incremental cost-effectiveness ratios (ICERs), defined as the additional cost per QALY gained for one screening strategy compared to the next least costly non-dominated strategy. Using the 50 natural history parameter sets, we also presented acceptability curves (given a WTP threshold, the proportion of parameter sets where a strategy was cost-effective) and acceptability frontiers (given a WTP threshold τ, the strategy with the highest mean net monetary benefit (NMB), where NMB= τ*QALE-Costs).

Strategies that were more costly and less effective (strongly dominated, labelled “D”), or more effective but with a higher cost per QALY gained (extendedly dominated, labelled “ED”), were considered dominated and cost inefficient. The cost-effective strategy was identified as the strategy with the highest ICER below a commonly accepted WTP threshold. Sweden does not have an officially recognised WTP threshold; we considered WTP thresholds of €50,000 and €100,000 per QALY gained.

### Economic and health state utility parameters

Screening and treatment-related costs were taken from previous analyses [15, 22, 23] and updated for the 2020 year by using the consumer price index [24] and converted to Euro (€) using the average annual 2020 exchange rate (€1 = 10.4867 SEK) [25]. Treatment pathways for cervical cancer were defined by national guidelines (see Table 2 and Appendix A). Costs for cervical cancer treatment were calculated in relation to International Federation of Gynecology and Obstetrics (FIGO) stage 2018 [26] and mapped to SEER staging used in the simulation model (see Table 2). We included costs reflecting a woman’s time associated with screening and treatment. For all women alive in the model, we applied age-specific health utility values for the general population from a Swedish study [27], as well as stage-specific utility values following cancer diagnosis (applied multiplicatively to the general population value) (Appendix B) [28].

**Table 2.** Selected input parameters for the base case and sensitivity analyses.

| Variables | Description | Base case analysis (€) 2020 | Sensitivity analyses (€) 2020 |  |
| --- | --- | --- | --- | --- |
| Costs |  | Base case | +/- 50% | Direct medical costs |
| Clinician collected screen with liquid-based cytology (LBC) test for cytology or HPV analysis | Material, midwife office consultant time, lab analysis and patient time cost. | €107 | €53-€160 | €74 |
| Self-sampled screen for HPV analysis | Material, post, lab analysis (no cost for patient time) | €23 | €11-€34 | €23 |
| Colposcopy examination | Material, gynecological office consultant time and patient time cost and lab analysis, and productivity loss associated with management. | €328 | €164-€492 | €259 |
| Treatment of high-grade precancer | Material, gynecological office consultant time, patient time cost and lab analysis; and productivity loss associated with treatment. | €862 | €431-€1293 | €779 |
| Treatment of local cervical cancer | Diagnosis, hysterectomy, radiotherapy chemotherapy and productivity loss associated with treatment 1 year.<br>(80% FIGO 1a1-1b2, 20% 1b3-2a) | €35 660 | €17 830-€53 490 | €27 140 |
| Treatment of regional cervical cancer | Diagnosis, hysterectomy* brachytherapy, radiotherapy chemotherapy and productivity loss associated with treatment 1 year.<br>(60% FIGO 2, 40% FIGO 3) | €69 780 | €34 890-€104 670 | €52 498 |
| Treatment of distant cervical cancer | Diagnosis, radiotherapy chemotherapy and productivity losses associated with treatment 1 year.<br>(100% FIGO 4) | €68 941 | €34 471-€103 412 | 28 928 |
| Discount rate |  | 3% | 0%, 5% |  |
| Compliance | Perfect compliance to screening and follow-up procedures and treatment. | 100% | Clinician-collected strategies assumed:<br>80% compliance to primary screening.<br>80% compliance to follow-up at midwife after a positive primary HPV test.<br>96% compliance to colposcopy after primary screening with positive tests or follow-up 18 or 36 months later.<br>100% compliance to treatment after diagnosis. |  |

HSIL, high-grade intraepithelial lesions; FIGO 2018, International Federation of Gynecology and Obstetrics.
\* Treatment of regional cervical cancer includes postoperative chemoradiotherapy (FIGO stages IIIC1-2p).

### Base case analysis

To determine the preferred screening strategy constrained to clinician-collection only or for the entire choice set of clinician- or self-sampling-based screening, we performed two sets of cost-effectiveness analyses: clinician-collected sampling alone; and all screening strategies, including both clinician-collected sampling and self-sampling.

### Sensitivity analyses

For the sensitivity analyses we considered: (i) direct medical costs alone; (ii) lower compliance to screening (see Table 2 and Appendix Table D.1); (iii) lower accuracy for self-sampling (see Appendix C); (iv) 0% discount rates for costs and QALYs; (v) 5% discount rates for costs and QALYs; (vi) 50% lower costs; (vii) 50% higher costs; and (viii) life expectancy rather than quality-adjusted life expectancy as a measure of effectiveness when calculating ICERs. As per the base case, we presented results for clinician-collected sampling and for all strategies. For each of these analyses, we present a table of means, the cost-efficiency frontier and the acceptability curves.

## Results

For an unvaccinated cohort of women, the 2015 recommendations (labelled as CC-HPV(s),23y,3-7y) were projected to have an average discounted lifetime cost per woman of €832 and an average discounted lifetime QALE of 25.6822. The 2015 recommendations were less effective and more costly compared with the clinician-collected 2022 recommendations (CC-HPV,23y,5-7y), with lifetime costs of €784 and a QALE of 25.6839 (Table 3 and Figure 2). In general, more intensive screening was associated with increased costs, more colposcopies, a lower risk of cancer, and a higher proportion of cancers detected at a local stage. When we restricted our cost-effectiveness analysis to *clinician-collected sampling strategies*, there were seven “cost-efficient” strategies on the cost-effectiveness frontier (see Table 3, Figure 2, Figure F.1 and Table F.2). At a WTP of €50,000 per QALY gained, the optimal strategy involved primary HPV screening starting at age 25 with 10-yearly screening (CC-HPV,25y,10-10y), while increasing the WTP threshold identified an optimal strategy that involved screening women more frequently at younger ages (CC-HPV,25y,7-10y). While the 2022 clinician-collected recommendations were on the efficiency frontier, the ICER exceeded a WTP threshold of €100,000 per QALY gained (see Figure F.1).

**Table 3:** Summary statistics, costs, quality-adjusted life-years and ICERs on the cost-efficiency frontier, base parameters for clinician-collected strategies and all strategies.

| Strategy | Expected number of colposcopy referrals per woman over lifetime | Mean lifetime risk of cancer (%) | Proportion of cancers detected at local stage (%) | Life-years (undiscounted, from age 9 years) | Total cost per woman (€; discounted) | QALE (discounted) | Incremental cost (€; discounted) | Incremental QALEs (discounted) | ICER (Cost (€) per QALY gained) | Status |
| --- | --- | --- | --- | --- | --- | --- | --- | --- | --- | --- |
| **Clinician-collected strategies** |  |  |  |  |  |  |  |  |  |  |
| No screening | 0.000 | 1.744 | 67.10 | 75.69084 | 145.51 | 25.64434 |  |  |  | ND |
| **CC-HPV, 25y, 10-10y** | **0.786** | **0.202** | **81.66** | **75.89505** | **515.05** | **25.68114** | **369.53** | **0.03680** | **10040** | **ND** |
| **CC-HPV, 25y, 7-10y** | **0.887** | **0.173** | **83.30** | **75.89943** | **589.51** | **25.68210** | **74.46** | **0.00096** | **77594** | **ND** |
| CC-HPV, 25y, 5-10y | 1.005 | 0.142 | 84.44 | 75.90247 | 679.91 | 25.68276 | 90.40 | 0.00066 | 136229 | ND |
| CC-HPV, 23y, 5-10y | 1.151 | 0.126 | 83.79 | 75.90396 | 771.10 | 25.68337 | 91.19 | 0.00060 | 151147 | ND |
| CC-HPV, 23y, 5-7y | 1.179 | 0.115 | 85.14 | 75.90480 | 787.93 | 25.68346 | 16.83 | 0.00009 | 180913 | ND |
| CC-HPV(s), 23y, 3-7y<sup>a</sup> | 0.889 | 0.145 | 85.93 | 75.90172 | 831.74 | 25.68224 |  |  |  | D |
| CC-HPV, 23y, 3-7y<sup>b</sup> | 1.396 | 0.093 | 85.63 | 75.90667 | 988.78 | 25.68396 | 200.85 | 0.00050 | 398279 | ND |
| **All strategies** |  |  |  |  |  |  |  |  |  |  |
| No screening | 0.000 | 1.744 | 67.10 | 75.69084 | 145.51 | 25.64434 |  |  |  | ND |
| SS-HPV EG, 25y, 10-10y | 0.626 | 0.277 | 79.46 | 75.88525 | 324.01 | 25.67897 | 178.49 | 0.03463 | 5154 | ND |
| SS-HPV(r), 25y, 10-10y | 0.976 | 0.190 | 82.32 | 75.89666 | 387.64 | 25.68153 | 63.63 | 0.00256 | 24809 | ND |
| **SS-HPV(r), 25y, 7-10y** | **1.115** | **0.160** | **83.64** | **75.90093** | **432.14** | **25.68248** | **44.50** | **0.00094** | **47104** | **ND** |
| SS-HPV(r), 25y, 5-10y | 1.277 | 0.127 | 84.87 | 75.90375 | 482.20 | 25.68310 | 50.07 | 0.00062 | 81087 | ND |
| **SS-HPV(r), 25y, 5-7y** | **1.313** | **0.120** | **85.54** | **75.90440** | **489.42** | **25.68317** | **7.22** | **0.00007** | **98938** | **ND** |
| SS-HPV(r), 23y, 5-7y | 1.510 | 0.106 | 86.31 | 75.90550 | 573.15 | 25.68366 | 83.73 | 0.00049 | 169740 | ND |
| SS-HPV(r), 23y, 3-7y | 1.820 | 0.085 | 86.51 | 75.90727 | 682.63 | 25.68411 | 109.49 | 0.00045 | 242014 | ND |
| CC-HPV(s), 23y, 3-7y<sup>a</sup> | 0.889 | 0.145 | 85.93 | 75.90172 | 831.74 | 25.68224 |  |  |  | D |
| CC-HPV, 23y, 3-7y<sup>b</sup> | 1.396 | 0.093 | 85.63 | 75.90667 | 988.78 | 25.68396 |  |  |  | D |

<sup>a</sup> 2015 recommendations. <sup>b</sup> 2022 recommendations. Strategies in bold are optimal at willingness-to-pay thresholds of either €50,000 or €100,000 per QALY gained. Abbreviations: SS=Self-sampling; CC=Clinician-collected; HPV=Human papillomavirus; EG=Extended genotyping; (s)=switch from primary cytology to primary HPV testing at age 30; (r)=direct referral to colposcopy for HPV 16/18/45; ICERs= Incremental Cost-Effectiveness Ratios; ND=Not Dominated; D=Dominated; QALY=quality-adjusted life-years; QALE=quality-adjusted life expectancy.

**Figure 2.**
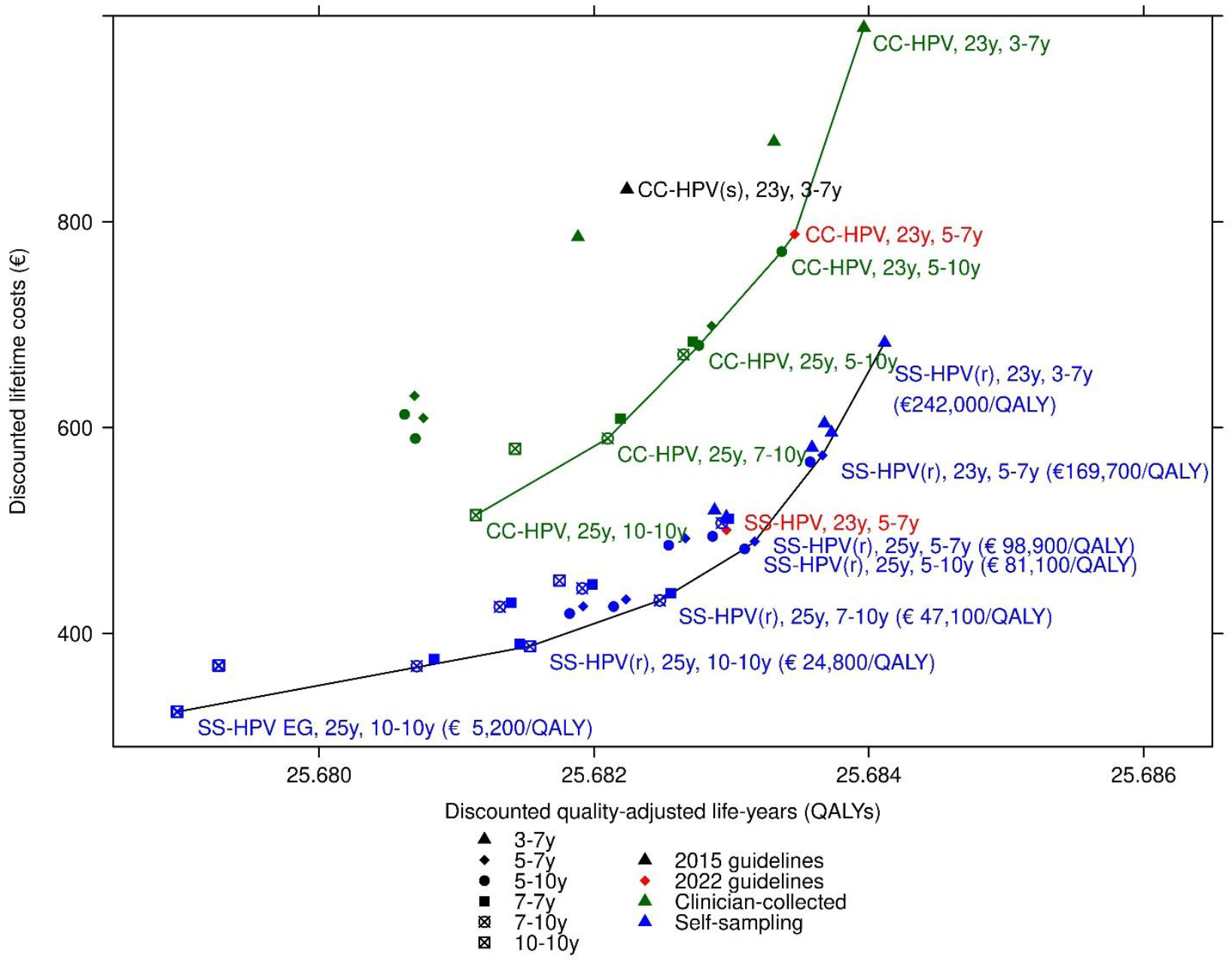
Cost-effectiveness plane for analysis reflecting unvaccinated women in Sweden. The solid green line reflects the cost-effectiveness frontier when considering the clinician-collected (CC) strategies only. The solid black line reflects cost-effectiveness frontier when considering all strategies, i.e., clinician-collected and self-sampling strategies. Abbreviations: SS=Self-sampling; CC=Clinician-collected; HPV=Human papillomavirus; EG=Extended genotyping; (s)=switch from primary cytology to primary HPV testing at age 30; (r)=direct referral to colposcopy for HPV 16/18/45.

When we compared both *clinician-collected sampling and self-sampling strategies*, we identified eight strategies on the frontier (Table 3, Figures 2-3). At the lower WTP threshold, the optimal strategy involved self-sampling starting at age 25 (with direct referral to colposcopy for HPV 16/18/45), with 7- and 10-yearly screening for women aged <50 and ≥50 years, respectively. A higher WTP threshold identified an optimal strategy involving self-sampling more frequently, i.e., 5- and 7-yearly screening for women aged <50 and ≥50 years, respectively. The probability these strategies were cost-effective ranged from 48-58% (Figure 3). In general, for strategies with comparable start ages and screening intervals, we found that the clinician-collected sampling strategies were dominated by, or had ICERs that exceeded €88,000 per QALY gained, compared with the self-sampling strategies (Table F.1).

**Figure 3:**
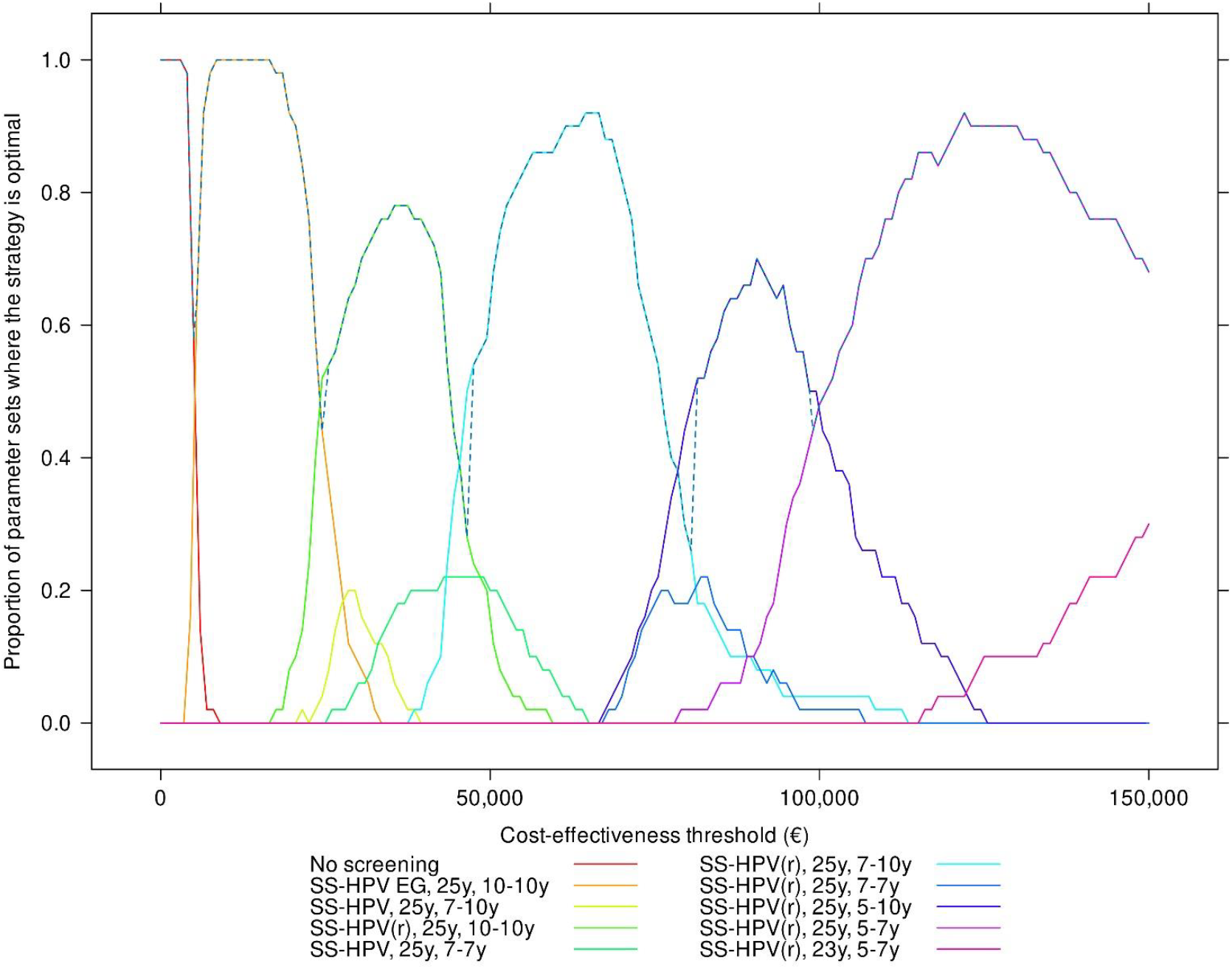
Acceptability curves for different willingness-to-pay thresholds for cervical cancer screening strategies in unvaccinated cohorts in Sweden, all strategies (the dashed line reflects the cost-effectiveness acceptability frontier) Abbreviations: SS=Self-sampling; HPV=Human papillomavirus; EG=Extended genotyping; (r)=direct referral to colposcopy for HPV 16/18/45.

The markedly improved discrimination associated with extended genotyping was offset by increased costs. For example, self-sampling from age 25 both with and without extended genotyping had very similar outcomes for 7- and 10-yearly screening (SS-HPV,25y,7-10y vs SS-HPV EG,25y,7-10y) and for 10-yearly screening (SS-HPV,25y,10-10y vs SS-HPV EG,25y,10-10y). We also found that strategies starting screening from age 25 were more cost-effective compared with starting at age 23 for WTP thresholds below €100,000 per QALY gained.

### Sensitivity analyses

For strategies based on clinician-collected sampling (Appendix G), assuming direct medical costs only, lower compliance, and reducing costs by 50% generally resulted in optimal strategies that were more intensive, i.e., more frequent screening intervals or a younger starting age, than in our base case. For example, at a WTP of €50,000 per QALY and assuming either direct medical costs, lower compliance or lower costs, the optimal strategy involved screening women aged <50 years every 7 years rather than every 10 years as in our base case. For the other sensitivity analyses, the optimal strategies did not change.

For the full set of strategies (Appendix H): for direct medical costs and for life expectancy as the outcome, the optimal strategies did not change; for lower compliance, the optimal strategy at €50,000 was SS-HPV(r),25y,5-7y, while the optimal strategy at €100,000 was SS-HPV(r),25y,3-7y; for undiscounted QALYs and costs, the only strategy on the frontier below €100,000 was SS-HPV EG,25y,10-10y, which had an ICER of €21,100 per QALY gained; for 5% discounting, the optimal strategy at €50,000 was SS-HPV(r),25y,10-10y, while SS-HPV(r),25y,7-10y was optimal at €100,000; lower costs supported more intensive strategies, whereas higher costs were associated for less intensive strategies; and, finally, for lower accuracy for self-sampling, the optimal strategies were for clinician-collected sampling, with CC-HPV,25y,10-10y being optimal at €50,000 and CC-HPV,25y,7-10y being optimal at €100,000.

## Discussion

Given our base-case assumptions of perfect compliance and equal diagnostic accuracy of HPV analysis for clinician-collected sampling and self-sampling, we found that strategies involving self-sampling were associated with lower costs and similar health benefits compared with clinician-collected sampling. When we restricted the cost-effectiveness analysis to include clinician-collected sampling strategies only, the 2022 Swedish recommendations were on the cost-efficiency frontier; however, less frequent screening than is currently recommended was considered optimal at €100,000 per QALY gained. When we expanded the analysis to include both clinician-collected and self-sampling strategies, we found that for the same WTP threshold, a self-sampling-based program enabled screening more frequently, contributing to greater health benefits but also lower costs than the optimal (less intensive) screening frequency identified in the clinician-collected analysis. Importantly, these findings were sensitive to the assumed test accuracy for self-sampling, where clinician-collected sampling was preferred when we assumed that self-sampling was considerably less accurate.

Our modelling approach has several advantages. The simulation model has been shown to have good validity [29, 30]. Moreover, our consultation with key stakeholders ensured that we were focused on the key strategies for the Swedish health care setting. However, our analysis has several limitations. Firstly, due to lack of empirical data, we assumed full compliance in the base case, which is optimistic [18]. If compliance were lower, we predicted that more frequent screening would be optimal. Second, the key policy questions shifted during our modelling effort – specifically, key stakeholders suggested that partial genotyping was of less interest than extended genotyping. Our model predicted that the improved test characteristics did not offset the increased costs associated with extended genotyping, although this finding may be sensitive to the cost of extended genotyping.

Before replacing clinician-collecting sampling with self-sampling, policy-makers need to consider potential barriers, including (a) women’s acceptance of self-sampling, (b) the method of vaginal sampling, (c) the choice of lab analysis, and (d) other logistical issues. Previous studies in high-risk women have shown that self-sampling has high acceptance [31]. However, it is important to ensure safety by monitoring the adherence to follow-up after a positive self-sampling.

## Conclusion

For unvaccinated women within the Swedish cervical cancer screening program, primary HPV self-sampling strategies are projected to be less costly with similar health benefits compared to primary HPV testing with clinician-collected samples. Within the context of Swedish cost-effectiveness framework, the self-sampling screening strategies with direct referral to colposcopy for women positive for HPV 16/18/45 should be considered in future decision-making of population-based screening programs for cervical cancer.

## Data Availability

All relevant data are within the manuscript and its Supporting Information files.

## Acknowledgements

We are grateful to Cecilia Ranhem for discussing the candidate strategies, Dr Miriam Elfström for discussion on national recommendations and implementation, and Professors Jane Kim and Pär Sparén for discussions.

## References

[1] Meijer CJ, Berkhof J, Castle PE, Hesselink AT, Franco EL, Ronco G, et al. Guidelines for human papillomavirus DNA test requirements for primary cervical cancer screening in women 30 years and older. Int J Cancer. 2009;124:516–20.

[2] Ronco G, Dillner J, Elfström KM, Tunesi S, Snijders PJ, Arbyn M, et al. Efficacy of HPV-based screening for prevention of invasive cervical cancer: follow-up of four European randomised controlled trials. Lancet. 2014;383:524–32.

[3] Hellsten C, Ernstson A, Bodelsson G, Forslund O, Borgfeldt C. Equal prevalence of severe cervical dysplasia by HPV self-sampling and by midwife-collected samples for primary HPV screening: a randomised controlled trial. Eur J Cancer Prev. 2021;30:334–40.

[4] Arbyn M, Smith SB, Temin S, Sultana F, Castle P, Testing CoS-SaH. Detecting cervical precancer and reaching underscreened women by using HPV testing on self samples: updated meta-analyses. BMJ. 2018;363:k4823.

[5] International Agency for Research on Cancer. Human papillomaviruses. Lyon: International Agency for Research on Cancer; 1995.

[6] Clifford GM, Smith JS, Aguado T, Franceschi S. Comparison of HPV type distribution in high-grade cervical lesions and cervical cancer: a meta-analysis. Br J Cancer. 2003;89:101–5.

[7] de Sanjose S, Quint WG, Alemany L, Geraets DT, Klaustermeier JE, Lloveras B, et al. Human papillomavirus genotype attribution in invasive cervical cancer: a retrospective cross-sectional worldwide study. Lancet Oncol. 2010;11:1048–56.

[8] Arbyn M, Depuydt C, Benoy I, Bogers J, Cuschieri K, Schmitt M, et al. VALGENT: A protocol for clinical validation of human papillomavirus assays. J Clin Virol. 2016;76 Suppl 1:S14–S21.

[9] Bonde JH, Sandri MT, Gary DS, Andrews JC. Clinical utility of human papillomavirus genotyping in cervical cancer screening: A systematic review. J Low Genit Tract Dis. 2020;24:1–13.

[10] Nationellt Kvalitetsregister för Cervixcancerprevention. Förebyggande av livmoderhalscancer i Sverige Verksamhetsberättelse och Årsrapport 2021 med data till och med 2020. Stockholm: Nationellt Kvalitetsregister för Cervixcancerprevention; 2021.

[11] Andersson S, Belkić K, Demirbüker SS, Mints M, Östensson E. Perceived cervical cancer risk among women treated for high-grade cervical intraepithelial neoplasia: The importance of specific knowledge. PLoS One. 2017;12:e0190156.

[12] Östensson E, Belkić K, Ramqvist T, Mints M, Andersson S. Self-sampling for high-risk human papillomavirus as a follow-up alternative after treatment of high-grade cervical intraepithelial neoplasia. Oncol Lett. 2021;21:240.

[13] Andersson S, Belkić K, Mints M, Östensson E. Is self-sampling to test for high-risk papillomavirus an acceptable option among women who have been treated for high-grade cervical intraepithelial neoplasia? PLoS One. 2018;13:e0199038.

[14] Pedersen K, Portnoy A, Sy S, Hansen BT, Tropé A, Kim JJ, et al. Switching clinic-based cervical cancer screening programs to human papillomavirus self-sampling: A cost-effectiveness analysis of vaccinated and unvaccinated Norwegian women. Int J Cancer. 2022;150:491–501.

[15] Fogelberg S, Clements MS, Pedersen K, Sy S, Sparén P, Kim JJ, et al. Cost-effectiveness of cervical cancer screening with primary HPV testing for unvaccinated women in Sweden. PLoS One. 2020;15:e0239611.

[16] Malone C, Barnabas RV, Buist DSM, Tiro JA, Winer RL. Cost-effectiveness studies of HPV self-sampling: A systematic review. Prev Med. 2020;132:105953.

[17] Campos NG, Burger EA, Sy S, Sharma M, Schiffman M, Rodriguez AC, et al. An updated natural history model of cervical cancer: derivation of model parameters. Am J Epidemiol. 2014;180:545–55.

[18] Pedersen K, Kristiansen IS, Sy S, Kim JJ, Burger EA. Designing guidelines for those who do not follow them: The impact of adherence assumptions on optimal screening guidelines. Value Health. 2023;26:1217–24.

[19] Drummond MF, Sculpher MJ, Claxton K, Stoddart GL, Torrance GW. Methods for The Economic Evaluation of Health Care Programmes. Oxford: Oxford University Press; 2015.

[20] National Board of Health and Welfare. Nationella Riktlinjer för Hjärtsjukvård. Hälsoekonomiskt Underlag Bilaga. Stockholm: National Board of Health and Welfare; 2018.

[21] Tandvårds-och Läkemedelsförmånsverkets. Tandvårds-och Läkemedelsförmånsverkets Allmänna Råd. TLVAR 2017:1 Ändring i Tandvårds-och Läkemedelsförmånsverkets Allmänna råd (TLVAR 2003:2) om Ekonomiska Utvärderingar. Stockholm 2017.

[22] Östensson E, Hellström AC, Hellman K, Gustavsson I, Gyllensten U, Wilander E, et al. Projected cost-effectiveness of repeat high-risk human papillomavirus testing using self-collected vaginal samples in the Swedish cervical cancer screening program. Acta Obstet Gynecol Scand. 2013;92:830–40.

[23] Östensson E, Froberg M, Leval A, Hellström AC, Backlund M, Zethraeus N, et al. Cost of preventing, managing, and treating human papillomavirus (HPV)-related diseases in Sweden before the introduction of quadrivalent HPV vaccination. PLoS One. 2015;10:e0139062.

[24] Statistics Sweden. CPI, Fixed Index Numbers. Statistics Sweden; 202.

[25] Sveriges Riksbank. Startsida.

[26] Bhatla N, Aoki D, Sharma DN, Sankaranarayanan R. Cancer of the cervix uteri. Int J Gynaecol Obstet. 2018;143 Suppl 2:22–36.

[27] Burström K, Johannesson M, Diderichsen F. Swedish population health-related quality of life results using the EQ-5D. Qual Life Res. 2001;10:621–35.

[28] Myers ER, Green S, Lipkus I. Patient preferences for health states related to HPV infection: visual analogue scale versus time tradeoff elicitation. 21st International Papillomavirus Conference. Mexico City, Mexico 2004.

[29] Kim JJ, Burger EA, Regan C, Sy S. Screening for cervical cancer in primary care: A decision analysis for the US Preventive Services Task Force. JAMA. 2018;320:706–14.

[30] Portnoy A, Pedersen K, Trogstad L, Hansen BT, Feiring B, Laake I, et al. Impact and cost-effectiveness of strategies to accelerate cervical cancer elimination: A model-based analysis. Prev Med. 2021;144:106276.

[31] Andersson S, Belkic K, Mints M, Ostensson E. Acceptance of Self-Sampling Among Long-Term Cervical Screening Non-Attenders with HPV-Positive Results: Promising Opportunity for Specific Cancer Education. J Cancer Educ. 2021;36:126–33.

