## Supplementary Material for "Cost-effectiveness of human papillomavirus self-sampling in the Swedish cervical screening program"

### Supplementary Material: Cost-effectiveness of HPV self-sampling in the Swedish cervical cancer screening program

Affiliations:
[1]Department of Medical Epidemiology and Biostatistics, Karolinska Institutet, Stockholm, Sweden.
[2]Department of Women's and Children's Health, Karolinska Institutet, Stockholm, Sweden.
[3] Department of Obstetrics and Gynaecology, Lund University, Lund, Sweden.
[4] Department of Gynecologic Cancer, Theme Cancer, Karolinska University Hospital in Solna and Dept of Oncology and Pathology, Karolinska Institutet, Stockholm, Sweden.
[5] Department of Health Management and Health Economics, University of Oslo, Oslo, Norway.
[6] Center for Health Decision Science, Harvard T. H. Chan School of Public Health, Boston, Massachusetts, United States of America.

#### Contents

**A Costs** 2

**B Health state utility values** 8

**C Diagnostic accuracy assumptions** 8

**D Screening algorithms and follow-up management** 9

**E Imputation for missing simulations** 9

**F Base analysis** 14

### A Costs

Costs were taken from previous published studies (Östensson, Hellstrom et al. 2013; Ostensson, Froberg et al. 2015; Fogelberg, Clements et al. 2020) and updated for treatment of cervical cancer by clinical gynecology and oncology experts Kristina Hellman and Henric Falconer at Karolinska University Hospital, Stockholm, Sweden to reflect national medical practice in 2020. Cervical cancer treatment costs were primarily calculated by using the cost-per-patient (KPP Database[1]) estimates from Karolinska University hospital, Solna-Stockholm, Sweden, which treats the majority of Swedish patients diagnosed with cervical cancer. We included indirect costs calculated from the loss of production using a human capital approach (Drummond et al. 2015) reflecting costs for women's time associated with screening, management, and treatment of diagnosed cervical precancerous lesions and cancer. We assumed full employment and used the average monthly salary of the general population in Sweden (36 100 SEK) in 2020[2], with an additional 37% for employer and social contributions[3]. When needed, all costs were adjusted to calendar year 2020 by using the consumer price index[4] and converted to Euro (€) using the average annual 2020 exchange rate (€1 = 10,4867 SEK) from the national bank[5].

[1]CPP Kostnad per patient database, Sveriges Kommuner och Regioner, https://skr.se/skr/halsasjukvard/ekonomiavgifter/kostnadperpatientkpp.1076.html. Accessed 2022-07-06.

[2]Salary structures, whole economy. Statistics Sweden; 2020. http://www.statistikdatabasen.scb.se/pxweb/en/ssd/START__AM__AM0110__AM0110A/LonYrkeUtbildning4A/. Accessed 2022-07-06.

[3]Carlgren F. Sociala avgifter över tid - Avtalade och lagstadgade avgifter - arbetare. ekonomifakta. https://www.ekonomifakta.se/Fakta/Skatter/Skatt-pa-arbete/Sociala-avgifter-over-tid/. Published 2020. Updated 2020-06-30. Accessed 2022-07-06.

[4]SCB. CPI, Fixed Index Numbers (1980=100). Statistics Sweden. https://www.scb.se/en/finding-statistics/statistics-by-subject-area/prices-and-consumption/consumer-price-index/consumer-price-index-cpi/pong/tables-and-graphs/consumer-price-index-cpi/cpi-fixed-index-numbers-1980100/. Published 2020. Updated 2020-11-12. Accessed 2022-07-06.

[5]Annual average exchange rates. Swedish National Bank; 2020. https://www.riksbank.se/en-gb/statistics/search-interest--exchange-rates/annual-average-exchange-rates/. Ac-

cessed 2022-07-06.

| Variables | Description | Base case analysis SEK, (€) 2020 | Uncertainty analysis, +/- 50%, SEK, (€) 2020 | Direct medical costs SEK, (€) 2020 |
|---|---|---|---|---|
| Clinician-collected screen with liquid-based cytology (LBC) test for cytology or HPV analysis | Material, midwife office consultant time, lab analysis and patient time cost. | €107 | €53–160 | €74 |
| Self-sampled screen for HPV analysis | Material, post, lab analysis (no cost for patient time) | €23 | €11–34 | €23 |
| Colposcopy examination | Material, gynecological office consultant time and patient time cost and lab analysis, and productivity loss associated with management. | €328 | €164–492 | €259 |
| Treatment of CIN | Material, gynecological office consultant time, patient time cost and lab analysis. and productivity loss associated with treatment. | €862 | €431–1293 | €779 |
| Treatment of local cervical cancer | Diagnosis, hysterectomy, radiotherapy, chemotherapy and productivity losses associated with treatment 1 year (80% FIGO 1a1-1b2, 20% 1b3-2a). | €35 660 | €17 830–53 490 | €27 140[6] |
| Treatment of regional cervical cancer | Diagnosis, hysterectomy brachytherapy, radiotherapy chemotherapy and productivity loss associated with treatment 1 year (60% Figo 2, 40% Figo 3). | €69 780 | €34 890–104 670 | €52 498[7] |
| Treatment of distant cervical cancer | Diagnosis, radiotherapy chemotherapy and productivity loss associated with treatment 1 year (100% Figo 4). | €68 941 | €34 471–103 412 | €28 928[8] |

Table A.1: Cost estimates.

[6]Including 3% discounted direct medical cost for follow-up by physician for 5 years calculated on stage specific survival. From Cancer Centrum. https://cancercentrum.se/globalassets/cancerdiagnoser/gynekologi/kvalitetsregister/nationell-kvalitetsrapport-gynekologisk-cancer-2018.pdf. Accessed 2022-07-06.

[7]See footnote 6.

[8]See footnote 6.

| Treatment variables | Quantity | Costs (€) 2020 |
|---|---|---|
| Pretreatment evaluation including clinical examination without anesthesia, MRT and DT abdomen/thorax, MDK, start of care plan, enrollment call. | 1 | 2 578 |
| Surgery (radical hysterectomy/trachelectomy/conization) * | 1 | 12 485 |
| Follow-up of treatment regimen with a clinical examination without anesthesia with DT abdomen/thorax and 1 physician visit | 1 | 939 |
| Total direct cost first year | | 16 002 |
| Follow-up visits for 5 years** | | 4 050 |
| Sick leave first year (weeks) | 9 | 10 611 |
| Total societal cost after 1st year | | 26 613 |

Table A.2: Cost for treatment, management and follow-up of cervical cancer: FIGO stage IAI–IB2

| Treatment variables | Quantity | Costs (€) 2020 |
|---|---|---|
| Pretreatment evaluation with examination of the patient without anesthesia and examination under general anesthesia to clinically assess tumor extension (visual examination), imaging with MRT and PET/DT, MDK, start of care plan, enrollment call | 1 | 4 280 |
| Brachytherapy, with 4 MRT pre-treatment each BT treatment 4 BT and bed place | 4 | 27 177 |
| Radiotherapy with 1 DT and 1 MR pre-treatment and 1 physician visit | 25 | 9 395 |
| Chemotherapy and follow up during treatment with 3 physician visit and 2 follow-up phone calls by health care personnel | 6 | 7 767 |
| Follow up after treatment regimen by oncologist specialist with 1 FDG-PET/DT + 1 DT and 2 MRT and 1 follow-up phone calls by health care personnel and 1 physician visit | 1 | 3 185 |
| Total direct cost 1st year | | 51 804 |
| Follow-up visits for 5 years | | 3 977 |
| Sick leave first year (weeks) | 17 | 20 044 |
| Total societal cost after 1st year | | 71 847 |

Table A.3: Cost for treatment, management and follow-up of cervical cancer: FIGO stage IB3–IV

| **Treatment variables** | **Quantity** | **Costs (€) 2020** |
|---|---|---|
| Pretreatment evaluation with examination of the patient under general anesthesia to clinically assess tumor extension (visual examination), imaging with MRT and PET/DT, MDK, start of care plan, enrollment call | 1 | 4280 |
| Brachytherapy, with 1 MRT pre-treatment) for each BT treatment (total 3) and 3 bed places | 3 | 20 383 |
| Radiotherapy including 1 DT and 1 MR pre-treatment and 1 physician visit | 30 | 11 022 |
| Chemotherapy + follow up during treatment with 3 physician visit and 2 follow-up phone calls by health care personnel | 6 | 7767 |
| Follow up after treatment regimen by physician with FDG-PET/DT + 1 DT and 2 MRT and 1 follow-up phone calls by health care personnel, 1 physician visit | 2 | 3185 |
| Total direct cost 1st year | | 46 636 |
| Follow-up for 5 years | | 1 920 |
| Sick leave 1st year (weeks) | 17 | 20 044 |
| Total societal cost after 1st year | | 66 679 |

Table A.4: Cost for treatment, management and follow-up of cervical cancer: FIGO 3

| **Treatment variables** | **Quantity** | **Costs (€) 2020** |
|---|---|---|
| Pretreatment evaluation with examination of the patient under general anesthesia in order to clinically assess tumor extension (visual examination), imaging with MRT and PET/DT, MDK, start of care plan, enrollment call | 1 | 4 280 |
| Radiotherapy with1 DT and 1 MR pre-treatment, 1 physician visit | 10 | 4 516 |
| Palliative care, Taxol cisplatin chemotherapy | 6 | 15 595 |
| Follow-up during radio chemotherapy with clinical examination under anesthesia and physician assessment | 1 | 1 084 |
| Follow up with physician after treatment regimen with 1 DT abdomen/thorax and 1 phone call by health care personnel and 2 physician visits | 3 | 2 200 |
| Total direct cost 1st year | | 27 675 |
| Follow-up for 5 years | | 1 254 |
| Sick leave 1st year (weeks) | 35 | 41 266 |
| Total societal cost after 1st year | | 68 941 |

Table A.5: Cost for treatment, management and follow-up of cervical cancer: FIGO 4

### B Health state utility values

According to Swedish guidelines for health economic analysis [9], we applied age-specific background utilities based on data from Sweden [10] (Table B.1). For women diagnosed with cervical cancer, an overall utility was calculated by the background utility multiplied by a stage-specific utility for a five-year period [11] (Table B.2).

| Age group (years) | 0 | 19- | 31- | 41- | 51- | 60- | 71- | 81- | 100- |
|---|---|---|---|---|---|---|---|---|---|
| Utility | 1 | 0.906 | 0.870 | 0.846 | 0.811 | 0.811 | 0.808 | 0.730 | 0.730 |

Table B.1: Age-specific background utilities, Sweden

| Stage | No Cancer | Local cancer | Regional Cancer | Distant Cancer | Dead |
|---|---|---|---|---|---|
| Utility | 1.00 | 0.90 | 0.74 | 0.48 | 0.00 |

Table B.2: Stage-specific utility for a five-year period

### C Diagnostic accuracy assumptions

**HPV test sensitivity and specificity:** For the base case analysis, sensitivity and specificity of both clinician-collected sampling and self-sampling were assumed to be 100%. Under a sensitivity analysis, we assumed that self-sampling had: 96.9% specificity for the healthy state and for low-risk HPV infections; 90% sensitivity for women with an HPV infection and no lesion; and 96% sensitivity for women with high-risk HPV infections and a lesion.

**Cytology sensitivity and specificity:** a 73% sensitivity (92% specificity) of cytology to detect (exclude) ASC-US or more severe given presence (absence) of CIN2+.[12]

**Diagnostic colposcopy accuracy:** 86% sensitivity (89% specificity) of colposcopy with biopsy to detect (exclude) CIN2+ given presence (absence) of CIN2+. [13]

---

[9] Läkemedelsförmånsverket T-o. Tandvårds- och Läkemedelsförmånsverkets Allmänna Råd. TLVAR 2017:1 Ändring i Tandvårds- och Läkemedelsförmånsverkets Allmänna råd (TLVAR 2003:2) om Ekonomiska Utvärderingar. Stockholm 2017.

[10] Burström K, Johannesson M, Diderichsen F. Swedish population health-related quality of life results using the EQ-5D. Qual Life Res. 2001;10(7):621-35. doi: 10.1023/a:1013171831202. PMID: 11822795.

[11] Myers et al (2004). Patient preferences for health states related to HPV infection: visual analogue scale versus time trade-off elicitation [abstract]. Proceeding of the 21st International Papillomavirus Conference. Mexico City

[12] Koliopoulos et al (2007). Diagnostic accuracy of human papillomavirus testing in primary cervical screening: a systematic review and meta-analysis of non-randomized studies. *Gynecol Oncol* 104(1):232–46.

[13] Stoler et al (2015). The interpretive variability of cervical biopsies and its relationship to HPV status. *Am J SurgPathol* 39:729-–736.

### D Screening algorithms and follow-up management

Guidelines are based on the National Working Group for Cervical Cancer Prevention[14].

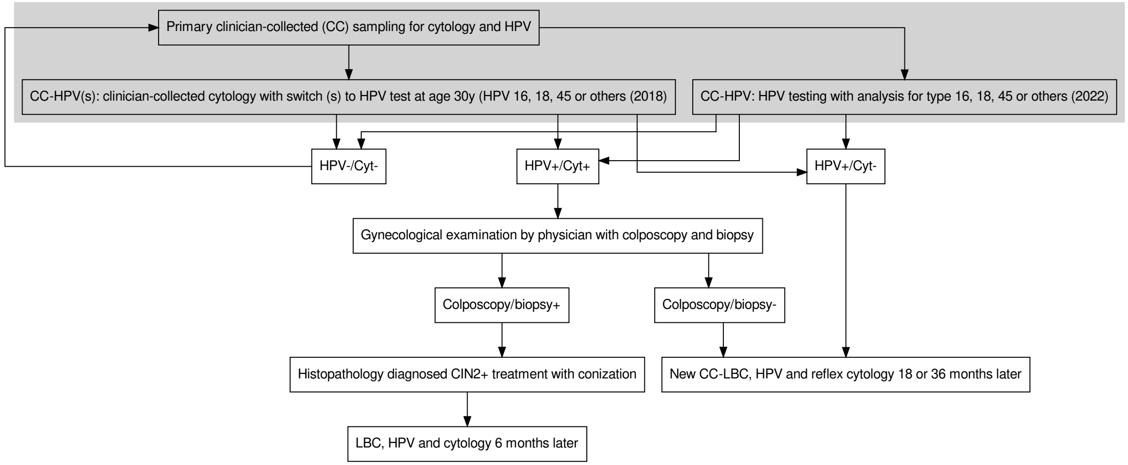

Figure D.1: Clinician-collected screening algorithms in the model

### E Imputation for missing simulations

Outcomes were imputed for the dropped simulations based on the specific strategy and specific parameter set. The missingness pattern was characterised by either all or none of the outcomes being available. Multiple imputation was implemented assuming multivariate linear regression with intercept terms, 54 main effects for the strategy and 49 main effects for the parameter set. The correlated error structure was sampled assuming a multivariate-normal distribution. As a methodological aside, the `mice` package in R, which is based on

[14] Nationella Arbetsgruppen för Cervixcancerprevention. *Livmoderhalscancerprevention Nationellt Vårdprogram (No. version 4.0)*. Stockholm, 2022. https://nkcx.se/templates/nationellt-vardprogram-cervixcancerprevention.pdf, accessed 2024-02-23.

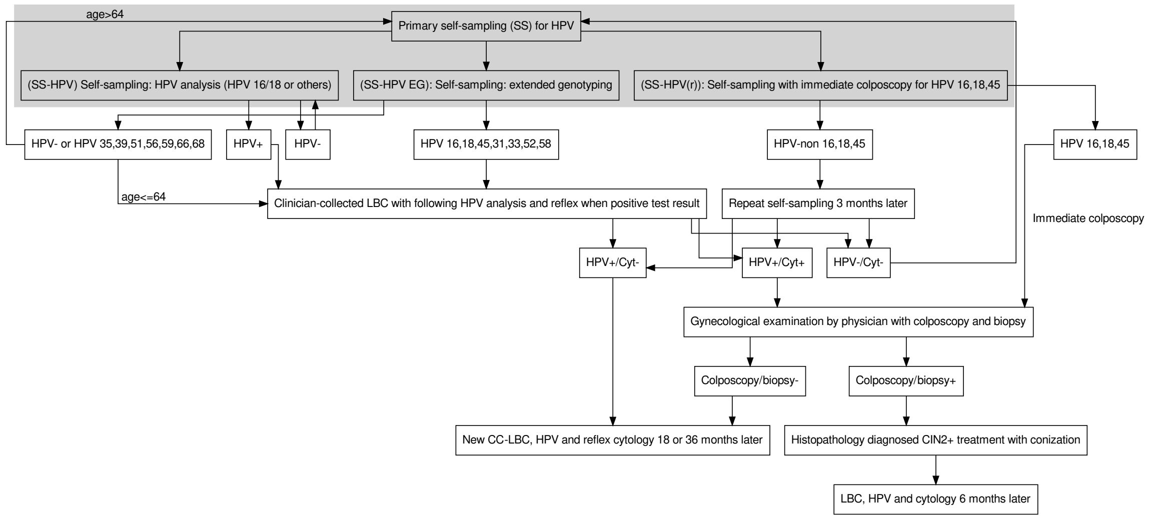

Figure D.2: Self-sampling screening algorithms in the model

| | | **Follow-up age** | | | | | | | | | | | | | | | | | | | | | | | | | | | | |
|---|---|---|---|---|---|---|---|---|---|---|---|---|---|---|---|---|---|---|---|---|---|---|---|---|---|---|---|---|---|---|
| **Years** | **Start age** | **23** | **25** | **26** | **28** | **29** | **30** | **31** | **32** | **33** | **34** | **35** | **37** | **38** | **39** | **40** | **41** | **43** | **44** | **45** | **46** | **47** | **48** | **49** | **50** | **57** | **60** | **64** | **70** | **Total** |
| 3-7y | 23 | 1 | | 1 | | 1 | | | 1 | | | 1 | | 1 | | | 1 | | 1 | | | 1 | | | 1 | 1 | | 1 | | 12 |
| 3-7y | 25 | | 1 | | 1 | | | 1 | | | 1 | | 1 | | | 1 | | 1 | | | 1 | | | 1 | | 1 | | 1 | | 11 |
| 5-7y | 23 | 1 | | | 1 | | | | | 1 | | | | 1 | | | | 1 | | | | | 1 | | | 1 | | 1 | | 8 |
| 5-7y | 25 | | 1 | | | | 1 | | | | | 1 | | | | 1 | | | | 1 | | | | | 1 | 1 | | 1 | | 8 |
| 7-7y | 23 | 1 | | | | | 1 | | | | | | 1 | | | | | | 1 | | | | | | | 1 | | 1 | | 6 |
| 7-7y | 25 | | 1 | | | | | | 1 | | | | | | 1 | | | | | | 1 | | | | | 1 | | 1 | | 6 |
| 5-10y | 23 | 1 | | | 1 | | | | | 1 | | | | 1 | | | | 1 | | | | | 1 | | | | 1 | | 1 | 8 |
| 5-10y | 25 | | 1 | | | | 1 | | | | | 1 | | | | 1 | | | | 1 | | | | | 1 | | 1 | | 1 | 8 |
| 7-10y | 23 | 1 | | | | | 1 | | | | | | 1 | | | | | | 1 | | | | | | | | 1 | | 1 | 6 |
| 7-10y | 25 | | 1 | | | | | | 1 | | | | | | 1 | | | | | | 1 | | | | | | 1 | | 1 | 6 |
| 10-10y | 23 | 1 | | | | | | | | 1 | | | | | | | | 1 | | | | | | | | | 1 | | 1 | 5 |
| 10-10y | 25 | | 1 | | | | | | | | | 1 | | | | | | | | 1 | | | | | | | 1 | | 1 | 5 |

Table D.1: Screen frequency based on a previous negative screen

| **Index result** | **Follow-up** |
|---|---|
| Cytology normal at index screen 23-29 years | Referral to routine screen 3 years later. |
| HPV-negative at index screen $\geq$30 years | Referral to routine screen 3 or 7 years later. |

Table D.2: Guidelines used for follow-up management (2018)

| **Index result** | **Follow-up with reflex HPV** | **Recommendation** |
|---|---|---|
| HSIL | No reflex HPV test | Colposcopy within 3 months |
| ASCUS/LSILcyt | Reflex HPV | HPV positive women with ASCUS/LSILcyt : $<$ 28 years referal to regular screening/no follow-up is needed; $\geq$28 years — Colposcopy within 3 months |

Table D.3: Guidelines for follow-up of cytology abnormal women below 30 years (2018)

| **Index result** | **Second test at midwife** | **Follow-up test result** | **Referral to gynaecologist** |
|---|---|---|---|
| HPV 16/18 –positive/Normal cytology | Follow-up with a second HPV test $>$ 18 months after index test | Persistent HPV 16/18 -positive | Colposcopy within 3 months |
| HPV non16/18 –positive/Normal cytology | Follow-up with a second HPV test $>$ 36 months after index test | Persistent non16/18 HPV-positive | Colposcopy within 3 months |

Table D.4: Guidelines for follow-up of HPV index screen positive with reflex cytology showing “normal” aged 30 years and over (2018)

univariate imputations, did not cope well with this missingness pattern. We used 100 imputations and then averaged different estimands across the parameter sets and strategies.

| ASCUS LSIL cyt and Normal colp | Post-colposcopy management |
|---|---|
| Normal colp, HPV neg, cyt neg | Referred to regular screening. |
| Normal colp, HPV neg, ASCUS or LSILcyt | Referred to regular screening. |
| Normal colp, HPV pos, cyt neg | Referred for further follow-up based on the HPV type (HPV16/18-18 months later/ HPV non16/18-36 months later). |
| Normal colp, HPV pos, cyt pos | Referred for further colp 3 months later |

Table D.5: Guidelines for follow-up of ASCUS LSIL with co-test and a normal colposcopy examination (no CIN has been diagnosed by colposcopic-directed cervical biopsies)

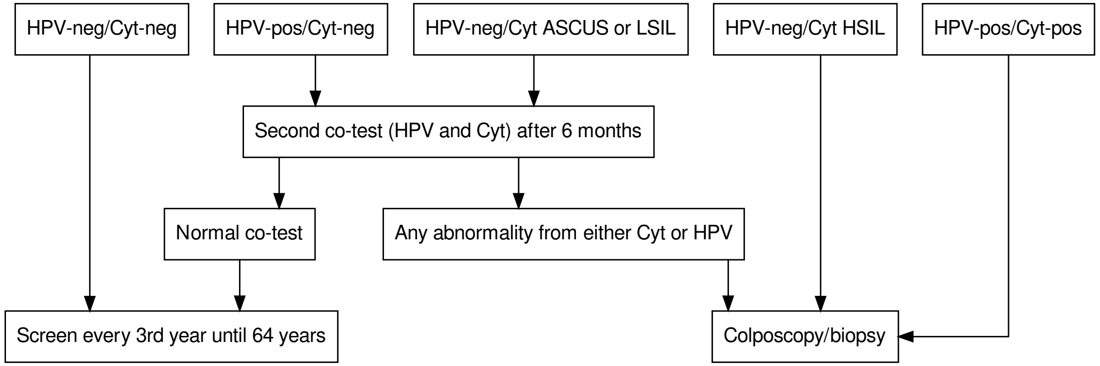

Figure D.3: Flow chart for follow up management after treatment at 6 months (first follow-up) with co-test OR when screened every 3rd year with co-test (HPV and Cytology).

| **1st follow-up after treatment of CIN2+ /OR at screen pos after treatment** | **1st Co-test result** | **Follow-up according to guidelines (2018 and 2020+)** | **2nd Co-test result** |
|---|---|---|---|
| Follow-up with a co-test (HPV and cytology) 6 months after treatment/ OR at screen every 3rd year by a midwife | HPV negative and cytology normal. | Continuous follow-up every 3 rd year with co-test (HPV and cyto) until she exits the screening program (pProcedure performed by a midwife). | |
| | HPV neg and ASCUS/LSILcyt | Follow-up with a second co-test after 6 months (no colp/biop) (procedure performed by a midwife). | Any abnormality at 2nd test is referred to colp. |
| | HPV neg and HSILcyt/ASC-H | Colposcopy within 3 months and biopsy (procedure performed by a gynecologist). | |
| | HPV positive and normal cytology. | Follow-up with a second co-test after 6 months (no colp/biopsy) (procedure performed by a midwife). | Any abnormality at 2nd test is referred to colp. |
| | HPV positive and ASCUS/LSILcyt/HSILcyt | Colposcopy within 3 months and biopsy. Procedure performed by a gynecologist. | |

Table D.6: Guidelines for follow-up after treatment OR after screen positive after treatment (2018 and 2020+)

### F Base analysis

#### F.1 Base analysis restricted to clinician-collected sampling

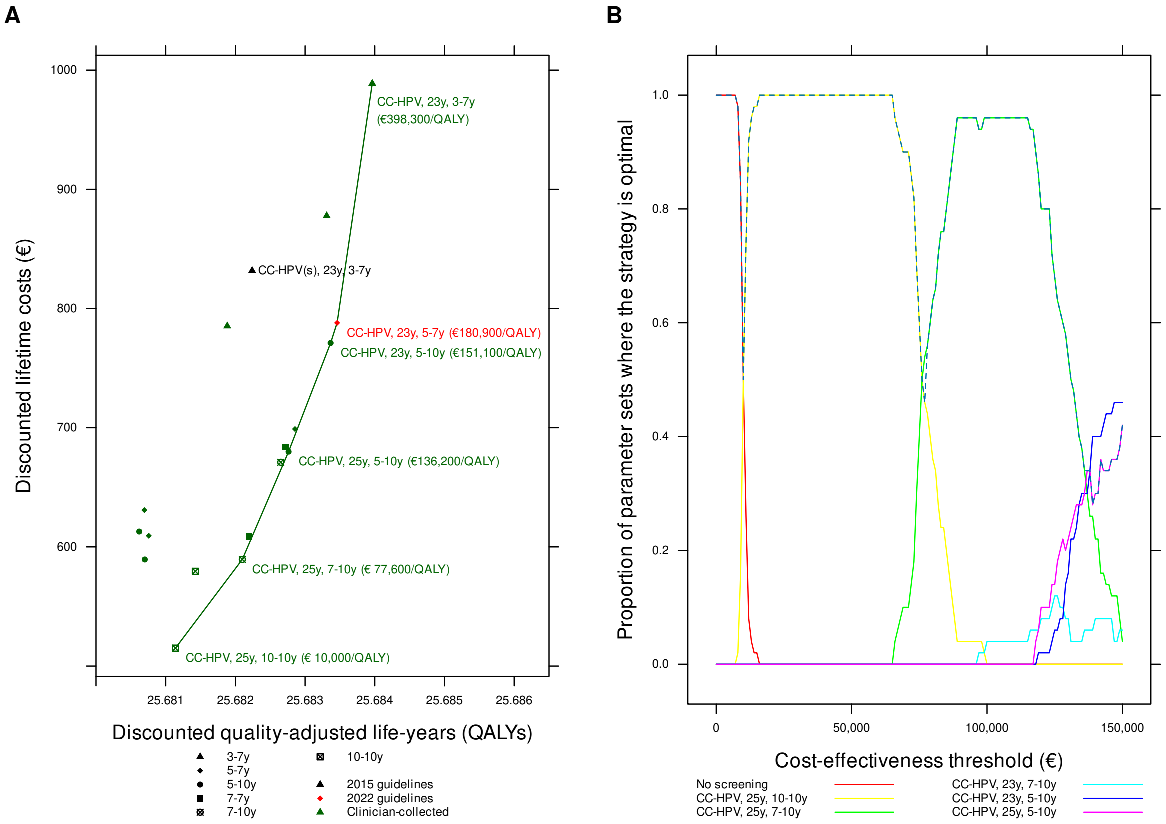

Figure F.1: Cost-effectiveness of cervical cancer screening strategies in unvaccinated cohorts in Sweden: *clinician-collected sampling*. Panel A: cost-efficiency frontier. Panel B: acceptability curves for different cost-effectiveness thresholds (the dashed line is the cost-effectiveness acceptability frontier).

| Strategy | Expected number of colposcopy referrals per woman over life-time | Mean life-time risk of cancer (%) | Proportion of cancers detected at local stage (%) | Life-years (undis-counted, from age 9 years) | Total cost per woman (€; dis-counted) | QALE (dis-counted) | Incremental cost (€; dis-counted) | Incremental QALEs (dis-counted) | ICER (Cost (€) per QALY gained) | Status |
|---|---|---|---|---|---|---|---|---|---|---|
| No screening | 0.000 | 1.744 | 67.10 | 75.69084 | 145.51 | 25.64434 | | | | ND |
| CC-HPV, 25y, 10-10y | 0.786 | 0.202 | 81.66 | 75.89505 | 515.05 | 25.68114 | 369.53 | 0.03680 | 10040 | ND |
| CC-HPV, 23y, 10-10y | 0.895 | 0.196 | 80.93 | 75.89520 | 579.52 | 25.68143 | | | | ED |
| CC-HPV(s), 25y, 5-10y | 0.739 | 0.198 | 83.74 | 75.89615 | 589.44 | 25.68070 | | | | D |
| CC-HPV, 25y, 7-10y | 0.887 | 0.173 | 83.30 | 75.89943 | 589.51 | 25.68210 | 74.46 | 0.00096 | 77594 | ND |
| CC-HPV, 25y, 7-7y | 0.920 | 0.158 | 84.44 | 75.90032 | 608.81 | 25.68219 | | | | ED |
| CC-HPV(s), 25y, 5-7y | 0.767 | 0.191 | 83.98 | 75.89664 | 609.29 | 25.68076 | | | | D |
| CC-HPV(s), 23y, 5-10y | 0.703 | 0.210 | 83.02 | 75.89514 | 612.87 | 25.68062 | | | | D |
| CC-HPV(s), 23y, 5-7y | 0.730 | 0.200 | 83.71 | 75.89583 | 630.89 | 25.68069 | | | | D |
| CC-HPV, 23y, 7-10y | 1.028 | 0.148 | 83.42 | 75.90099 | 671.00 | 25.68265 | | | | ED |
| CC-HPV, 25y, 5-10y | 1.005 | 0.142 | 84.44 | 75.90247 | 679.91 | 25.68276 | 90.40 | 0.00066 | 136229 | ND |
| CC-HPV, 23y, 7-7y | 1.046 | 0.144 | 83.94 | 75.90159 | 683.65 | 25.68272 | | | | D |
| CC-HPV, 25y, 5-7y | 1.035 | 0.133 | 85.29 | 75.90330 | 698.90 | 25.68286 | | | | ED |
| CC-HPV, 23y, 5-10y | 1.151 | 0.126 | 83.79 | 75.90396 | 771.10 | 25.68337 | 91.19 | 0.00060 | 151147 | ND |
| CC-HPV(s), 25y, 3-7y | 0.903 | 0.156 | 84.71 | 75.90072 | 785.28 | 25.68188 | | | | D |
| CC-HPV, 23y, 5-7y | 1.179 | 0.115 | 85.14 | 75.90480 | 787.93 | 25.68346 | 16.83 | 0.00009 | 180913 | ND |
| CC-HPV(s), 23y, 3-7y | 0.889 | 0.145 | 85.93 | 75.90172 | 831.74 | 25.68224 | | | | D |
| CC-HPV, 25y, 3-7y | 1.214 | 0.117 | 85.66 | 75.90498 | 877.75 | 25.68331 | | | | D |
| CC-HPV, 23y, 3-7y | 1.396 | 0.093 | 85.63 | 75.90667 | 988.78 | 25.68396 | 200.85 | 0.00050 | 398279 | ND |

Table F.1: Summary statistics, costs, quality-adjusted life-years and ICERs on the cost-efficiency frontier, *restricted to clinician-collected sampling*, Sweden. Abbreviations: QALE=Quality-adjusted life expectancy; QALY: Quality-adjusted life-year; ICER=Incremental cost-effectiveness ratio; ND=Not dominated; D=Dominated; ED=Extended dominated.

#### F.2 Base analysis for all strategies

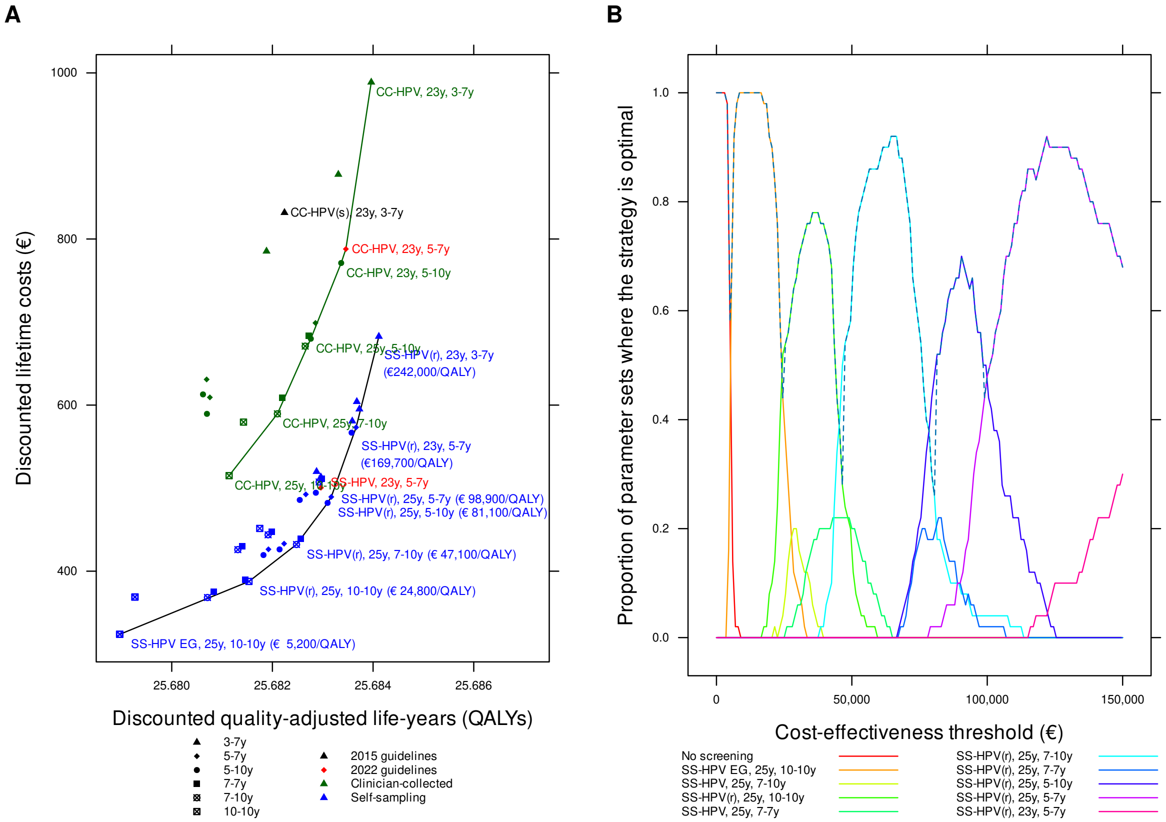

Figure F.2: Cost-effectiveness of cervical cancer screening strategies in unvaccinated cohorts in Sweden: *all strategies*. Panel A: cost-efficiency frontier for clinician-collected tests (green) and all testing (black). Panel B: acceptability curves for different cost-effectiveness thresholds (the dashed line is the cost-effectiveness acceptability frontier).

| Strategy | Expected number of colposcopy referrals per woman over lifetime | Mean lifetime risk of cancer (%) | Proportion of cancers detected at local stage (%) | Life-years (undiscounted, from age 9 years) | Total cost per woman (€; discounted) | QALE (discounted) | Incremental cost (€; discounted) | Incremental QALEs (discounted) | ICER (Cost (€) per QALY gained) | Status |
|---|---|---|---|---|---|---|---|---|---|---|
| No screening | 0.000 | 1.744 | 67.10 | 75.69084 | 145.51 | 25.64434 | | | | ND |
| SS-HPV EG, 25y, 10-10y | 0.626 | 0.277 | 79.46 | 75.88525 | 324.01 | 25.67897 | 178.49 | 0.03463 | 5154 | ND |
| SS-HPV, 25y, 10-10y | 0.626 | 0.278 | 79.44 | 75.88523 | 324.03 | 25.67896 | | | | D |
| SS-HPV EG, 25y, 7-10y | 0.736 | 0.224 | 81.14 | 75.89366 | 368.26 | 25.68071 | | | | ED |
| SS-HPV, 25y, 7-10y | 0.736 | 0.224 | 81.14 | 75.89366 | 368.26 | 25.68071 | | | | D |
| SS-HPV, 23y, 10-10y | 0.710 | 0.274 | 79.08 | 75.88550 | 368.78 | 25.67927 | | | | D |
| SS-HPV EG, 23y, 10-10y | 0.711 | 0.274 | 79.07 | 75.88552 | 368.97 | 25.67927 | | | | D |
| SS-HPV EG, 25y, 7-7y | 0.763 | 0.205 | 82.27 | 75.89489 | 375.25 | 25.68084 | | | | ED |
| SS-HPV(r), 25y, 10-10y | 0.976 | 0.190 | 82.32 | 75.89666 | 387.64 | 25.68153 | 63.63 | 0.00256 | 24809 | ND |
| SS-HPV, 25y, 7-7y | 0.751 | 0.185 | 82.99 | 75.89743 | 389.67 | 25.68146 | | | | D |
| SS-HPV EG, 25y, 5-10y | 0.864 | 0.173 | 83.11 | 75.89866 | 419.41 | 25.68182 | | | | ED |
| SS-HPV EG, 23y, 7-10y | 0.856 | 0.197 | 82.41 | 75.89549 | 425.96 | 25.68131 | | | | D |
| SS-HPV, 25y, 5-10y | 0.824 | 0.163 | 83.28 | 75.89991 | 426.22 | 25.68214 | | | | ED |
| SS-HPV EG, 25y, 5-7y | 0.885 | 0.162 | 83.73 | 75.89952 | 426.34 | 25.68192 | | | | D |
| SS-HPV EG, 23y, 7-7y | 0.867 | 0.192 | 82.75 | 75.89619 | 429.80 | 25.68140 | | | | D |
| SS-HPV(r), 25y, 7-10y | 1.115 | 0.160 | 83.64 | 75.90093 | 432.14 | 25.68248 | 44.50 | 0.00094 | 47104 | ND |
| SS-HPV, 25y, 5-7y | 0.845 | 0.152 | 84.08 | 75.90071 | 433.14 | 25.68223 | | | | D |
| SS-HPV(r), 25y, 7-7y | 1.153 | 0.146 | 84.91 | 75.90172 | 439.16 | 25.68256 | | | | ED |
| SS-HPV, 23y, 7-10y | 0.849 | 0.174 | 82.75 | 75.89785 | 443.82 | 25.68191 | | | | D |
| SS-HPV, 23y, 7-7y | 0.858 | 0.170 | 82.89 | 75.89845 | 447.70 | 25.68199 | | | | D |
| SS-HPV(r), 23y, 10-10y | 1.124 | 0.183 | 81.41 | 75.89652 | 451.45 | 25.68175 | | | | D |
| SS-HPV(r), 25y, 5-10y | 1.277 | 0.127 | 84.87 | 75.90375 | 482.20 | 25.68310 | 50.07 | 0.00062 | 81087 | ND |
| SS-HPV EG, 23y, 5-10y | 0.992 | 0.162 | 82.52 | 75.90044 | 485.76 | 25.68254 | | | | D |
| SS-HPV(r), 25y, 5-7y | 1.313 | 0.120 | 85.54 | 75.90440 | 489.42 | 25.68317 | 7.22 | 0.00007 | 98938 | ND |
| SS-HPV EG, 23y, 5-7y | 1.017 | 0.144 | 84.10 | 75.90159 | 492.49 | 25.68266 | | | | D |
| SS-HPV, 23y, 5-10y | 0.950 | 0.149 | 82.81 | 75.90184 | 494.44 | 25.68286 | | | | D |
| SS-HPV, 23y, 5-7y | 0.970 | 0.135 | 84.39 | 75.90278 | 500.49 | 25.68296 | | | | D |
| SS-HPV(r), 23y, 7-10y | 1.304 | 0.135 | 84.62 | 75.90205 | 507.39 | 25.68293 | | | | D |
| SS-HPV(r), 23y, 7-7y | 1.321 | 0.133 | 84.76 | 75.90252 | 511.46 | 25.68298 | | | | D |
| SS-HPV, 25y, 3-7y | 0.989 | 0.131 | 84.60 | 75.90384 | 513.30 | 25.68296 | | | | D |
| CC-HPV, 25y, 10-10y | 0.786 | 0.202 | 81.66 | 75.89505 | 515.05 | 25.68114 | | | | D |
| SS-HPV EG, 25y, 3-7y | 1.086 | 0.134 | 84.55 | 75.90349 | 519.90 | 25.68288 | | | | D |
| SS-HPV(r), 23y, 5-10y | 1.476 | 0.118 | 84.34 | 75.90464 | 566.76 | 25.68357 | | | | ED |
| SS-HPV(r), 23y, 5-7y | 1.510 | 0.106 | 86.31 | 75.90550 | 573.15 | 25.68366 | 83.73 | 0.00049 | 169740 | ND |
| CC-HPV, 23y, 10-10y | 0.895 | 0.196 | 80.93 | 75.89520 | 579.52 | 25.68143 | | | | D |
| SS-HPV(r), 25y, 3-7y | 1.566 | 0.106 | 86.02 | 75.90596 | 580.71 | 25.68359 | | | | D |
| CC-HPV(s), 25y, 5-10y | 0.739 | 0.198 | 83.74 | 75.89615 | 589.44 | 25.68070 | | | | D |
| CC-HPV, 25y, 7-10y | 0.887 | 0.173 | 83.30 | 75.89943 | 589.51 | 25.68210 | | | | D |
| SS-HPV, 23y, 3-7y | 1.147 | 0.103 | 85.18 | 75.90598 | 595.19 | 25.68373 | | | | ED |

Continued on next page

| Strategy | Expected number of colposcopy referrals per woman over life-time | Mean life-time risk of cancer (%) | Proportion of cancers detected at local stage (%) | Life-years (undis-counted, from age 9 years) | Total cost per woman (€; dis-counted) | QALE (dis-counted) | Incremental cost (€; dis-counted) | Incremental QALEs (dis-counted) | ICER (Cost (€) per QALY gained) | Status |
|---|---|---|---|---|---|---|---|---|---|---|
| SS-HPV EG, 23y, 3-7y | 1.256 | 0.105 | 85.10 | 75.90575 | 604.11 | 25.68368 | | | | D |
| CC-HPV, 25y, 7-7y | 0.920 | 0.158 | 84.44 | 75.90032 | 608.81 | 25.68219 | | | | D |
| CC-HPV(s), 25y, 5-7y | 0.767 | 0.191 | 83.98 | 75.89664 | 609.29 | 25.68076 | | | | D |
| CC-HPV(s), 23y, 5-10y | 0.703 | 0.210 | 83.02 | 75.89514 | 612.87 | 25.68062 | | | | D |
| CC-HPV(s), 23y, 5-7y | 0.730 | 0.200 | 83.71 | 75.89583 | 630.89 | 25.68069 | | | | D |
| CC-HPV, 23y, 7-10y | 1.028 | 0.148 | 83.42 | 75.90099 | 671.00 | 25.68265 | | | | D |
| CC-HPV, 25y, 5-10y | 1.005 | 0.142 | 84.44 | 75.90247 | 679.91 | 25.68276 | | | | D |
| SS-HPV(r), 23y, 3-7y | 1.820 | 0.085 | 86.51 | 75.90727 | 682.63 | 25.68411 | 109.49 | 0.00045 | 242014 | ND |
| CC-HPV, 23y, 7-7y | 1.046 | 0.144 | 83.94 | 75.90159 | 683.65 | 25.68272 | | | | D |
| CC-HPV, 25y, 5-7y | 1.035 | 0.133 | 85.29 | 75.90330 | 698.90 | 25.68286 | | | | D |
| CC-HPV, 23y, 5-10y | 1.151 | 0.126 | 83.79 | 75.90396 | 771.10 | 25.68337 | | | | D |
| CC-HPV(s), 25y, 3-7y | 0.903 | 0.156 | 84.71 | 75.90072 | 785.28 | 25.68188 | | | | D |
| CC-HPV, 23y, 5-7y | 1.179 | 0.115 | 85.14 | 75.90480 | 787.93 | 25.68346 | | | | D |
| CC-HPV(s), 23y, 3-7y | 0.889 | 0.145 | 85.93 | 75.90172 | 831.74 | 25.68224 | | | | D |
| CC-HPV, 25y, 3-7y | 1.214 | 0.117 | 85.66 | 75.90498 | 877.75 | 25.68331 | | | | D |
| CC-HPV, 23y, 3-7y | 1.396 | 0.093 | 85.63 | 75.90667 | 988.78 | 25.68396 | | | | D |

Table F.2: Summary statistics, costs, quality-adjusted life-years and ICERs on the cost-efficiency frontier, *base analysis*, Sweden. Abbreviations: QALE=Quality-adjusted life expectancy; QALY: Quality-adjusted life-year; ICER=Incremental cost-effectiveness ratio; ND=Not dominated; D=Dominated; ED=Extended dominated.

### G Sensitivity analyses restricted to clinician-collected sampling

#### G.1 Direct medical costs

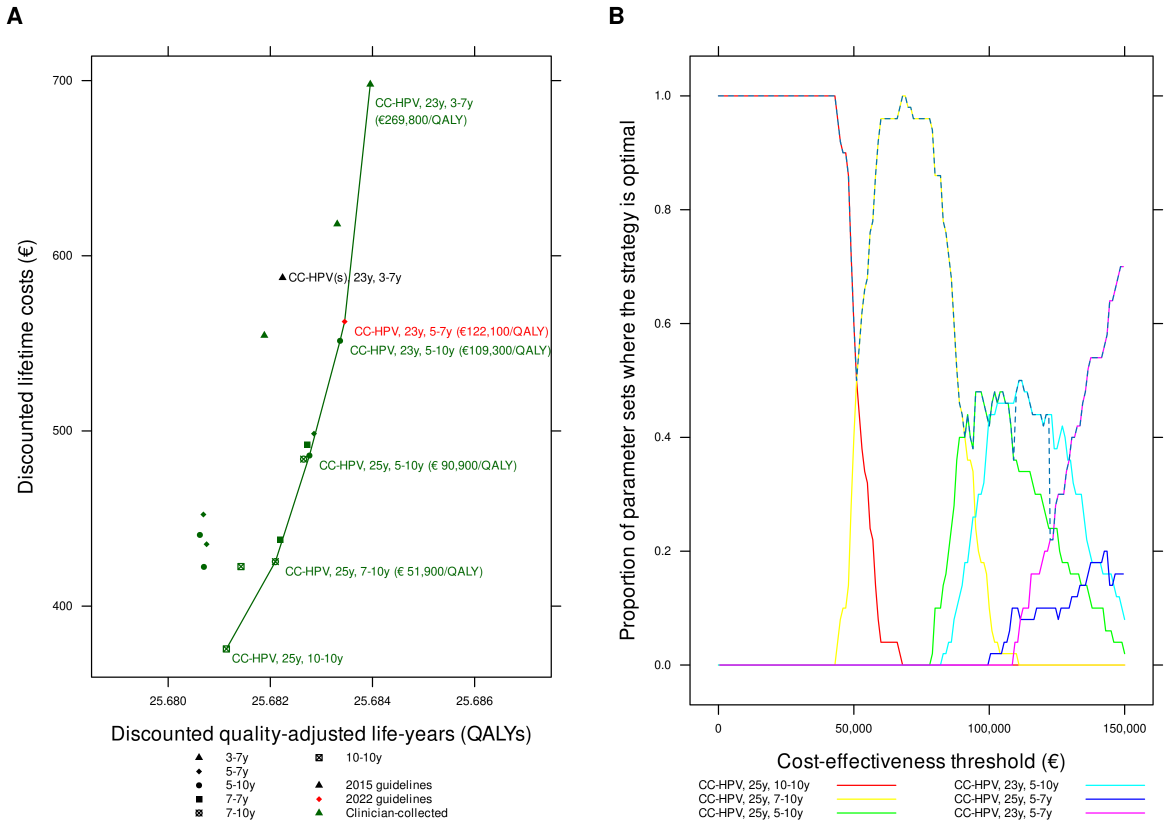

Figure G.1: Cost-effectiveness of cervical cancer screening strategies in unvaccinated cohorts in Sweden: *restricted to clinician-collected sampling, direct medical costs*. Panel A: cost-efficiency frontier. Panel B: acceptability curves for different cost-effectiveness thresholds (the dashed line is the cost-effectiveness acceptability frontier).

| Strategy | Expected number of colposcopy referrals per woman over lifetime | Mean lifetime risk of cancer (%) | Proportion of cancers detected at local stage (%) | Life-years (undiscounted, from age 9 years) | Total cost per woman (€; discounted) | QALE (discounted) | Incremental cost (€; discounted) | Incremental QALEs (discounted) | ICER (Cost (€) per QALY gained) | Status |
|---|---|---|---|---|---|---|---|---|---|---|
| CC-HPV, 25y, 10-10y | 0.786 | 0.202 | 81.66 | 75.89505 | 375.56 | 25.68114 | | | | ND |
| CC-HPV(s), 25y, 5-10y | 0.739 | 0.198 | 83.74 | 75.89615 | 422.40 | 25.68070 | | | | D |
| CC-HPV, 23y, 10-10y | 0.895 | 0.196 | 80.93 | 75.89520 | 422.58 | 25.68143 | | | | ED |
| CC-HPV, 25y, 7-10y | 0.887 | 0.173 | 83.30 | 75.89943 | 425.40 | 25.68210 | 49.84 | 0.00096 | 51941 | ND |
| CC-HPV(s), 25y, 5-7y | 0.766 | 0.191 | 83.98 | 75.89662 | 435.34 | 25.68075 | | | | D |
| CC-HPV, 25y, 7-7y | 0.920 | 0.158 | 84.44 | 75.90032 | 437.98 | 25.68219 | | | | ED |
| CC-HPV(s), 23y, 5-10y | 0.703 | 0.210 | 83.02 | 75.89514 | 440.64 | 25.68062 | | | | D |
| CC-HPV(s), 23y, 5-7y | 0.729 | 0.200 | 83.71 | 75.89581 | 452.31 | 25.68069 | | | | D |
| CC-HPV, 23y, 7-10y | 1.028 | 0.148 | 83.42 | 75.90099 | 483.93 | 25.68265 | | | | ED |
| CC-HPV, 25y, 5-10y | 1.005 | 0.142 | 84.45 | 75.90248 | 485.96 | 25.68277 | 60.56 | 0.00067 | 90880 | ND |
| CC-HPV, 23y, 7-7y | 1.046 | 0.144 | 83.94 | 75.90159 | 492.23 | 25.68272 | | | | D |
| CC-HPV, 25y, 5-7y | 1.035 | 0.133 | 85.29 | 75.90330 | 498.45 | 25.68286 | | | | ED |
| CC-HPV, 23y, 5-10y | 1.151 | 0.126 | 83.78 | 75.90395 | 551.45 | 25.68337 | 65.49 | 0.00060 | 109274 | ND |
| CC-HPV(s), 25y, 3-7y | 0.903 | 0.156 | 84.71 | 75.90072 | 554.52 | 25.68188 | | | | D |
| CC-HPV, 23y, 5-7y | 1.179 | 0.115 | 85.15 | 75.90479 | 562.43 | 25.68346 | 10.98 | 0.00009 | 122064 | ND |
| CC-HPV(s), 23y, 3-7y | 0.889 | 0.145 | 85.93 | 75.90172 | 587.48 | 25.68224 | | | | D |
| CC-HPV, 25y, 3-7y | 1.214 | 0.117 | 85.65 | 75.90497 | 618.12 | 25.68331 | | | | D |
| CC-HPV, 23y, 3-7y | 1.397 | 0.093 | 85.64 | 75.90666 | 697.89 | 25.68396 | 135.46 | 0.00050 | 269805 | ND |

Table G.1: Summary statistics, costs, quality-adjusted life-years and ICERs on the cost-efficiency frontier, *restricted to clinician-collected sampling, direct medical costs only*, Sweden. Abbreviations: QALE=Quality-adjusted life expectancy; QALY: Quality-adjusted life-year; ICER=Incremental cost-effectiveness ratio; ND=Not dominated; D=Dominated; ED=Extended dominated.

#### G.2 Assumed lower compliance

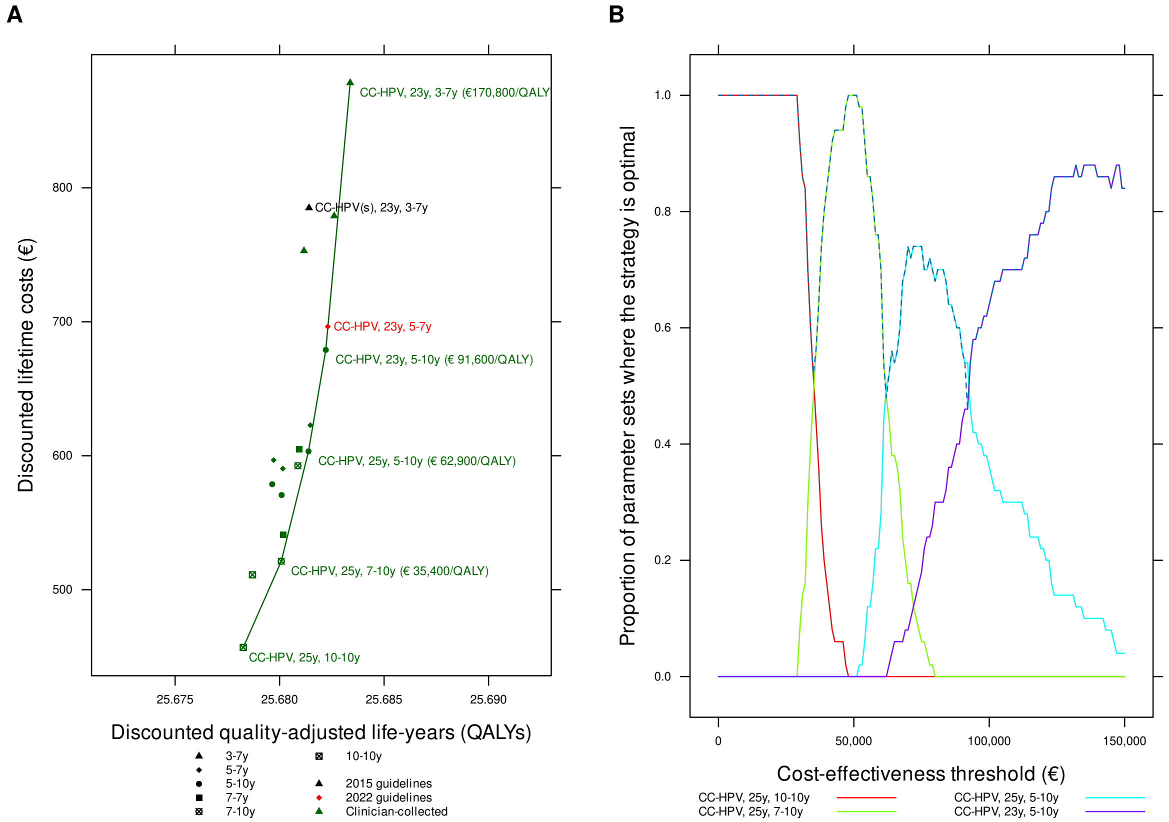

Figure G.2: Cost-effectiveness of cervical cancer screening strategies in unvaccinated cohorts in Sweden: *restricted to clinician-collected sampling, assumed lower compliance.* Panel A: cost-efficiency frontier. Panel B: acceptability curves for different cost-effectiveness thresholds (the dashed line is the cost-effectiveness acceptability frontier).

| Strategy | Expected number of colposcopy referrals per woman over life-time | Mean life-time risk of cancer (%) | Proportion of cancers detected at local stage (%) | Life-years (undis-counted, from age 9 years) | Total cost per woman (€; dis-counted) | QALE (dis-counted) | Incremental cost (€; dis-counted) | Incremental QALEs (dis-counted) | ICER (Cost (€) per QALY gained) | Status |
|---|---|---|---|---|---|---|---|---|---|---|
| CC-HPV, 25y, 10-10y | 0.677 | 0.294 | 79.00 | 75.88249 | 456.87 | 25.67826 | | | | ND |
| CC-HPV, 23y, 10-10y | 0.768 | 0.283 | 78.54 | 75.88339 | 511.13 | 25.67870 | | | | ED |
| CC-HPV, 25y, 7-10y | 0.770 | 0.237 | 81.13 | 75.89125 | 521.25 | 25.68008 | 64.38 | 0.00182 | 35405 | ND |
| CC-HPV, 25y, 7-7y | 0.803 | 0.222 | 81.79 | 75.89217 | 540.91 | 25.68018 | | | | ED |
| CC-HPV(s), 25y, 5-10y | 0.718 | 0.214 | 83.13 | 75.89420 | 570.67 | 25.68010 | | | | D |
| CC-HPV(s), 23y, 5-10y | 0.670 | 0.237 | 82.14 | 75.89190 | 578.74 | 25.67964 | | | | D |
| CC-HPV(s), 25y, 5-7y | 0.745 | 0.207 | 83.31 | 75.89470 | 590.40 | 25.68015 | | | | D |
| CC-HPV, 23y, 7-10y | 0.894 | 0.206 | 81.43 | 75.89402 | 592.59 | 25.68087 | | | | ED |
| CC-HPV(s), 23y, 5-7y | 0.695 | 0.227 | 82.73 | 75.89259 | 596.73 | 25.67971 | | | | D |
| CC-HPV, 25y, 5-10y | 0.884 | 0.185 | 82.61 | 75.89730 | 603.28 | 25.68138 | 82.03 | 0.00130 | 62940 | ND |
| CC-HPV, 23y, 7-7y | 0.910 | 0.203 | 81.71 | 75.89459 | 604.74 | 25.68093 | | | | D |
| CC-HPV, 25y, 5-7y | 0.915 | 0.176 | 83.08 | 75.89808 | 622.80 | 25.68147 | | | | ED |
| CC-HPV, 23y, 5-10y | 1.005 | 0.167 | 82.41 | 75.89974 | 679.02 | 25.68221 | 75.75 | 0.00083 | 91557 | ND |
| CC-HPV, 23y, 5-7y | 1.034 | 0.155 | 83.52 | 75.90064 | 696.38 | 25.68230 | | | | ED |
| CC-HPV(s), 25y, 3-7y | 0.869 | 0.177 | 83.99 | 75.89844 | 752.97 | 25.68116 | | | | D |
| CC-HPV, 25y, 3-7y | 1.081 | 0.140 | 84.65 | 75.90270 | 779.01 | 25.68261 | | | | ED |
| CC-HPV(s), 23y, 3-7y | 0.849 | 0.170 | 85.21 | 75.89911 | 785.01 | 25.68141 | | | | D |
| CC-HPV, 23y, 3-7y | 1.245 | 0.114 | 85.05 | 75.90474 | 878.36 | 25.68338 | 199.33 | 0.00117 | 170771 | ND |

Table G.2: Summary statistics, costs, quality-adjusted life-years and ICERs on the cost-efficiency frontier, *restricted to clinician-collected sampling, assumed lower compliance*, Sweden. Abbreviations: QALE=Quality-adjusted life expectancy; QALY: Quality-adjusted life-year; ICER=Incremental cost-effectiveness ratio; ND=Not dominated; D=Dominated; ED=Extended dominated.

#### G.3 Undiscounted QALYs and costs

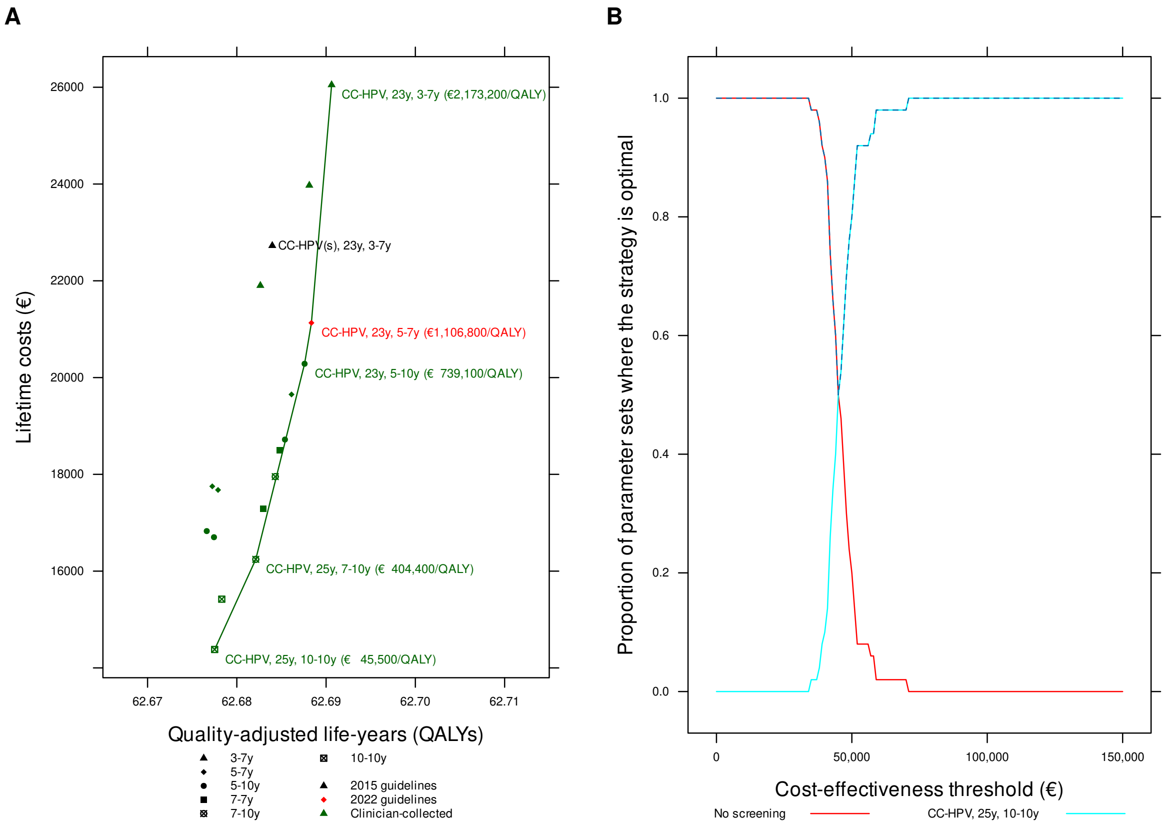

Figure G.3: Cost-effectiveness of cervical cancer screening strategies in unvaccinated cohorts in Sweden: *restricted to clinician-collected sampling, undiscounted.* Panel A: cost-efficiency frontier. Panel B: acceptability curves for different cost-effectiveness thresholds.

| Strategy | Expected number of colposcopy referrals per woman over life-time | Mean life-time risk of cancer (%) | Proportion of cancers detected at local stage (%) | Life-years (undis-counted, from age 9 years) | Total cost per woman (€; undis-counted) | QALE (undis-counted) | Incremental cost (€; undis-counted) | Incremental QALEs (undis-counted) | ICER (Cost (€) per QALY gained) | Status |
|---|---|---|---|---|---|---|---|---|---|---|
| No screening | 0.000 | 1.744 | 67.10 | 75.69084 | 5241.66 | 62.47672 | | | | ND |
| CC-HPV, 25y, 10-10y | 0.786 | 0.202 | 81.66 | 75.89505 | 14380.39 | 62.67752 | 9138.73 | 0.20080 | 45511 | ND |
| CC-HPV, 23y, 10-10y | 0.895 | 0.196 | 80.93 | 75.89520 | 15420.71 | 62.67831 | | | | ED |
| CC-HPV, 25y, 7-10y | 0.887 | 0.173 | 83.30 | 75.89943 | 16243.68 | 62.68212 | 1863.29 | 0.00461 | 404449 | ND |
| CC-HPV(s), 25y, 5-10y | 0.739 | 0.198 | 83.74 | 75.89615 | 16700.90 | 62.67744 | | | | D |
| CC-HPV(s), 23y, 5-10y | 0.703 | 0.210 | 83.02 | 75.89514 | 16827.49 | 62.67663 | | | | D |
| CC-HPV, 25y, 7-7y | 0.920 | 0.158 | 84.44 | 75.90032 | 17287.44 | 62.68294 | | | | ED |
| CC-HPV(s), 25y, 5-7y | 0.767 | 0.191 | 83.98 | 75.89664 | 17677.42 | 62.67790 | | | | D |
| CC-HPV(s), 23y, 5-7y | 0.730 | 0.200 | 83.71 | 75.89583 | 17752.43 | 62.67725 | | | | D |
| CC-HPV, 23y, 7-10y | 1.028 | 0.148 | 83.42 | 75.90099 | 17953.46 | 62.68431 | | | | ED |
| CC-HPV, 23y, 7-7y | 1.046 | 0.144 | 83.94 | 75.90159 | 18493.44 | 62.68483 | | | | ED |
| CC-HPV, 25y, 5-10y | 1.005 | 0.142 | 84.44 | 75.90247 | 18719.51 | 62.68539 | | | | ED |
| CC-HPV, 25y, 5-7y | 1.035 | 0.133 | 85.29 | 75.90330 | 19651.39 | 62.68613 | | | | ED |
| CC-HPV, 23y, 5-10y | 1.151 | 0.126 | 83.79 | 75.90396 | 20285.13 | 62.68759 | 4041.46 | 0.00547 | 739057 | ND |
| CC-HPV, 23y, 5-7y | 1.179 | 0.115 | 85.14 | 75.90480 | 21130.59 | 62.68836 | 845.46 | 0.00076 | 1106756 | ND |
| CC-HPV(s), 25y, 3-7y | 0.903 | 0.156 | 84.71 | 75.90072 | 21901.96 | 62.68264 | | | | D |
| CC-HPV(s), 23y, 3-7y | 0.889 | 0.145 | 85.93 | 75.90172 | 22726.39 | 62.68397 | | | | D |
| CC-HPV, 25y, 3-7y | 1.214 | 0.117 | 85.66 | 75.90498 | 23972.60 | 62.68812 | | | | D |
| CC-HPV, 23y, 3-7y | 1.396 | 0.093 | 85.63 | 75.90667 | 26049.52 | 62.69062 | 4918.92 | 0.00226 | 2173247 | ND |

Table G.3: Summary statistics, costs, quality-adjusted life-years and ICERs on the cost-efficiency frontier, *restricted to clinician-collected sampling, undiscounted*, Sweden. Abbreviations: QALE=Quality-adjusted life expectancy; QALY: Quality-adjusted life-year; ICER=Incremental cost-effectiveness ratio; ND=Not dominated; D=Dominated; ED=Extended dominated.

#### G.4 5% discounted QALYs and costs

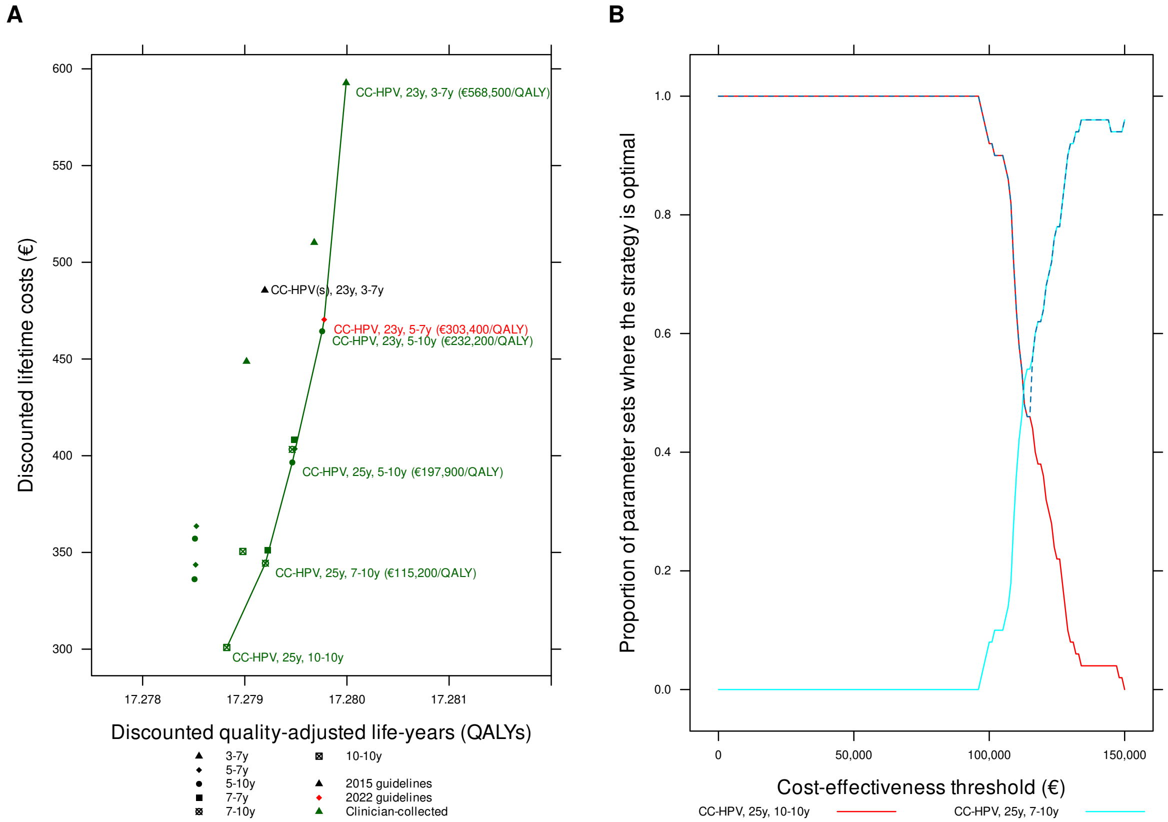

Figure G.4: Cost-effectiveness of cervical cancer screening strategies in unvaccinated cohorts in Sweden: *restricted to clinician-collected sampling, 5% discount rate for QALYs and costs.* Panel A: cost-efficiency frontier. Panel B: acceptability curves for different cost-effectiveness thresholds (the dashed line is the cost-effectiveness acceptability frontier).

| Strategy | Expected number of colposcopy referrals per woman over lifetime | Mean lifetime risk of cancer (%) | Proportion of cancers detected at local stage (%) | Life-years (undiscounted, from age 9 years) | Total cost per woman (€; discounted) | QALE (discounted) | Incremental cost (€; discounted) | Incremental QALEs (discounted) | ICER (Cost (€) per QALY gained) | Status |
|---|---|---|---|---|---|---|---|---|---|---|
| CC-HPV, 25y, 10-10y | 0.786 | 0.202 | 81.66 | 75.89505 | 300.86 | 17.27882 | | | | ND |
| CC-HPV(s), 25y, 5-10y | 0.739 | 0.198 | 83.74 | 75.89615 | 336.15 | 17.27851 | | | | D |
| CC-HPV(s), 25y, 5-7y | 0.767 | 0.191 | 83.97 | 75.89660 | 343.61 | 17.27852 | | | | D |
| CC-HPV, 25y, 7-10y | 0.887 | 0.173 | 83.30 | 75.89943 | 344.43 | 17.27920 | 43.57 | 0.00038 | 115180 | ND |
| CC-HPV, 23y, 10-10y | 0.895 | 0.195 | 80.94 | 75.89521 | 350.50 | 17.27898 | | | | D |
| CC-HPV, 25y, 7-7y | 0.920 | 0.158 | 84.43 | 75.90032 | 351.12 | 17.27922 | | | | ED |
| CC-HPV(s), 23y, 5-10y | 0.703 | 0.210 | 83.03 | 75.89516 | 357.14 | 17.27851 | | | | D |
| CC-HPV(s), 23y, 5-7y | 0.729 | 0.200 | 83.71 | 75.89581 | 363.59 | 17.27853 | | | | D |
| CC-HPV, 25y, 5-10y | 1.005 | 0.142 | 84.44 | 75.90248 | 396.53 | 17.27947 | 52.10 | 0.00026 | 197883 | ND |
| CC-HPV, 23y, 7-10y | 1.028 | 0.148 | 83.42 | 75.90099 | 403.25 | 17.27946 | | | | D |
| CC-HPV, 25y, 5-7y | 1.035 | 0.133 | 85.29 | 75.90330 | 403.54 | 17.27949 | | | | ED |
| CC-HPV, 23y, 7-7y | 1.046 | 0.144 | 83.94 | 75.90159 | 408.27 | 17.27948 | | | | D |
| CC-HPV(s), 25y, 3-7y | 0.903 | 0.156 | 84.71 | 75.90072 | 448.73 | 17.27902 | | | | D |
| CC-HPV, 23y, 5-10y | 1.151 | 0.126 | 83.79 | 75.90396 | 464.34 | 17.27976 | 67.81 | 0.00029 | 232209 | ND |
| CC-HPV, 23y, 5-7y | 1.178 | 0.115 | 85.14 | 75.90477 | 470.37 | 17.27978 | 6.03 | 0.00002 | 303444 | ND |
| CC-HPV(s), 23y, 3-7y | 0.889 | 0.145 | 85.93 | 75.90172 | 485.52 | 17.27920 | | | | D |
| CC-HPV, 25y, 3-7y | 1.214 | 0.117 | 85.66 | 75.90498 | 510.20 | 17.27968 | | | | D |
| CC-HPV, 23y, 3-7y | 1.397 | 0.093 | 85.61 | 75.90667 | 592.76 | 17.27999 | 122.39 | 0.00022 | 568506 | ND |

Table G.4: Summary statistics, costs, quality-adjusted life-years and ICERs on the cost-efficiency frontier, *restricted to clinician-collected sampling, 5% discount rate for QALYs and costs*, Sweden. Abbreviations: QALE=Quality-adjusted life expectancy; QALY: Quality-adjusted life-year; ICER=Incremental cost-effectiveness ratio; ND=Not dominated; D=Dominated; ED=Extended dominated.

#### G.5 50% lower costs

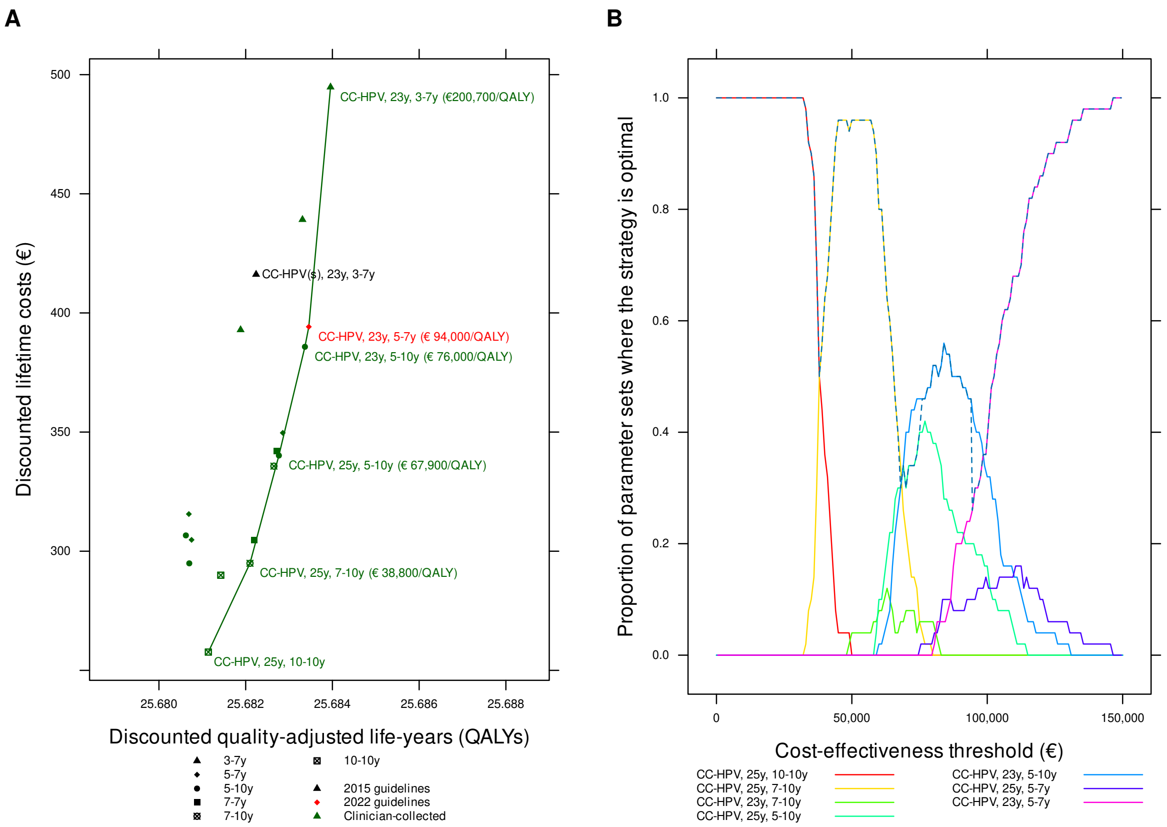

Figure G.5: Cost-effectiveness of cervical cancer screening strategies in unvaccinated cohorts in Sweden: *restricted to clinician-collected sampling, 50% lower costs*. Panel A: cost-efficiency frontier. Panel B: acceptability curves for different cost-effectiveness thresholds (the dashed line is the cost-effectiveness acceptability frontier).

| Strategy | Expected number of colposcopy referrals per woman over lifetime | Mean lifetime risk of cancer (%) | Proportion of cancers detected at local stage (%) | Life-years (undiscounted, from age 9 years) | Total cost per woman (€; discounted) | QALE (discounted) | Incremental cost (€; discounted) | Incremental QALEs (discounted) | ICER (Cost (€) per QALY gained) | Status |
|---|---|---|---|---|---|---|---|---|---|---|
| CC-HPV, 25y, 10-10y | 0.786 | 0.202 | 81.66 | 75.89505 | 257.67 | 25.68114 | | | | ND |
| CC-HPV, 23y, 10-10y | 0.895 | 0.196 | 80.93 | 75.89520 | 289.93 | 25.68143 | | | | ED |
| CC-HPV(s), 25y, 5-10y | 0.739 | 0.198 | 83.74 | 75.89615 | 294.91 | 25.68070 | | | | D |
| CC-HPV, 25y, 7-10y | 0.887 | 0.173 | 83.30 | 75.89943 | 294.93 | 25.68210 | 37.26 | 0.00096 | 38825 | ND |
| CC-HPV, 25y, 7-7y | 0.920 | 0.158 | 84.44 | 75.90032 | 304.59 | 25.68219 | | | | ED |
| CC-HPV(s), 25y, 5-7y | 0.766 | 0.191 | 83.98 | 75.89662 | 304.77 | 25.68075 | | | | D |
| CC-HPV(s), 23y, 5-10y | 0.703 | 0.210 | 83.02 | 75.89514 | 306.64 | 25.68062 | | | | D |
| CC-HPV(s), 23y, 5-7y | 0.729 | 0.200 | 83.71 | 75.89581 | 315.60 | 25.68069 | | | | D |
| CC-HPV, 23y, 7-10y | 1.028 | 0.148 | 83.42 | 75.90099 | 335.70 | 25.68265 | | | | ED |
| CC-HPV, 25y, 5-10y | 1.005 | 0.142 | 84.45 | 75.90248 | 340.16 | 25.68277 | 45.22 | 0.00067 | 67868 | ND |
| CC-HPV, 23y, 7-7y | 1.046 | 0.144 | 83.94 | 75.90159 | 342.03 | 25.68272 | | | | D |
| CC-HPV, 25y, 5-7y | 1.035 | 0.133 | 85.29 | 75.90330 | 349.67 | 25.68286 | | | | ED |
| CC-HPV, 23y, 5-10y | 1.151 | 0.126 | 83.79 | 75.90396 | 385.79 | 25.68337 | 45.63 | 0.00060 | 75985 | ND |
| CC-HPV(s), 25y, 3-7y | 0.903 | 0.156 | 84.71 | 75.90072 | 392.91 | 25.68188 | | | | D |
| CC-HPV, 23y, 5-7y | 1.179 | 0.115 | 85.15 | 75.90480 | 394.19 | 25.68346 | 8.40 | 0.00009 | 94015 | ND |
| CC-HPV(s), 23y, 3-7y | 0.889 | 0.145 | 85.93 | 75.90172 | 416.16 | 25.68224 | | | | D |
| CC-HPV, 25y, 3-7y | 1.214 | 0.117 | 85.66 | 75.90498 | 439.16 | 25.68331 | | | | D |
| CC-HPV, 23y, 3-7y | 1.397 | 0.093 | 85.63 | 75.90666 | 494.78 | 25.68396 | 100.59 | 0.00050 | 200729 | ND |

Table G.5: Summary statistics, costs, quality-adjusted life-years and ICERs on the cost-efficiency frontier, *restricted to clinician-collected sampling, 50% lower costs*, Sweden. Abbreviations: QALE=Quality-adjusted life expectancy; QALY: Quality-adjusted life-year; ICER=Incremental cost-effectiveness ratio; ND=Not dominated; D=Dominated; ED=Extended dominated.

#### G.6 50% higher costs

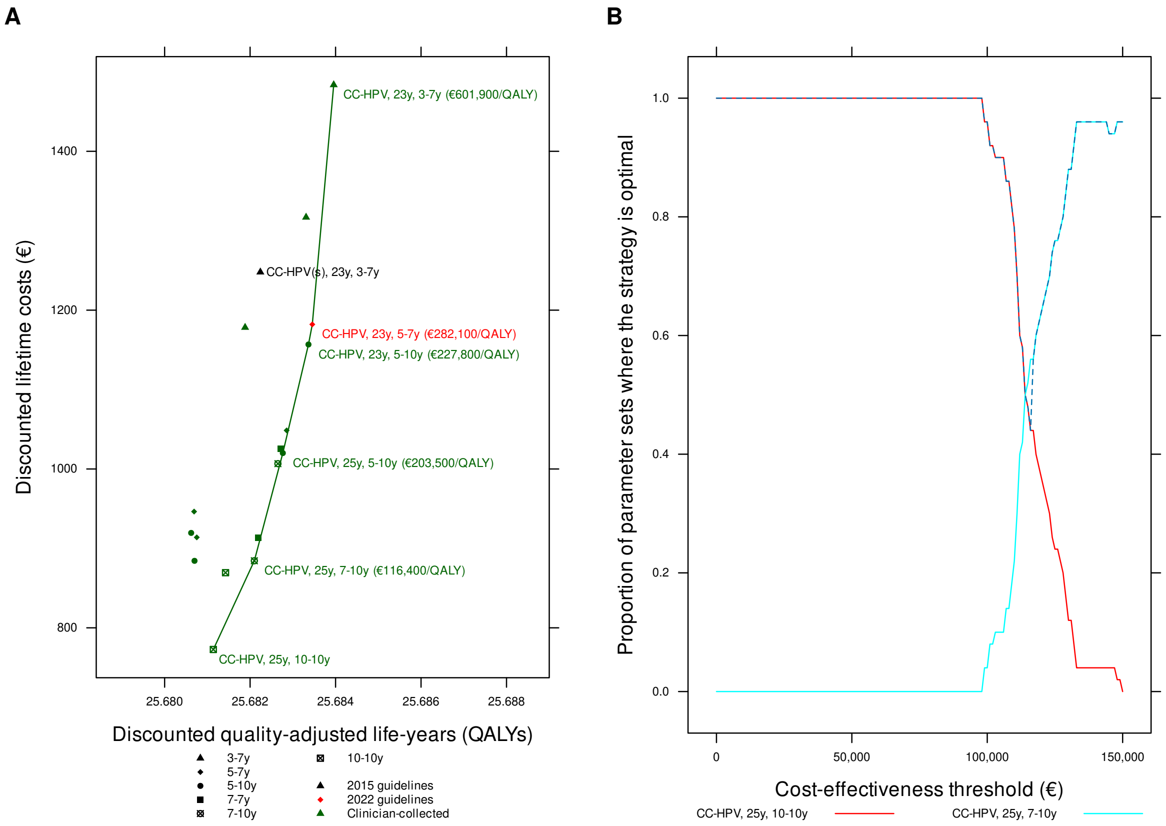

Figure G.6: Cost-effectiveness of cervical cancer screening strategies in unvaccinated cohorts in Sweden: *restricted to clinician-collected sampling, 50% higher costs*. Panel A: cost-efficiency frontier. Panel B: acceptability curves for different cost-effectiveness thresholds (the dashed line is the cost-effectiveness acceptability frontier).

| Strategy | Expected number of colposcopy referrals per woman over lifetime | Mean lifetime risk of cancer (%) | Proportion of cancers detected at local stage (%) | Life-years (undiscounted, from age 9 years) | Total cost per woman (€; discounted) | QALE (discounted) | Incremental cost (€; discounted) | Incremental QALEs (discounted) | ICER (Cost (€) per QALY gained) | Status |
|---|---|---|---|---|---|---|---|---|---|---|
| CC-HPV, 25y, 10-10y | 0.786 | 0.202 | 81.66 | 75.89505 | 772.72 | 25.68114 | | | | ND |
| CC-HPV, 23y, 10-10y | 0.895 | 0.196 | 80.93 | 75.89520 | 869.45 | 25.68143 | | | | ED |
| CC-HPV(s), 25y, 5-10y | 0.739 | 0.198 | 83.74 | 75.89615 | 884.35 | 25.68070 | | | | D |
| CC-HPV, 25y, 7-10y | 0.887 | 0.173 | 83.30 | 75.89943 | 884.44 | 25.68210 | 111.72 | 0.00096 | 116419 | ND |
| CC-HPV, 25y, 7-7y | 0.920 | 0.158 | 84.44 | 75.90032 | 913.39 | 25.68219 | | | | ED |
| CC-HPV(s), 25y, 5-7y | 0.766 | 0.191 | 83.98 | 75.89662 | 913.91 | 25.68075 | | | | D |
| CC-HPV(s), 23y, 5-10y | 0.703 | 0.210 | 83.02 | 75.89514 | 919.51 | 25.68062 | | | | D |
| CC-HPV(s), 23y, 5-7y | 0.729 | 0.200 | 83.71 | 75.89581 | 946.38 | 25.68069 | | | | D |
| CC-HPV, 23y, 7-10y | 1.028 | 0.148 | 83.42 | 75.90099 | 1006.70 | 25.68265 | | | | ED |
| CC-HPV, 25y, 5-10y | 1.005 | 0.142 | 84.45 | 75.90248 | 1020.04 | 25.68277 | 135.60 | 0.00067 | 203500 | ND |
| CC-HPV, 23y, 7-7y | 1.046 | 0.144 | 83.94 | 75.90159 | 1025.68 | 25.68272 | | | | D |
| CC-HPV, 25y, 5-7y | 1.035 | 0.133 | 85.29 | 75.90330 | 1048.57 | 25.68286 | | | | ED |
| CC-HPV, 23y, 5-10y | 1.151 | 0.126 | 83.78 | 75.90396 | 1156.78 | 25.68337 | 136.73 | 0.00060 | 227808 | ND |
| CC-HPV(s), 25y, 3-7y | 0.903 | 0.156 | 84.71 | 75.90072 | 1178.19 | 25.68188 | | | | D |
| CC-HPV, 23y, 5-7y | 1.179 | 0.115 | 85.15 | 75.90480 | 1182.07 | 25.68346 | 25.29 | 0.00009 | 282068 | ND |
| CC-HPV(s), 23y, 3-7y | 0.889 | 0.145 | 85.93 | 75.90172 | 1247.89 | 25.68224 | | | | D |
| CC-HPV, 25y, 3-7y | 1.214 | 0.117 | 85.66 | 75.90498 | 1316.91 | 25.68331 | | | | D |
| CC-HPV, 23y, 3-7y | 1.397 | 0.093 | 85.63 | 75.90666 | 1483.68 | 25.68396 | 301.61 | 0.00050 | 601882 | ND |

Table G.6: Summary statistics, costs, quality-adjusted life-years and ICERs on the cost-efficiency frontier, *restricted to clinician-collected sampling, 50% higher costs*, Sweden. Abbreviations: QALE=Quality-adjusted life expectancy; QALY: Quality-adjusted life-year; ICER=Incremental cost-effectiveness ratio; ND=Not dominated; D=Dominated; ED=Extended dominated.

#### G.7 Life expectancy

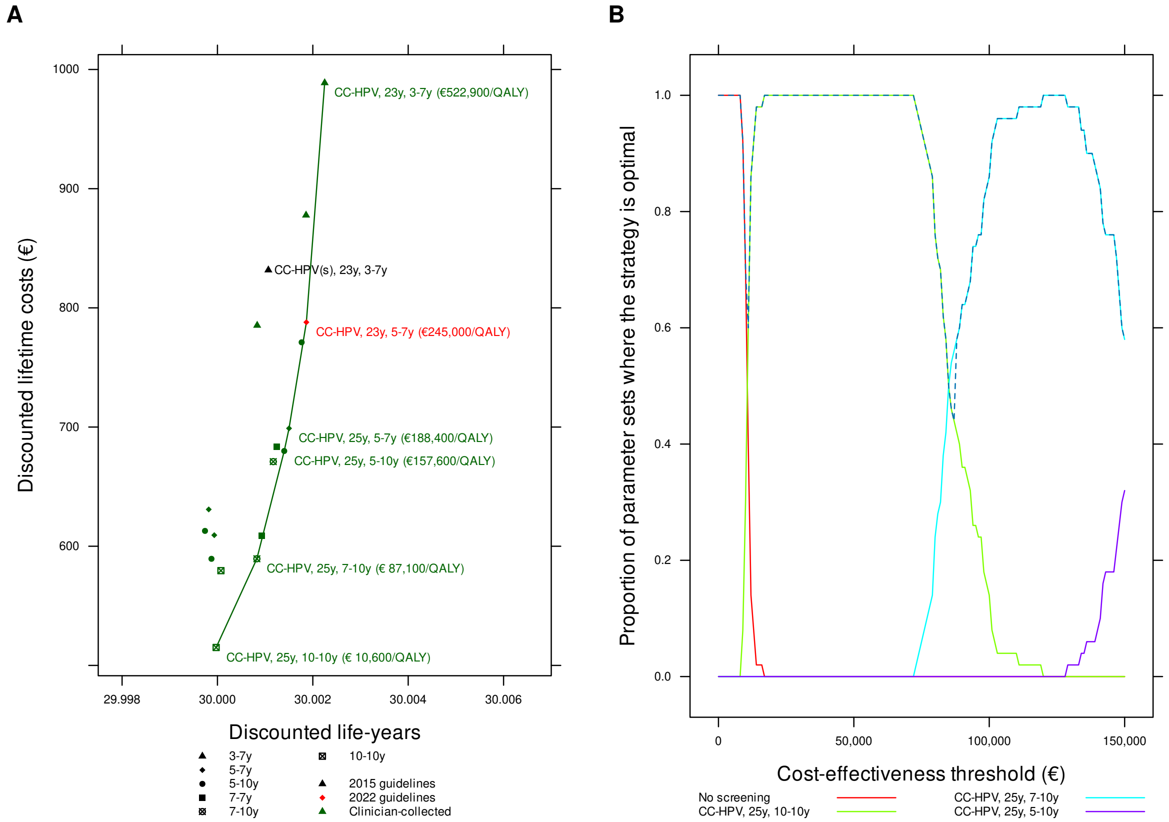

Figure G.7: Cost-effectiveness of cervical cancer screening strategies in unvaccinated cohorts in Sweden: *restricted to clinician-collected sampling, effectiveness measured using life expectancy*. Panel A: cost-efficiency frontier. Panel B: acceptability curves for different cost-effectiveness thresholds (the dashed line is the cost-effectiveness acceptability frontier).

| Strategy | Expected number of colposcopy referrals per woman over lifetime | Mean lifetime risk of cancer (%) | Proportion of cancers detected at local stage (%) | Life-years (undiscounted, from age 9 years) | Total cost per woman (€; discounted) | QALE (discounted) | Incremental cost (€; discounted) | Incremental QALEs (discounted) | ICER (Cost (€) per QALY gained) | Status |
|---|---|---|---|---|---|---|---|---|---|---|
| No screening | 0.000 | 1.744 | 67.10 | 75.69084 | 145.51 | 25.64434 | | | | ND |
| CC-HPV, 25y, 10-10y | 0.786 | 0.202 | 81.66 | 75.89505 | 515.05 | 25.68114 | 369.53 | 0.03680 | 10040 | ND |
| CC-HPV, 23y, 10-10y | 0.895 | 0.196 | 80.93 | 75.89520 | 579.52 | 25.68143 | | | | ED |
| CC-HPV(s), 25y, 5-10y | 0.739 | 0.198 | 83.74 | 75.89615 | 589.44 | 25.68070 | | | | D |
| CC-HPV, 25y, 7-10y | 0.887 | 0.173 | 83.30 | 75.89943 | 589.51 | 25.68210 | 74.46 | 0.00096 | 77594 | ND |
| CC-HPV, 25y, 7-7y | 0.920 | 0.158 | 84.44 | 75.90032 | 608.81 | 25.68219 | | | | ED |
| CC-HPV(s), 25y, 5-7y | 0.767 | 0.191 | 83.98 | 75.89664 | 609.29 | 25.68076 | | | | D |
| CC-HPV(s), 23y, 5-10y | 0.703 | 0.210 | 83.02 | 75.89514 | 612.87 | 25.68062 | | | | D |
| CC-HPV(s), 23y, 5-7y | 0.730 | 0.200 | 83.71 | 75.89583 | 630.89 | 25.68069 | | | | D |
| CC-HPV, 23y, 7-10y | 1.028 | 0.148 | 83.42 | 75.90099 | 671.00 | 25.68265 | | | | ED |
| CC-HPV, 25y, 5-10y | 1.005 | 0.142 | 84.44 | 75.90247 | 679.91 | 25.68276 | 90.40 | 0.00066 | 136229 | ND |
| CC-HPV, 23y, 7-7y | 1.046 | 0.144 | 83.94 | 75.90159 | 683.65 | 25.68272 | | | | D |
| CC-HPV, 25y, 5-7y | 1.035 | 0.133 | 85.29 | 75.90330 | 698.90 | 25.68286 | | | | ED |
| CC-HPV, 23y, 5-10y | 1.151 | 0.126 | 83.79 | 75.90396 | 771.10 | 25.68337 | 91.19 | 0.00060 | 151147 | ND |
| CC-HPV(s), 25y, 3-7y | 0.903 | 0.156 | 84.71 | 75.90072 | 785.28 | 25.68188 | | | | D |
| CC-HPV, 23y, 5-7y | 1.179 | 0.115 | 85.14 | 75.90480 | 787.93 | 25.68346 | 16.83 | 0.00009 | 180913 | ND |
| CC-HPV(s), 23y, 3-7y | 0.889 | 0.145 | 85.93 | 75.90172 | 831.74 | 25.68224 | | | | D |
| CC-HPV, 25y, 3-7y | 1.214 | 0.117 | 85.66 | 75.90498 | 877.75 | 25.68331 | | | | D |
| CC-HPV, 23y, 3-7y | 1.396 | 0.093 | 85.63 | 75.90667 | 988.78 | 25.68396 | 200.85 | 0.00050 | 398279 | ND |

Table G.7: Summary statistics, costs, life-years and ICERs on the cost-efficiency frontier, *restricted to clinician-collected sampling, effectiveness measured using life expectancy*, Sweden. Abbreviations: ICER=Incremental cost-effectiveness ratio; ND=Not dominated; D=Dominated; ED=Extended dominated.

### H Sensitivity analyses for all strategies

#### H.1 Direct medical costs

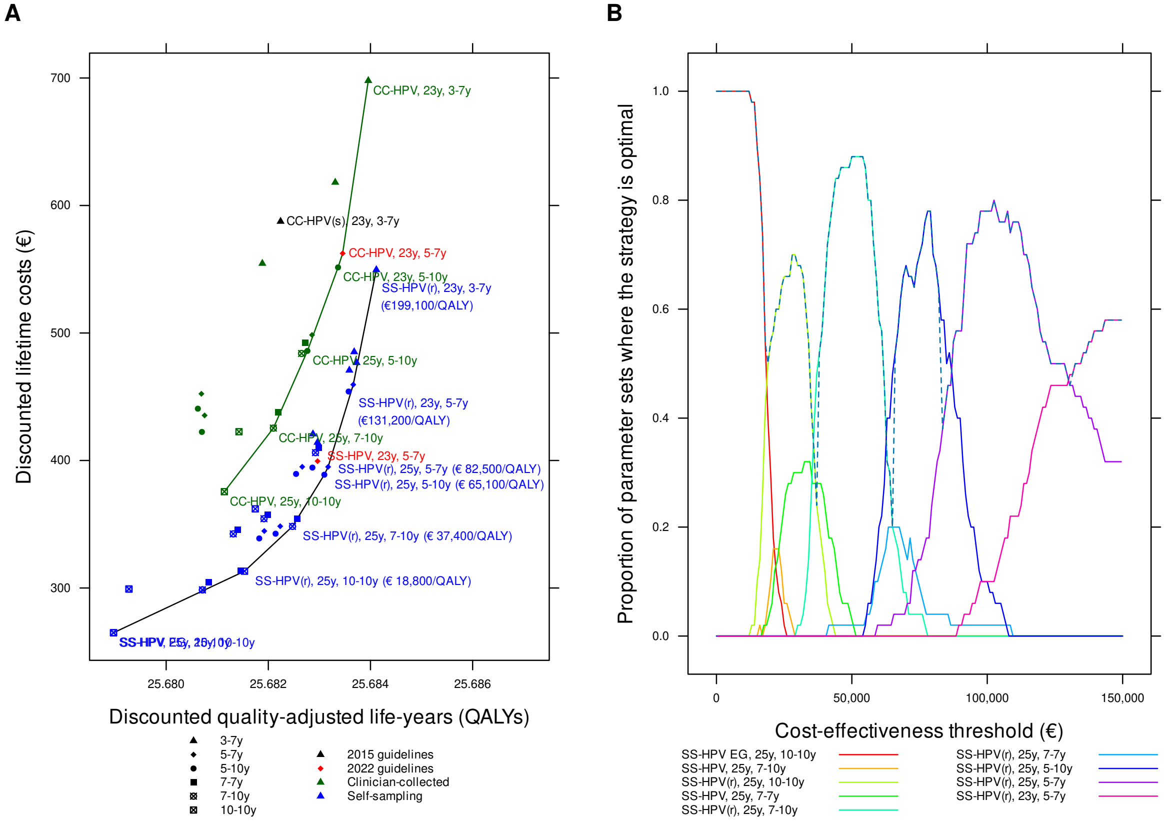

Figure H.1: Cost-effectiveness of cervical cancer screening strategies in unvaccinated cohorts in Sweden: *direct medical costs*. Panel A: cost-efficiency frontier for clinician-collected tests (green) and all testing (black). Panel B: acceptability curves for different cost-effectiveness thresholds (the dashed line is the cost-effectiveness acceptability frontier).

| Strategy | Expected number of colposcopy referrals per woman over lifetime | Mean lifetime risk of cancer (%) | Proportion of cancers detected at local stage (%) | Life-years (undiscounted, from age 9 years) | Total cost per woman (€; discounted) | QALE (discounted) | Incremental cost (€; discounted) | Incremental QALEs (discounted) | ICER (Cost (€) per QALY gained) | Status |
|---|---|---|---|---|---|---|---|---|---|---|
| SS-HPV EG, 25y, 10-10y | 0.626 | 0.277 | 79.46 | 75.88525 | 264.91 | 25.67897 | | | | ND |
| SS-HPV, 25y, 10-10y | 0.626 | 0.277 | 79.46 | 75.88525 | 264.91 | 25.67897 | | | | D |
| SS-HPV EG, 25y, 7-10y | 0.736 | 0.224 | 81.14 | 75.89366 | 298.52 | 25.68071 | | | | ED |
| SS-HPV, 25y, 7-10y | 0.736 | 0.224 | 81.14 | 75.89366 | 298.52 | 25.68071 | | | | D |
| SS-HPV EG, 23y, 10-10y | 0.711 | 0.274 | 79.07 | 75.88552 | 299.19 | 25.67927 | | | | D |
| SS-HPV, 23y, 10-10y | 0.711 | 0.274 | 79.07 | 75.88552 | 299.19 | 25.67927 | | | | D |
| SS-HPV EG, 25y, 7-7y | 0.763 | 0.205 | 82.27 | 75.89489 | 304.34 | 25.68084 | | | | ED |
| SS-HPV(r), 25y, 10-10y | 0.976 | 0.190 | 82.32 | 75.89666 | 313.17 | 25.68153 | 48.26 | 0.00256 | 18818 | ND |
| SS-HPV, 25y, 7-7y | 0.752 | 0.185 | 82.98 | 75.89743 | 313.67 | 25.68146 | | | | D |
| SS-HPV EG, 25y, 5-10y | 0.864 | 0.173 | 83.11 | 75.89866 | 338.88 | 25.68182 | | | | ED |
| SS-HPV EG, 23y, 7-10y | 0.856 | 0.197 | 82.41 | 75.89549 | 342.45 | 25.68131 | | | | D |
| SS-HPV, 25y, 5-10y | 0.824 | 0.163 | 83.28 | 75.89991 | 342.57 | 25.68214 | | | | ED |
| SS-HPV EG, 25y, 5-7y | 0.885 | 0.162 | 83.73 | 75.89952 | 344.68 | 25.68192 | | | | D |
| SS-HPV EG, 23y, 7-7y | 0.867 | 0.192 | 82.74 | 75.89621 | 345.61 | 25.68140 | | | | D |
| SS-HPV(r), 25y, 7-10y | 1.115 | 0.160 | 83.65 | 75.90091 | 348.31 | 25.68247 | 35.14 | 0.00094 | 37399 | ND |
| SS-HPV, 25y, 5-7y | 0.845 | 0.152 | 84.08 | 75.90071 | 348.42 | 25.68223 | | | | D |
| SS-HPV(r), 25y, 7-7y | 1.152 | 0.146 | 84.91 | 75.90173 | 354.25 | 25.68256 | | | | ED |
| SS-HPV, 23y, 7-10y | 0.849 | 0.174 | 82.75 | 75.89785 | 354.35 | 25.68191 | | | | D |
| SS-HPV, 23y, 7-7y | 0.858 | 0.170 | 82.89 | 75.89844 | 357.51 | 25.68198 | | | | D |
| SS-HPV(r), 23y, 10-10y | 1.123 | 0.183 | 81.41 | 75.89651 | 362.06 | 25.68174 | | | | D |
| CC-HPV, 25y, 10-10y | 0.786 | 0.202 | 81.66 | 75.89505 | 375.56 | 25.68114 | | | | D |
| SS-HPV(r), 25y, 5-10y | 1.277 | 0.127 | 84.87 | 75.90375 | 388.84 | 25.68310 | 40.53 | 0.00062 | 65110 | ND |
| SS-HPV EG, 23y, 5-10y | 0.992 | 0.162 | 82.52 | 75.90044 | 389.45 | 25.68254 | | | | D |
| SS-HPV, 23y, 5-10y | 0.950 | 0.149 | 82.81 | 75.90184 | 394.42 | 25.68286 | | | | D |
| SS-HPV(r), 25y, 5-7y | 1.312 | 0.120 | 85.55 | 75.90440 | 394.99 | 25.68317 | 6.15 | 0.00007 | 82507 | ND |
| SS-HPV EG, 23y, 5-7y | 1.017 | 0.144 | 84.10 | 75.90159 | 395.05 | 25.68266 | | | | D |
| SS-HPV, 23y, 5-7y | 0.970 | 0.135 | 84.39 | 75.90278 | 399.48 | 25.68296 | | | | D |
| SS-HPV(r), 23y, 7-10y | 1.304 | 0.136 | 84.61 | 75.90204 | 406.17 | 25.68292 | | | | D |
| SS-HPV(r), 23y, 7-7y | 1.321 | 0.133 | 84.76 | 75.90253 | 409.85 | 25.68299 | | | | D |
| SS-HPV, 25y, 3-7y | 0.989 | 0.131 | 84.60 | 75.90384 | 414.16 | 25.68296 | | | | D |
| SS-HPV EG, 25y, 3-7y | 1.086 | 0.134 | 84.54 | 75.90348 | 420.78 | 25.68287 | | | | D |
| CC-HPV(s), 25y, 5-10y | 0.739 | 0.198 | 83.74 | 75.89615 | 422.40 | 25.68070 | | | | D |
| CC-HPV, 23y, 10-10y | 0.895 | 0.196 | 80.93 | 75.89520 | 422.58 | 25.68143 | | | | D |
| CC-HPV, 25y, 7-10y | 0.887 | 0.173 | 83.30 | 75.89943 | 425.40 | 25.68210 | | | | D |
| CC-HPV(s), 25y, 5-7y | 0.766 | 0.191 | 83.98 | 75.89662 | 435.34 | 25.68075 | | | | D |
| CC-HPV, 25y, 7-7y | 0.920 | 0.158 | 84.44 | 75.90032 | 437.98 | 25.68219 | | | | D |
| CC-HPV(s), 23y, 5-10y | 0.703 | 0.210 | 83.02 | 75.89514 | 440.64 | 25.68062 | | | | D |
| CC-HPV(s), 23y, 5-7y | 0.729 | 0.200 | 83.71 | 75.89581 | 452.31 | 25.68069 | | | | D |
| SS-HPV(r), 23y, 5-10y | 1.476 | 0.118 | 84.34 | 75.90464 | 454.08 | 25.68357 | | | | ED |

Continued on next page

| Strategy | Expected number of colposcopy referrals per woman over life-time | Mean life-time risk of cancer (%) | Proportion of cancers detected at local stage (%) | Life-years (undis-counted, from age 9 years) | Total cost per woman (€; dis-counted) | QALE (dis-counted) | Incremental cost (€; dis-counted) | Incremental QALEs (dis-counted) | ICER (Cost (€) per QALY gained) | Status |
|---|---|---|---|---|---|---|---|---|---|---|
| SS-HPV(r), 23y, 5-7y | 1.510 | 0.106 | 86.31 | 75.90550 | 459.49 | 25.68366 | 64.50 | 0.00049 | 131193 | ND |
| SS-HPV(r), 25y, 3-7y | 1.566 | 0.106 | 86.02 | 75.90596 | 470.77 | 25.68359 | | | | D |
| SS-HPV, 23y, 3-7y | 1.147 | 0.103 | 85.18 | 75.90598 | 476.68 | 25.68373 | | | | ED |
| CC-HPV, 23y, 7-10y | 1.028 | 0.148 | 83.42 | 75.90099 | 483.93 | 25.68265 | | | | D |
| SS-HPV EG, 23y, 3-7y | 1.256 | 0.105 | 85.10 | 75.90577 | 485.19 | 25.68368 | | | | D |
| CC-HPV, 25y, 5-10y | 1.005 | 0.142 | 84.45 | 75.90248 | 485.96 | 25.68277 | | | | D |
| CC-HPV, 23y, 7-7y | 1.046 | 0.144 | 83.94 | 75.90159 | 492.23 | 25.68272 | | | | D |
| CC-HPV, 25y, 5-7y | 1.035 | 0.133 | 85.29 | 75.90330 | 498.45 | 25.68286 | | | | D |
| SS-HPV(r), 23y, 3-7y | 1.820 | 0.085 | 86.51 | 75.90727 | 549.58 | 25.68411 | 90.09 | 0.00045 | 199139 | ND |
| CC-HPV, 23y, 5-10y | 1.151 | 0.126 | 83.78 | 75.90395 | 551.45 | 25.68337 | | | | D |
| CC-HPV(s), 25y, 3-7y | 0.903 | 0.156 | 84.71 | 75.90072 | 554.52 | 25.68188 | | | | D |
| CC-HPV, 23y, 5-7y | 1.179 | 0.115 | 85.15 | 75.90479 | 562.43 | 25.68346 | | | | D |
| CC-HPV(s), 23y, 3-7y | 0.889 | 0.145 | 85.93 | 75.90172 | 587.48 | 25.68224 | | | | D |
| CC-HPV, 25y, 3-7y | 1.214 | 0.117 | 85.65 | 75.90497 | 618.12 | 25.68331 | | | | D |
| CC-HPV, 23y, 3-7y | 1.397 | 0.093 | 85.64 | 75.90666 | 697.89 | 25.68396 | | | | D |

Table H.1: Summary statistics, costs, quality-adjusted life-years and ICERs on the cost-efficiency frontier, *direct medical costs only*, Sweden. Abbreviations: QALE=Quality-adjusted life expectancy; QALY: Quality-adjusted life-year; ICER=Incremental cost-effectiveness ratio; ND=Not dominated; D=Dominated; ED=Extended dominated.

#### H.2 Assumed lower compliance

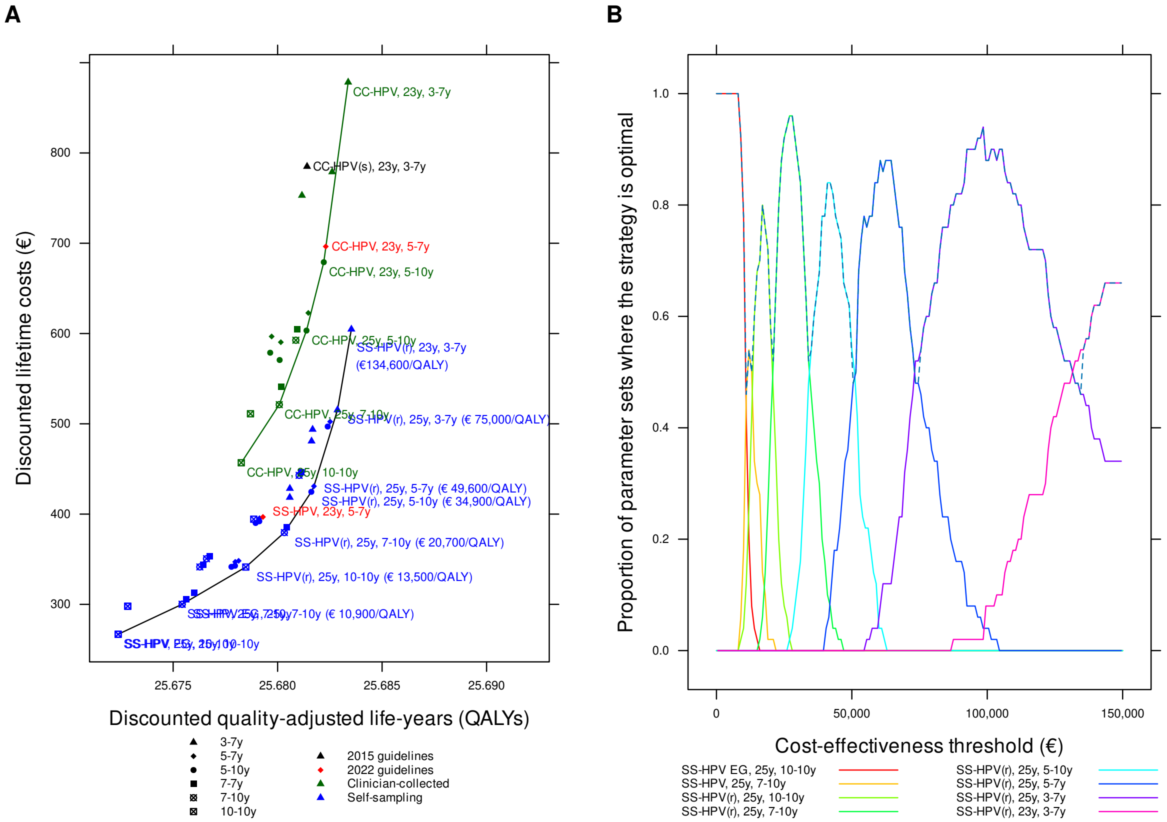

Figure H.2: Cost-effectiveness of cervical cancer screening strategies in unvaccinated cohorts in Sweden: *assumed lower compliance*. Panel A: cost-efficiency frontier for clinician-collected tests (green) and all testing (black). Panel B: acceptability curves for different cost-effectiveness thresholds (the dashed line is the cost-effectiveness acceptability frontier).

| Strategy | Expected number of colposcopy referrals per woman over life-time | Mean life-time risk of cancer (%) | Proportion of cancers detected at local stage (%) | Life-years (undis-counted, from age 9 years) | Total cost per woman (€; dis-counted) | QALE (dis-counted) | Incremental cost (€; dis-counted) | Incremental QALEs (dis-counted) | ICER (Cost (€) per QALY gained) | Status |
|---|---|---|---|---|---|---|---|---|---|---|
| SS-HPV EG, 25y, 10-10y | 0.445 | 0.523 | 74.74 | 75.85301 | 266.88 | 25.67237 | | | | ND |
| SS-HPV, 25y, 10-10y | 0.445 | 0.523 | 74.74 | 75.85301 | 266.88 | 25.67237 | | | | D |
| SS-HPV EG, 23y, 10-10y | 0.505 | 0.515 | 74.62 | 75.85406 | 297.94 | 25.67283 | | | | ED |
| SS-HPV, 23y, 10-10y | 0.505 | 0.515 | 74.62 | 75.85406 | 297.94 | 25.67283 | | | | D |
| SS-HPV EG, 25y, 7-10y | 0.536 | 0.419 | 76.38 | 75.86890 | 300.11 | 25.67543 | 33.24 | 0.00306 | 10857 | ND |
| SS-HPV, 25y, 7-10y | 0.536 | 0.419 | 76.38 | 75.86890 | 300.11 | 25.67543 | | | | D |
| SS-HPV EG, 25y, 7-7y | 0.556 | 0.392 | 77.18 | 75.87080 | 305.47 | 25.67563 | | | | ED |
| SS-HPV, 25y, 7-7y | 0.538 | 0.380 | 77.25 | 75.87228 | 312.75 | 25.67601 | | | | ED |
| SS-HPV(r), 25y, 10-10y | 0.822 | 0.302 | 78.57 | 75.88245 | 341.35 | 25.67848 | 41.24 | 0.00305 | 13529 | ND |
| SS-HPV EG, 23y, 7-10y | 0.624 | 0.384 | 77.10 | 75.87194 | 341.48 | 25.67628 | | | | D |
| SS-HPV EG, 25y, 5-10y | 0.644 | 0.324 | 78.07 | 75.88044 | 341.59 | 25.67779 | | | | D |
| SS-HPV, 25y, 5-10y | 0.604 | 0.319 | 77.96 | 75.88096 | 342.76 | 25.67796 | | | | D |
| SS-HPV EG, 23y, 7-7y | 0.632 | 0.372 | 77.39 | 75.87329 | 344.16 | 25.67644 | | | | D |
| SS-HPV EG, 25y, 5-7y | 0.662 | 0.305 | 78.73 | 75.88206 | 347.08 | 25.67797 | | | | D |
| SS-HPV, 25y, 5-7y | 0.620 | 0.300 | 78.64 | 75.88253 | 348.22 | 25.67813 | | | | D |
| SS-HPV, 23y, 7-10y | 0.606 | 0.371 | 77.06 | 75.87311 | 350.57 | 25.67660 | | | | D |
| SS-HPV, 23y, 7-7y | 0.613 | 0.360 | 77.33 | 75.87433 | 353.19 | 25.67674 | | | | D |
| SS-HPV(r), 25y, 7-10y | 0.948 | 0.239 | 80.33 | 75.89145 | 379.57 | 25.68032 | 38.22 | 0.00184 | 20715 | ND |
| SS-HPV(r), 25y, 7-7y | 0.981 | 0.220 | 81.28 | 75.89263 | 385.49 | 25.68045 | | | | ED |
| SS-HPV EG, 23y, 5-10y | 0.741 | 0.301 | 77.75 | 75.88455 | 390.14 | 25.67895 | | | | D |
| SS-HPV, 23y, 5-10y | 0.696 | 0.295 | 77.75 | 75.88527 | 392.06 | 25.67912 | | | | D |
| SS-HPV(r), 23y, 10-10y | 0.946 | 0.292 | 78.20 | 75.88325 | 394.37 | 25.67886 | | | | D |
| SS-HPV EG, 23y, 5-7y | 0.760 | 0.277 | 78.79 | 75.88632 | 395.41 | 25.67913 | | | | D |
| SS-HPV, 23y, 5-7y | 0.711 | 0.274 | 78.73 | 75.88694 | 396.83 | 25.67930 | | | | D |
| SS-HPV, 25y, 3-7y | 0.754 | 0.225 | 80.84 | 75.89399 | 418.45 | 25.68058 | | | | ED |
| SS-HPV(r), 25y, 5-10y | 1.097 | 0.186 | 82.10 | 75.89743 | 424.66 | 25.68161 | 45.09 | 0.00129 | 34937 | ND |
| SS-HPV EG, 25y, 3-7y | 0.843 | 0.222 | 80.83 | 75.89403 | 428.35 | 25.68059 | | | | D |
| SS-HPV(r), 25y, 5-7y | 1.129 | 0.173 | 82.80 | 75.89857 | 430.88 | 25.68174 | 6.22 | 0.00013 | 49599 | ND |
| SS-HPV(r), 23y, 7-10y | 1.110 | 0.209 | 80.99 | 75.89389 | 442.92 | 25.68103 | | | | D |
| SS-HPV(r), 23y, 7-7y | 1.124 | 0.204 | 81.26 | 75.89468 | 446.38 | 25.68112 | | | | D |
| CC-HPV, 25y, 10-10y | 0.677 | 0.294 | 79.00 | 75.88249 | 456.87 | 25.67826 | | | | D |
| SS-HPV, 23y, 3-7y | 0.875 | 0.188 | 81.24 | 75.89717 | 480.82 | 25.68162 | | | | D |
| SS-HPV EG, 23y, 3-7y | 0.976 | 0.186 | 81.55 | 75.89735 | 493.81 | 25.68168 | | | | D |
| SS-HPV(r), 23y, 5-10y | 1.270 | 0.170 | 81.63 | 75.89991 | 496.99 | 25.68240 | | | | ED |
| SS-HPV(r), 23y, 5-7y | 1.300 | 0.154 | 83.08 | 75.90099 | 502.39 | 25.68251 | | | | ED |
| CC-HPV, 23y, 10-10y | 0.768 | 0.283 | 78.54 | 75.88339 | 511.13 | 25.67870 | | | | D |
| SS-HPV(r), 25y, 3-7y | 1.370 | 0.137 | 83.95 | 75.90324 | 515.41 | 25.68287 | 84.53 | 0.00113 | 74966 | ND |
| CC-HPV, 25y, 7-10y | 0.770 | 0.237 | 81.13 | 75.89125 | 521.25 | 25.68008 | | | | D |
| CC-HPV, 25y, 7-7y | 0.803 | 0.222 | 81.79 | 75.89217 | 540.91 | 25.68018 | | | | D |

Continued on next page

| Strategy | Expected number of colposcopy referrals per woman over lifetime | Mean lifetime risk of cancer (%) | Proportion of cancers detected at local stage (%) | Life-years (undiscounted, from age 9 years) | Total cost per woman (€; discounted) | QALE (discounted) | Incremental cost (€; discounted) | Incremental QALEs (discounted) | ICER (Cost (€) per QALY gained) | Status |
|---|---|---|---|---|---|---|---|---|---|---|
| CC-HPV(s), 25y, 5-10y | 0.718 | 0.214 | 83.13 | 75.89420 | 570.67 | 25.68010 | | | | D |
| CC-HPV(s), 23y, 5-10y | 0.670 | 0.237 | 82.14 | 75.89190 | 578.74 | 25.67964 | | | | D |
| CC-HPV(s), 25y, 5-7y | 0.745 | 0.207 | 83.31 | 75.89470 | 590.40 | 25.68015 | | | | D |
| CC-HPV, 23y, 7-10y | 0.894 | 0.206 | 81.43 | 75.89402 | 592.59 | 25.68087 | | | | D |
| CC-HPV(s), 23y, 5-7y | 0.695 | 0.227 | 82.73 | 75.89259 | 596.73 | 25.67971 | | | | D |
| CC-HPV, 25y, 5-10y | 0.884 | 0.185 | 82.61 | 75.89730 | 603.28 | 25.68138 | | | | D |
| CC-HPV, 23y, 7-7y | 0.910 | 0.203 | 81.71 | 75.89459 | 604.74 | 25.68093 | | | | D |
| SS-HPV(r), 23y, 3-7y | 1.595 | 0.112 | 84.72 | 75.90501 | 604.76 | 25.68353 | 89.35 | 0.00066 | 134600 | ND |
| CC-HPV, 25y, 5-7y | 0.915 | 0.176 | 83.08 | 75.89808 | 622.80 | 25.68147 | | | | D |
| CC-HPV, 23y, 5-10y | 1.005 | 0.167 | 82.41 | 75.89974 | 679.02 | 25.68221 | | | | D |
| CC-HPV, 23y, 5-7y | 1.034 | 0.155 | 83.52 | 75.90064 | 696.38 | 25.68230 | | | | D |
| CC-HPV(s), 25y, 3-7y | 0.869 | 0.177 | 83.99 | 75.89844 | 752.97 | 25.68116 | | | | D |
| CC-HPV, 25y, 3-7y | 1.081 | 0.140 | 84.65 | 75.90270 | 779.01 | 25.68261 | | | | D |
| CC-HPV(s), 23y, 3-7y | 0.849 | 0.170 | 85.21 | 75.89911 | 785.01 | 25.68141 | | | | D |
| CC-HPV, 23y, 3-7y | 1.245 | 0.114 | 85.05 | 75.90474 | 878.36 | 25.68338 | | | | D |

Table H.2: Summary statistics, costs, quality-adjusted life-years and ICERs on the cost-efficiency frontier, *assumed lower compliance*, Sweden. Abbreviations: QALE=Quality-adjusted life expectancy; QALY: Quality-adjusted life-year; ICER=Incremental cost-effectiveness ratio; ND=Not dominated; D=Dominated; ED=Extended dominated.

#### H.3 Undiscounted QALYs and costs

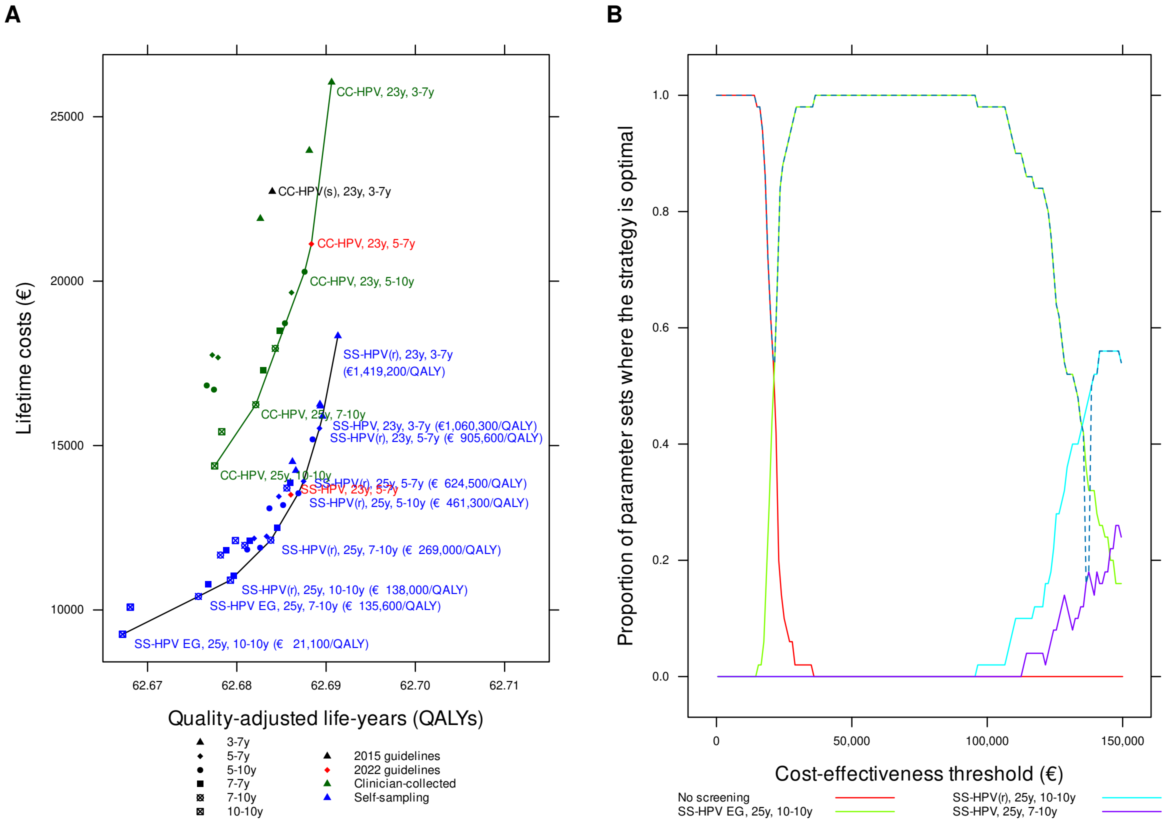

Figure H.3: Cost-effectiveness of cervical cancer screening strategies in unvaccinated cohorts in Sweden: *undiscounted*. Panel A: cost-efficiency frontier for clinician-collected tests (green) and all testing (black). Panel B: acceptability curves for different cost-effectiveness thresholds.

| Strategy | Expected number of colposcopy referrals per woman over life-time | Mean life-time risk of cancer (%) | Proportion of cancers detected at local stage (%) | Life-years (undis-counted, from age 9 years) | Total cost per woman (€; undis-counted) | QALE (undis-counted) | Incremental cost (€; undis-counted) | Incremental QALEs (undis-counted) | ICER (Cost (€) per QALY gained) | Status |
|---|---|---|---|---|---|---|---|---|---|---|
| No screening | 0.000 | 1.744 | 67.10 | 75.69084 | 5241.66 | 62.47672 | | | | ND |
| SS-HPV EG, 25y, 10-10y | 0.626 | 0.277 | 79.46 | 75.88525 | 9260.64 | 62.66721 | 4018.97 | 0.19049 | 21098 | ND |
| SS-HPV, 25y, 10-10y | 0.626 | 0.278 | 79.44 | 75.88523 | 9261.87 | 62.66718 | | | | D |
| SS-HPV, 23y, 10-10y | 0.710 | 0.274 | 79.08 | 75.88550 | 10084.49 | 62.66806 | | | | ED |
| SS-HPV EG, 23y, 10-10y | 0.711 | 0.274 | 79.07 | 75.88552 | 10089.55 | 62.66809 | | | | ED |
| SS-HPV EG, 25y, 7-10y | 0.736 | 0.224 | 81.14 | 75.89366 | 10411.48 | 62.67570 | 1150.84 | 0.00849 | 135551 | ND |
| SS-HPV, 25y, 7-10y | 0.736 | 0.224 | 81.14 | 75.89366 | 10411.48 | 62.67570 | | | | D |
| SS-HPV EG, 25y, 7-7y | 0.763 | 0.205 | 82.27 | 75.89489 | 10789.48 | 62.67681 | | | | ED |
| SS-HPV(r), 25y, 10-10y | 0.976 | 0.190 | 82.32 | 75.89666 | 10906.29 | 62.67929 | 494.81 | 0.00359 | 137964 | ND |
| SS-HPV, 25y, 7-7y | 0.751 | 0.185 | 82.99 | 75.89743 | 11038.62 | 62.67964 | | | | ED |
| SS-HPV EG, 23y, 7-10y | 0.856 | 0.197 | 82.41 | 75.89549 | 11670.04 | 62.67817 | | | | D |
| SS-HPV EG, 23y, 7-7y | 0.867 | 0.192 | 82.75 | 75.89619 | 11810.64 | 62.67879 | | | | D |
| SS-HPV EG, 25y, 5-10y | 0.864 | 0.173 | 83.11 | 75.89866 | 11837.55 | 62.68116 | | | | ED |
| SS-HPV, 25y, 5-10y | 0.824 | 0.163 | 83.28 | 75.89991 | 11893.32 | 62.68261 | | | | ED |
| SS-HPV, 23y, 7-10y | 0.849 | 0.174 | 82.75 | 75.89785 | 11963.84 | 62.68090 | | | | D |
| SS-HPV, 23y, 7-7y | 0.858 | 0.170 | 82.89 | 75.89845 | 12111.29 | 62.68142 | | | | D |
| SS-HPV(r), 23y, 10-10y | 1.124 | 0.183 | 81.41 | 75.89652 | 12111.72 | 62.67985 | | | | D |
| SS-HPV(r), 25y, 7-10y | 1.115 | 0.160 | 83.64 | 75.90093 | 12121.52 | 62.68380 | 1215.24 | 0.00452 | 269001 | ND |
| SS-HPV EG, 25y, 5-7y | 0.885 | 0.162 | 83.73 | 75.89952 | 12175.86 | 62.68194 | | | | D |
| SS-HPV, 25y, 5-7y | 0.845 | 0.152 | 84.08 | 75.90071 | 12239.18 | 62.68334 | | | | D |
| SS-HPV(r), 25y, 7-7y | 1.153 | 0.146 | 84.91 | 75.90172 | 12504.76 | 62.68454 | | | | ED |
| SS-HPV EG, 23y, 5-10y | 0.992 | 0.162 | 82.52 | 75.90044 | 13090.95 | 62.68365 | | | | D |
| SS-HPV, 23y, 5-10y | 0.950 | 0.149 | 82.81 | 75.90184 | 13190.40 | 62.68519 | | | | ED |
| SS-HPV EG, 23y, 5-7y | 1.017 | 0.144 | 84.10 | 75.90159 | 13450.80 | 62.68471 | | | | D |
| SS-HPV, 23y, 5-7y | 0.970 | 0.135 | 84.39 | 75.90278 | 13504.21 | 62.68605 | | | | ED |
| SS-HPV(r), 25y, 5-10y | 1.277 | 0.127 | 84.87 | 75.90375 | 13549.04 | 62.68690 | 1427.52 | 0.00309 | 461331 | ND |
| SS-HPV(r), 23y, 7-10y | 1.304 | 0.135 | 84.62 | 75.90205 | 13707.29 | 62.68561 | | | | D |
| SS-HPV(r), 23y, 7-7y | 1.321 | 0.133 | 84.76 | 75.90252 | 13876.51 | 62.68600 | | | | D |
| SS-HPV(r), 25y, 5-7y | 1.313 | 0.120 | 85.54 | 75.90440 | 13912.48 | 62.68748 | 363.44 | 0.00058 | 624525 | ND |
| SS-HPV, 25y, 3-7y | 0.989 | 0.131 | 84.60 | 75.90384 | 14239.54 | 62.68661 | | | | D |
| CC-HPV, 25y, 10-10y | 0.786 | 0.202 | 81.66 | 75.89505 | 14380.39 | 62.67752 | | | | D |
| SS-HPV EG, 25y, 3-7y | 1.086 | 0.134 | 84.55 | 75.90349 | 14509.52 | 62.68623 | | | | D |
| SS-HPV(r), 23y, 5-10y | 1.476 | 0.118 | 84.34 | 75.90464 | 15187.81 | 62.68849 | | | | ED |
| CC-HPV, 23y, 10-10y | 0.895 | 0.196 | 80.93 | 75.89520 | 15420.71 | 62.67831 | | | | D |
| SS-HPV(r), 23y, 5-7y | 1.510 | 0.106 | 86.31 | 75.90550 | 15523.94 | 62.68926 | 1611.46 | 0.00178 | 905575 | ND |
| SS-HPV, 23y, 3-7y | 1.147 | 0.103 | 85.18 | 75.90598 | 15888.35 | 62.68960 | 364.41 | 0.00034 | 1060275 | ND |
| SS-HPV EG, 23y, 3-7y | 1.256 | 0.105 | 85.10 | 75.90575 | 16199.99 | 62.68936 | | | | D |
| CC-HPV, 25y, 7-10y | 0.887 | 0.173 | 83.30 | 75.89943 | 16243.68 | 62.68212 | | | | D |
| SS-HPV(r), 25y, 3-7y | 1.566 | 0.106 | 86.02 | 75.90596 | 16256.35 | 62.68931 | | | | D |

Continued on next page

| Strategy | Expected number of colposcopy referrals per woman over life-time | Mean life-time risk of cancer (%) | Proportion of cancers detected at local stage (%) | Life-years (undis-counted, from age 9 years) | Total cost per woman (€; undis-counted) | QALE (undis-counted) | Incremental cost (€; undis-counted) | Incremental QALEs (undis-counted) | ICER (Cost (€) per QALY gained) | Status |
|---|---|---|---|---|---|---|---|---|---|---|
| CC-HPV(s), 25y, 5-10y | 0.739 | 0.198 | 83.74 | 75.89615 | 16700.90 | 62.67744 | | | | D |
| CC-HPV(s), 23y, 5-10y | 0.703 | 0.210 | 83.02 | 75.89514 | 16827.49 | 62.67663 | | | | D |
| CC-HPV, 25y, 7-7y | 0.920 | 0.158 | 84.44 | 75.90032 | 17287.44 | 62.68294 | | | | D |
| CC-HPV(s), 25y, 5-7y | 0.767 | 0.191 | 83.98 | 75.89664 | 17677.42 | 62.67790 | | | | D |
| CC-HPV(s), 23y, 5-7y | 0.730 | 0.200 | 83.71 | 75.89583 | 17752.43 | 62.67725 | | | | D |
| CC-HPV, 23y, 7-10y | 1.028 | 0.148 | 83.42 | 75.90099 | 17953.46 | 62.68431 | | | | D |
| SS-HPV(r), 23y, 3-7y | 1.820 | 0.085 | 86.51 | 75.90727 | 18335.23 | 62.69133 | 2446.88 | 0.00172 | 1419173 | ND |
| CC-HPV, 23y, 7-7y | 1.046 | 0.144 | 83.94 | 75.90159 | 18493.44 | 62.68483 | | | | D |
| CC-HPV, 25y, 5-10y | 1.005 | 0.142 | 84.44 | 75.90247 | 18719.51 | 62.68539 | | | | D |
| CC-HPV, 25y, 5-7y | 1.035 | 0.133 | 85.29 | 75.90330 | 19651.39 | 62.68613 | | | | D |
| CC-HPV, 23y, 5-10y | 1.151 | 0.126 | 83.79 | 75.90396 | 20285.13 | 62.68759 | | | | D |
| CC-HPV, 23y, 5-7y | 1.179 | 0.115 | 85.14 | 75.90480 | 21130.59 | 62.68836 | | | | D |
| CC-HPV(s), 25y, 3-7y | 0.903 | 0.156 | 84.71 | 75.90072 | 21901.96 | 62.68264 | | | | D |
| CC-HPV(s), 23y, 3-7y | 0.889 | 0.145 | 85.93 | 75.90172 | 22726.39 | 62.68397 | | | | D |
| CC-HPV, 25y, 3-7y | 1.214 | 0.117 | 85.66 | 75.90498 | 23972.60 | 62.68812 | | | | D |
| CC-HPV, 23y, 3-7y | 1.396 | 0.093 | 85.63 | 75.90667 | 26049.52 | 62.69062 | | | | D |

Table H.3: Summary statistics, costs, quality-adjusted life-years and ICERs on the cost-efficiency frontier, *undiscounted*, Sweden. Abbreviations: QALE=Quality-adjusted life expectancy; QALY: Quality-adjusted life-year; ICER=Incremental cost-effectiveness ratio; ND=Not dominated; D=Dominated; ED=Extended dominated.

#### H.4 5% discounted QALYs and costs

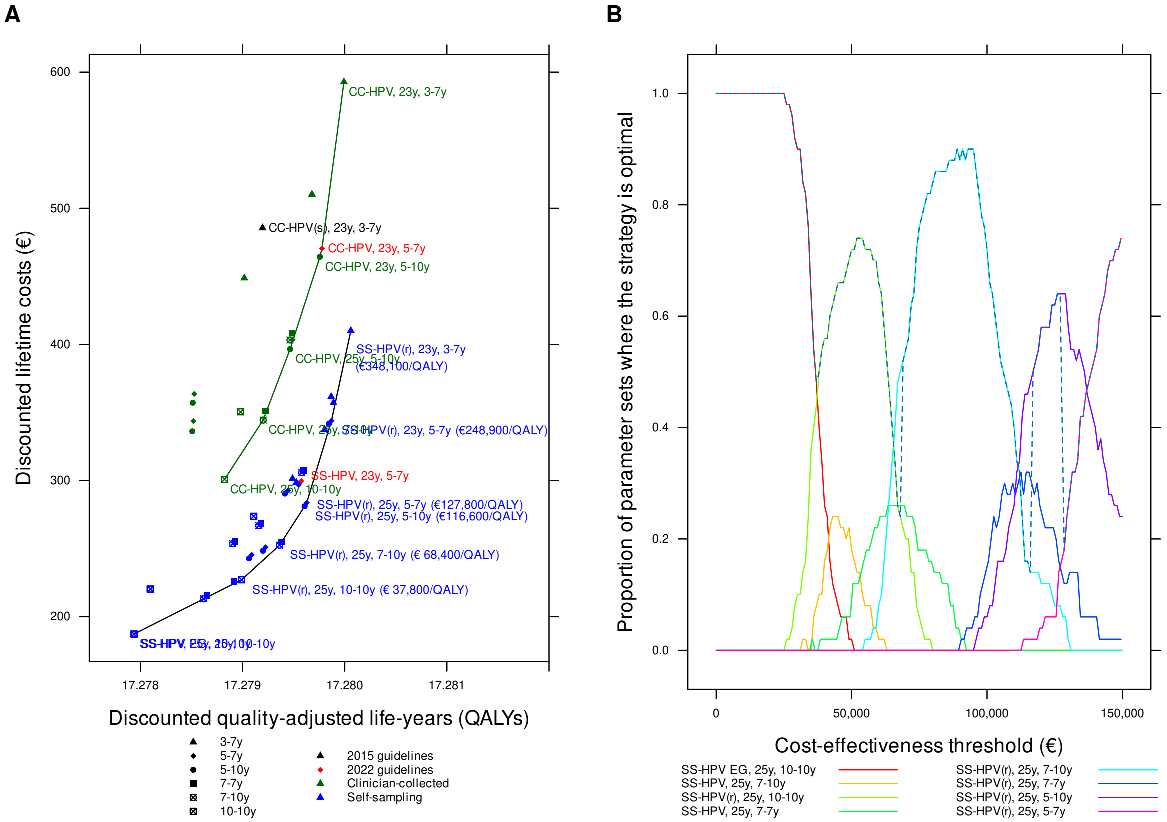

Figure H.4: Cost-effectiveness of cervical cancer screening strategies in unvaccinated cohorts in Sweden: *5% discount rate for QALYs and costs*. Panel A: cost-efficiency frontier for clinician-collected tests (green) and all testing (black). Panel B: acceptability curves for different cost-effectiveness thresholds (the dashed line is the cost-effectiveness acceptability frontier).

| Strategy | Expected number of colposcopy referrals per woman over lifetime | Mean lifetime risk of cancer (%) | Proportion of cancers detected at local stage (%) | Life-years (undiscounted, from age 9 years) | Total cost per woman (€; discounted) | QALE (discounted) | Incremental cost (€; discounted) | Incremental QALEs (discounted) | ICER (Cost (€) per QALY gained) | Status |
|---|---|---|---|---|---|---|---|---|---|---|
| SS-HPV EG, 25y, 10-10y | 0.626 | 0.277 | 79.46 | 75.88525 | 187.24 | 17.27794 | | | | ND |
| SS-HPV, 25y, 10-10y | 0.626 | 0.277 | 79.46 | 75.88525 | 187.24 | 17.27794 | | | | D |
| SS-HPV EG, 25y, 7-10y | 0.736 | 0.224 | 81.14 | 75.89366 | 213.17 | 17.27862 | | | | ED |
| SS-HPV, 25y, 7-10y | 0.736 | 0.224 | 81.14 | 75.89366 | 213.17 | 17.27862 | | | | D |
| SS-HPV EG, 25y, 7-7y | 0.763 | 0.205 | 82.27 | 75.89489 | 215.61 | 17.27865 | | | | ED |
| SS-HPV EG, 23y, 10-10y | 0.711 | 0.274 | 79.07 | 75.88552 | 220.28 | 17.27810 | | | | D |
| SS-HPV, 23y, 10-10y | 0.711 | 0.274 | 79.07 | 75.88552 | 220.28 | 17.27810 | | | | D |
| SS-HPV, 25y, 7-7y | 0.751 | 0.185 | 82.99 | 75.89743 | 225.97 | 17.27891 | | | | ED |
| SS-HPV(r), 25y, 10-10y | 0.976 | 0.190 | 82.32 | 75.89666 | 227.09 | 17.27899 | 39.85 | 0.00105 | 37835 | ND |
| SS-HPV EG, 25y, 5-10y | 0.864 | 0.173 | 83.11 | 75.89866 | 242.84 | 17.27906 | | | | ED |
| SS-HPV EG, 25y, 5-7y | 0.885 | 0.162 | 83.73 | 75.89952 | 245.42 | 17.27909 | | | | ED |
| SS-HPV, 25y, 5-10y | 0.824 | 0.163 | 83.28 | 75.89991 | 248.41 | 17.27920 | | | | ED |
| SS-HPV, 25y, 5-7y | 0.845 | 0.152 | 84.08 | 75.90071 | 250.96 | 17.27922 | | | | ED |
| SS-HPV(r), 25y, 7-10y | 1.115 | 0.160 | 83.65 | 75.90091 | 252.53 | 17.27936 | 25.44 | 0.00037 | 68427 | ND |
| SS-HPV EG, 23y, 7-10y | 0.856 | 0.197 | 82.41 | 75.89549 | 253.62 | 17.27890 | | | | D |
| SS-HPV(r), 25y, 7-7y | 1.153 | 0.146 | 84.91 | 75.90170 | 254.99 | 17.27938 | | | | ED |
| SS-HPV EG, 23y, 7-7y | 0.867 | 0.192 | 82.74 | 75.89621 | 255.22 | 17.27893 | | | | D |
| SS-HPV, 23y, 7-10y | 0.849 | 0.174 | 82.75 | 75.89785 | 266.83 | 17.27916 | | | | D |
| SS-HPV, 23y, 7-7y | 0.858 | 0.170 | 82.89 | 75.89844 | 268.39 | 17.27918 | | | | D |
| SS-HPV(r), 23y, 10-10y | 1.123 | 0.183 | 81.41 | 75.89651 | 273.76 | 17.27911 | | | | D |
| SS-HPV(r), 25y, 5-10y | 1.277 | 0.127 | 84.87 | 75.90374 | 281.09 | 17.27961 | 28.56 | 0.00024 | 116606 | ND |
| SS-HPV(r), 25y, 5-7y | 1.313 | 0.120 | 85.54 | 75.90440 | 283.72 | 17.27963 | 2.63 | 0.00002 | 127793 | ND |
| SS-HPV EG, 23y, 5-10y | 0.992 | 0.162 | 82.52 | 75.90044 | 290.44 | 17.27941 | | | | D |
| SS-HPV EG, 23y, 5-7y | 1.017 | 0.144 | 84.10 | 75.90159 | 292.80 | 17.27945 | | | | D |
| SS-HPV, 23y, 5-10y | 0.950 | 0.149 | 82.81 | 75.90184 | 297.53 | 17.27955 | | | | D |
| SS-HPV, 25y, 3-7y | 0.989 | 0.131 | 84.60 | 75.90384 | 298.46 | 17.27953 | | | | D |
| SS-HPV, 23y, 5-7y | 0.970 | 0.135 | 84.39 | 75.90278 | 299.70 | 17.27957 | | | | D |
| CC-HPV, 25y, 10-10y | 0.786 | 0.202 | 81.66 | 75.89505 | 300.86 | 17.27882 | | | | D |
| SS-HPV EG, 25y, 3-7y | 1.086 | 0.134 | 84.55 | 75.90349 | 301.40 | 17.27949 | | | | D |
| SS-HPV(r), 23y, 7-10y | 1.304 | 0.136 | 84.61 | 75.90204 | 305.85 | 17.27958 | | | | D |
| SS-HPV(r), 23y, 7-7y | 1.321 | 0.134 | 84.77 | 75.90251 | 307.45 | 17.27959 | | | | D |
| CC-HPV(s), 25y, 5-10y | 0.739 | 0.198 | 83.74 | 75.89615 | 336.15 | 17.27851 | | | | D |
| SS-HPV(r), 25y, 3-7y | 1.566 | 0.106 | 86.02 | 75.90596 | 337.52 | 17.27980 | | | | ED |
| SS-HPV(r), 23y, 5-10y | 1.476 | 0.118 | 84.34 | 75.90462 | 341.59 | 17.27984 | | | | ED |
| CC-HPV(s), 25y, 5-7y | 0.767 | 0.191 | 83.97 | 75.89660 | 343.61 | 17.27852 | | | | D |
| SS-HPV(r), 23y, 5-7y | 1.510 | 0.106 | 86.31 | 75.90550 | 343.97 | 17.27987 | 60.25 | 0.00024 | 248877 | ND |
| CC-HPV, 25y, 7-10y | 0.887 | 0.173 | 83.30 | 75.89943 | 344.43 | 17.27920 | | | | D |
| CC-HPV, 23y, 10-10y | 0.895 | 0.195 | 80.94 | 75.89521 | 350.50 | 17.27898 | | | | D |
| CC-HPV, 25y, 7-7y | 0.920 | 0.158 | 84.43 | 75.90032 | 351.12 | 17.27922 | | | | D |

Continued on next page

| Strategy | Expected number of colposcopy referrals per woman over lifetime | Mean lifetime risk of cancer (%) | Proportion of cancers detected at local stage (%) | Life-years (undiscounted, from age 9 years) | Total cost per woman (€; discounted) | QALE (discounted) | Incremental cost (€; discounted) | Incremental QALEs (discounted) | ICER (Cost (€) per QALY gained) | Status |
|---|---|---|---|---|---|---|---|---|---|---|
| SS-HPV, 23y, 3-7y | 1.147 | 0.103 | 85.18 | 75.90598 | 357.08 | 17.27989 | | | | ED |
| CC-HPV(s), 23y, 5-10y | 0.703 | 0.210 | 83.03 | 75.89516 | 357.14 | 17.27851 | | | | D |
| SS-HPV EG, 23y, 3-7y | 1.256 | 0.105 | 85.10 | 75.90575 | 361.55 | 17.27987 | | | | D |
| CC-HPV(s), 23y, 5-7y | 0.729 | 0.200 | 83.71 | 75.89581 | 363.59 | 17.27853 | | | | D |
| CC-HPV, 25y, 5-10y | 1.005 | 0.142 | 84.44 | 75.90248 | 396.53 | 17.27947 | | | | D |
| CC-HPV, 23y, 7-10y | 1.028 | 0.148 | 83.42 | 75.90099 | 403.25 | 17.27946 | | | | D |
| CC-HPV, 25y, 5-7y | 1.035 | 0.133 | 85.29 | 75.90330 | 403.54 | 17.27949 | | | | D |
| CC-HPV, 23y, 7-7y | 1.046 | 0.144 | 83.94 | 75.90159 | 408.27 | 17.27948 | | | | D |
| SS-HPV(r), 23y, 3-7y | 1.820 | 0.085 | 86.51 | 75.90727 | 410.07 | 17.28006 | 66.10 | 0.00019 | 348135 | ND |
| CC-HPV(s), 25y, 3-7y | 0.903 | 0.156 | 84.71 | 75.90072 | 448.73 | 17.27902 | | | | D |
| CC-HPV, 23y, 5-10y | 1.151 | 0.126 | 83.79 | 75.90396 | 464.34 | 17.27976 | | | | D |
| CC-HPV, 23y, 5-7y | 1.178 | 0.115 | 85.14 | 75.90477 | 470.37 | 17.27978 | | | | D |
| CC-HPV(s), 23y, 3-7y | 0.889 | 0.145 | 85.93 | 75.90172 | 485.52 | 17.27920 | | | | D |
| CC-HPV, 25y, 3-7y | 1.214 | 0.117 | 85.66 | 75.90498 | 510.20 | 17.27968 | | | | D |
| CC-HPV, 23y, 3-7y | 1.397 | 0.093 | 85.61 | 75.90667 | 592.76 | 17.27999 | | | | D |

Table H.4: Summary statistics, costs, quality-adjusted life-years and ICERs on the cost-efficiency frontier, *5% discount rate for QALYs and costs*, Sweden. Abbreviations: QALE=Quality-adjusted life expectancy; QALY: Quality-adjusted life-year; ICER=Incremental cost-effectiveness ratio; ND=Not dominated; D=Dominated; ED=Extended dominated.

#### H.5 50% lower costs

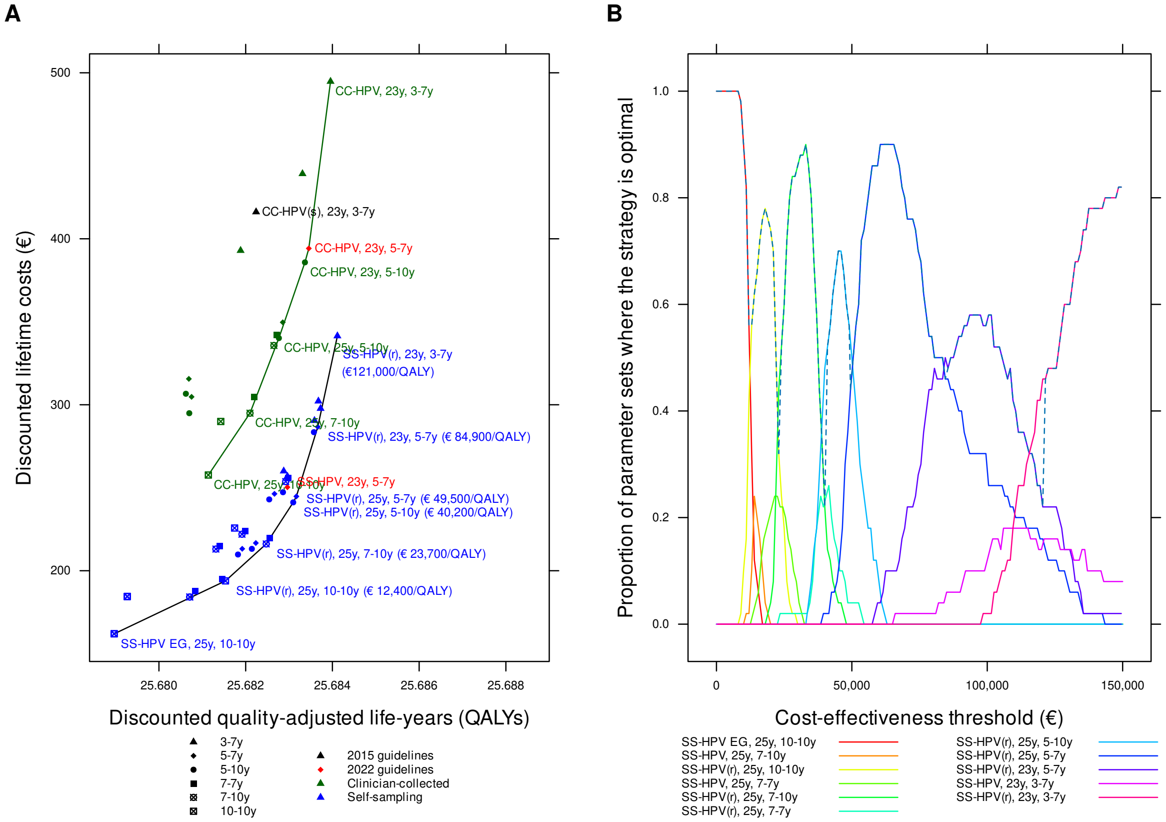

Figure H.5: Cost-effectiveness of cervical cancer screening strategies in unvaccinated cohorts in Sweden: *50% lower costs*. Panel A: cost-efficiency frontier for clinician-collected tests (green) and all testing (black). Panel B: acceptability curves for different cost-effectiveness thresholds (the dashed line is the cost-effectiveness acceptability frontier).

| Strategy | Expected number of colposcopy referrals per woman over lifetime | Mean lifetime risk of cancer (%) | Proportion of cancers detected at local stage (%) | Life-years (undiscounted, from age 9 years) | Total cost per woman (€; discounted) | QALE (discounted) | Incremental cost (€; discounted) | Incremental QALEs (discounted) | ICER (Cost (€) per QALY gained) | Status |
|---|---|---|---|---|---|---|---|---|---|---|
| SS-HPV EG, 25y, 10-10y | 0.625 | 0.278 | 79.46 | 75.88523 | 161.98 | 25.67897 | | | | ND |
| SS-HPV, 25y, 10-10y | 0.626 | 0.277 | 79.46 | 75.88525 | 162.06 | 25.67897 | | | | ED |
| SS-HPV EG, 25y, 7-10y | 0.736 | 0.224 | 81.14 | 75.89366 | 184.20 | 25.68071 | | | | ED |
| SS-HPV, 25y, 7-10y | 0.736 | 0.224 | 81.14 | 75.89366 | 184.20 | 25.68071 | | | | D |
| SS-HPV EG, 23y, 10-10y | 0.710 | 0.274 | 79.08 | 75.88551 | 184.40 | 25.67927 | | | | D |
| SS-HPV, 23y, 10-10y | 0.711 | 0.274 | 79.07 | 75.88552 | 184.55 | 25.67927 | | | | D |
| SS-HPV EG, 25y, 7-7y | 0.763 | 0.205 | 82.27 | 75.89489 | 187.69 | 25.68084 | | | | ED |
| SS-HPV(r), 25y, 10-10y | 0.976 | 0.190 | 82.32 | 75.89666 | 193.89 | 25.68153 | 31.91 | 0.00257 | 12425 | ND |
| SS-HPV, 25y, 7-7y | 0.751 | 0.185 | 83.00 | 75.89744 | 194.94 | 25.68146 | | | | D |
| SS-HPV EG, 25y, 5-10y | 0.864 | 0.173 | 83.11 | 75.89866 | 209.79 | 25.68182 | | | | ED |
| SS-HPV EG, 23y, 7-10y | 0.856 | 0.197 | 82.41 | 75.89549 | 213.06 | 25.68131 | | | | D |
| SS-HPV, 25y, 5-10y | 0.824 | 0.163 | 83.28 | 75.89991 | 213.19 | 25.68214 | | | | ED |
| SS-HPV EG, 25y, 5-7y | 0.885 | 0.162 | 83.72 | 75.89952 | 213.23 | 25.68192 | | | | D |
| SS-HPV EG, 23y, 7-7y | 0.867 | 0.192 | 82.74 | 75.89621 | 214.98 | 25.68140 | | | | D |
| SS-HPV(r), 25y, 7-10y | 1.115 | 0.160 | 83.65 | 75.90091 | 216.16 | 25.68247 | 22.27 | 0.00094 | 23701 | ND |
| SS-HPV, 25y, 5-7y | 0.845 | 0.152 | 84.07 | 75.90071 | 216.70 | 25.68223 | | | | D |
| SS-HPV(r), 25y, 7-7y | 1.152 | 0.146 | 84.90 | 75.90172 | 219.62 | 25.68256 | | | | ED |
| SS-HPV, 23y, 7-10y | 0.849 | 0.174 | 82.75 | 75.89785 | 222.00 | 25.68191 | | | | D |
| SS-HPV, 23y, 7-7y | 0.858 | 0.170 | 82.89 | 75.89843 | 223.84 | 25.68198 | | | | D |
| SS-HPV(r), 23y, 10-10y | 1.123 | 0.183 | 81.41 | 75.89651 | 225.74 | 25.68174 | | | | D |
| SS-HPV(r), 25y, 5-10y | 1.277 | 0.127 | 84.87 | 75.90375 | 241.19 | 25.68310 | 25.03 | 0.00062 | 40209 | ND |
| SS-HPV EG, 23y, 5-10y | 0.992 | 0.162 | 82.52 | 75.90044 | 242.97 | 25.68254 | | | | D |
| SS-HPV(r), 25y, 5-7y | 1.313 | 0.120 | 85.54 | 75.90440 | 244.80 | 25.68317 | 3.61 | 0.00007 | 49481 | ND |
| SS-HPV EG, 23y, 5-7y | 1.017 | 0.144 | 84.10 | 75.90159 | 246.34 | 25.68266 | | | | D |
| SS-HPV, 23y, 5-10y | 0.950 | 0.149 | 82.79 | 75.90183 | 247.30 | 25.68286 | | | | D |
| SS-HPV, 23y, 5-7y | 0.969 | 0.135 | 84.39 | 75.90278 | 250.32 | 25.68296 | | | | D |
| SS-HPV(r), 23y, 7-10y | 1.304 | 0.136 | 84.61 | 75.90204 | 253.71 | 25.68292 | | | | D |
| SS-HPV(r), 23y, 7-7y | 1.321 | 0.133 | 84.76 | 75.90252 | 255.82 | 25.68298 | | | | D |
| SS-HPV, 25y, 3-7y | 0.989 | 0.131 | 84.59 | 75.90384 | 256.83 | 25.68296 | | | | D |
| CC-HPV, 25y, 10-10y | 0.786 | 0.202 | 81.66 | 75.89505 | 257.67 | 25.68114 | | | | D |
| SS-HPV EG, 25y, 3-7y | 1.086 | 0.134 | 84.55 | 75.90349 | 260.05 | 25.68288 | | | | D |
| SS-HPV(r), 23y, 5-10y | 1.476 | 0.118 | 84.34 | 75.90464 | 283.49 | 25.68357 | | | | ED |
| SS-HPV(r), 23y, 5-7y | 1.510 | 0.106 | 86.31 | 75.90550 | 286.68 | 25.68366 | 41.88 | 0.00049 | 84907 | ND |
| CC-HPV, 23y, 10-10y | 0.895 | 0.196 | 80.93 | 75.89520 | 289.93 | 25.68143 | | | | D |
| SS-HPV(r), 25y, 3-7y | 1.566 | 0.106 | 86.02 | 75.90596 | 290.46 | 25.68359 | | | | D |
| CC-HPV(s), 25y, 5-10y | 0.739 | 0.198 | 83.74 | 75.89615 | 294.91 | 25.68070 | | | | D |
| CC-HPV, 25y, 7-10y | 0.887 | 0.173 | 83.30 | 75.89943 | 294.93 | 25.68210 | | | | D |
| SS-HPV, 23y, 3-7y | 1.147 | 0.103 | 85.19 | 75.90598 | 297.74 | 25.68373 | | | | ED |
| SS-HPV EG, 23y, 3-7y | 1.256 | 0.105 | 85.10 | 75.90575 | 302.17 | 25.68368 | | | | D |

Continued on next page

| Strategy | Expected number of colposcopy referrals per woman over lifetime | Mean lifetime risk of cancer (%) | Proportion of cancers detected at local stage (%) | Life-years (undiscounted, from age 9 years) | Total cost per woman (€; discounted) | QALE (discounted) | Incremental cost (€; discounted) | Incremental QALEs (discounted) | ICER (Cost (€) per QALY gained) | Status |
|---|---|---|---|---|---|---|---|---|---|---|
| CC-HPV, 25y, 7-7y | 0.920 | 0.158 | 84.44 | 75.90032 | 304.59 | 25.68219 | | | | D |
| CC-HPV(s), 25y, 5-7y | 0.766 | 0.191 | 83.98 | 75.89662 | 304.77 | 25.68075 | | | | D |
| CC-HPV(s), 23y, 5-10y | 0.703 | 0.210 | 83.02 | 75.89514 | 306.64 | 25.68062 | | | | D |
| CC-HPV(s), 23y, 5-7y | 0.729 | 0.200 | 83.71 | 75.89581 | 315.60 | 25.68069 | | | | D |
| CC-HPV, 23y, 7-10y | 1.028 | 0.148 | 83.42 | 75.90099 | 335.70 | 25.68265 | | | | D |
| CC-HPV, 25y, 5-10y | 1.005 | 0.142 | 84.45 | 75.90248 | 340.16 | 25.68277 | | | | D |
| SS-HPV(r), 23y, 3-7y | 1.820 | 0.085 | 86.51 | 75.90727 | 341.44 | 25.68411 | 54.76 | 0.00045 | 121047 | ND |
| CC-HPV, 23y, 7-7y | 1.046 | 0.144 | 83.94 | 75.90159 | 342.03 | 25.68272 | | | | D |
| CC-HPV, 25y, 5-7y | 1.035 | 0.133 | 85.29 | 75.90330 | 349.67 | 25.68286 | | | | D |
| CC-HPV, 23y, 5-10y | 1.151 | 0.126 | 83.79 | 75.90396 | 385.79 | 25.68337 | | | | D |
| CC-HPV(s), 25y, 3-7y | 0.903 | 0.156 | 84.71 | 75.90072 | 392.91 | 25.68188 | | | | D |
| CC-HPV, 23y, 5-7y | 1.179 | 0.115 | 85.15 | 75.90480 | 394.19 | 25.68346 | | | | D |
| CC-HPV(s), 23y, 3-7y | 0.889 | 0.145 | 85.93 | 75.90172 | 416.16 | 25.68224 | | | | D |
| CC-HPV, 25y, 3-7y | 1.214 | 0.117 | 85.66 | 75.90498 | 439.16 | 25.68331 | | | | D |
| CC-HPV, 23y, 3-7y | 1.397 | 0.093 | 85.63 | 75.90666 | 494.78 | 25.68396 | | | | D |

Table H.5: Summary statistics, costs, quality-adjusted life-years and ICERs on the cost-efficiency frontier, *50% lower costs*, Sweden. Abbreviations: QALE=Quality-adjusted life expectancy; QALY: Quality-adjusted life-year; ICER=Incremental cost-effectiveness ratio; ND=Not dominated; D=Dominated; ED=Extended dominated.

#### H.6 50% higher costs

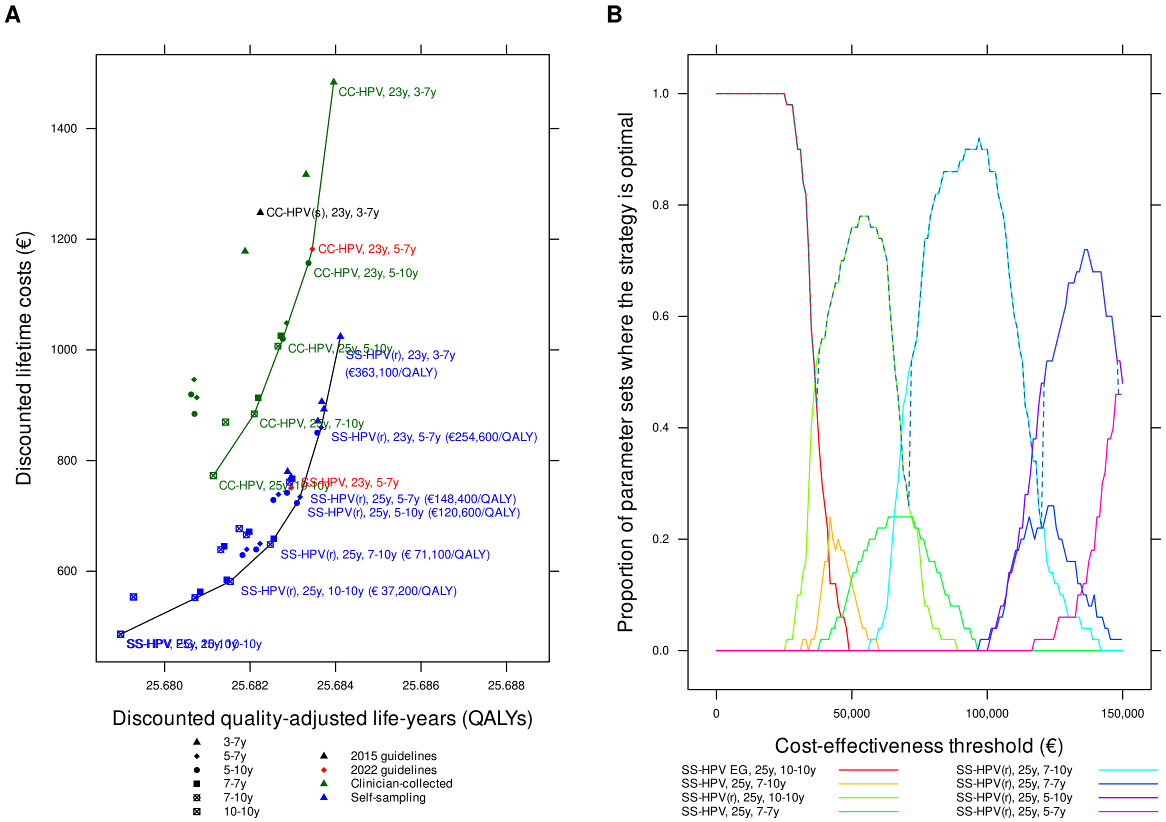

Figure H.6: Cost-effectiveness of cervical cancer screening strategies in unvaccinated cohorts in Sweden: *50% higher costs*. Panel A: cost-efficiency frontier for clinician-collected tests (green) and all testing (black). Panel B: acceptability curves for different cost-effectiveness thresholds (the dashed line is the cost-effectiveness acceptability frontier).

| Strategy | Expected number of colposcopy referrals per woman over lifetime | Mean lifetime risk of cancer (%) | Proportion of cancers detected at local stage (%) | Life-years (undiscounted, from age 9 years) | Total cost per woman (€; discounted) | QALE (discounted) | Incremental cost (€; discounted) | Incremental QALEs (discounted) | ICER (Cost (€) per QALY gained) | Status |
|---|---|---|---|---|---|---|---|---|---|---|
| SS-HPV EG, 25y, 10-10y | 0.626 | 0.277 | 79.46 | 75.88525 | 486.07 | 25.67897 | | | | ND |
| SS-HPV, 25y, 10-10y | 0.626 | 0.277 | 79.46 | 75.88525 | 486.07 | 25.67897 | | | | D |
| SS-HPV EG, 25y, 7-10y | 0.736 | 0.224 | 81.14 | 75.89366 | 552.46 | 25.68071 | | | | ED |
| SS-HPV, 25y, 7-10y | 0.736 | 0.224 | 81.14 | 75.89366 | 552.46 | 25.68071 | | | | D |
| SS-HPV EG, 23y, 10-10y | 0.711 | 0.274 | 79.07 | 75.88552 | 553.52 | 25.67927 | | | | D |
| SS-HPV, 23y, 10-10y | 0.711 | 0.274 | 79.07 | 75.88552 | 553.52 | 25.67927 | | | | D |
| SS-HPV EG, 25y, 7-7y | 0.763 | 0.205 | 82.27 | 75.89489 | 562.94 | 25.68084 | | | | ED |
| SS-HPV(r), 25y, 10-10y | 0.976 | 0.190 | 82.32 | 75.89666 | 581.52 | 25.68153 | 95.46 | 0.00256 | 37218 | ND |
| SS-HPV, 25y, 7-7y | 0.751 | 0.185 | 82.99 | 75.89743 | 584.58 | 25.68146 | | | | D |
| SS-HPV EG, 25y, 5-10y | 0.864 | 0.173 | 83.11 | 75.89866 | 629.20 | 25.68182 | | | | ED |
| SS-HPV EG, 23y, 7-10y | 0.856 | 0.197 | 82.41 | 75.89549 | 639.02 | 25.68131 | | | | D |
| SS-HPV, 25y, 5-10y | 0.824 | 0.163 | 83.28 | 75.89991 | 639.41 | 25.68214 | | | | ED |
| SS-HPV EG, 25y, 5-7y | 0.885 | 0.162 | 83.73 | 75.89952 | 639.58 | 25.68192 | | | | D |
| SS-HPV EG, 23y, 7-7y | 0.867 | 0.192 | 82.74 | 75.89621 | 644.77 | 25.68140 | | | | D |
| SS-HPV(r), 25y, 7-10y | 1.115 | 0.160 | 83.65 | 75.90091 | 648.32 | 25.68247 | 66.79 | 0.00094 | 71084 | ND |
| SS-HPV, 25y, 5-7y | 0.845 | 0.152 | 84.08 | 75.90071 | 649.79 | 25.68223 | | | | D |
| SS-HPV(r), 25y, 7-7y | 1.153 | 0.146 | 84.91 | 75.90172 | 658.82 | 25.68256 | | | | ED |
| SS-HPV, 23y, 7-10y | 0.849 | 0.174 | 82.75 | 75.89785 | 665.82 | 25.68191 | | | | D |
| SS-HPV, 23y, 7-7y | 0.858 | 0.170 | 82.89 | 75.89843 | 671.43 | 25.68198 | | | | D |
| SS-HPV(r), 23y, 10-10y | 1.123 | 0.183 | 81.41 | 75.89651 | 677.06 | 25.68174 | | | | D |
| SS-HPV(r), 25y, 5-10y | 1.277 | 0.127 | 84.87 | 75.90375 | 723.39 | 25.68310 | 75.07 | 0.00062 | 120599 | ND |
| SS-HPV EG, 23y, 5-10y | 0.992 | 0.162 | 82.52 | 75.90044 | 728.74 | 25.68254 | | | | D |
| SS-HPV(r), 25y, 5-7y | 1.313 | 0.120 | 85.54 | 75.90440 | 734.22 | 25.68317 | 10.83 | 0.00007 | 148419 | ND |
| SS-HPV EG, 23y, 5-7y | 1.017 | 0.144 | 84.10 | 75.90159 | 738.83 | 25.68266 | | | | D |
| SS-HPV, 23y, 5-10y | 0.950 | 0.149 | 82.81 | 75.90184 | 741.76 | 25.68286 | | | | D |
| SS-HPV, 23y, 5-7y | 0.970 | 0.135 | 84.39 | 75.90278 | 750.84 | 25.68296 | | | | D |
| SS-HPV(r), 23y, 7-10y | 1.304 | 0.136 | 84.61 | 75.90204 | 760.94 | 25.68292 | | | | D |
| SS-HPV(r), 23y, 7-7y | 1.321 | 0.133 | 84.76 | 75.90252 | 767.29 | 25.68298 | | | | D |
| SS-HPV, 25y, 3-7y | 0.989 | 0.131 | 84.60 | 75.90384 | 770.04 | 25.68296 | | | | D |
| CC-HPV, 25y, 10-10y | 0.786 | 0.202 | 81.66 | 75.89505 | 772.72 | 25.68114 | | | | D |
| SS-HPV EG, 25y, 3-7y | 1.086 | 0.134 | 84.55 | 75.90349 | 779.94 | 25.68288 | | | | D |
| SS-HPV(r), 23y, 5-10y | 1.476 | 0.118 | 84.34 | 75.90464 | 850.25 | 25.68357 | | | | ED |
| SS-HPV(r), 23y, 5-7y | 1.510 | 0.106 | 86.31 | 75.90550 | 859.82 | 25.68366 | 125.61 | 0.00049 | 254647 | ND |
| CC-HPV, 23y, 10-10y | 0.895 | 0.196 | 80.93 | 75.89520 | 869.45 | 25.68143 | | | | D |
| SS-HPV(r), 25y, 3-7y | 1.566 | 0.106 | 86.02 | 75.90596 | 871.16 | 25.68359 | | | | D |
| CC-HPV(s), 25y, 5-10y | 0.739 | 0.198 | 83.74 | 75.89615 | 884.35 | 25.68070 | | | | D |
| CC-HPV, 25y, 7-10y | 0.887 | 0.173 | 83.30 | 75.89943 | 884.44 | 25.68210 | | | | D |
| SS-HPV, 23y, 3-7y | 1.147 | 0.103 | 85.18 | 75.90598 | 892.90 | 25.68373 | | | | ED |
| SS-HPV EG, 23y, 3-7y | 1.256 | 0.105 | 85.10 | 75.90575 | 906.28 | 25.68368 | | | | D |

Continued on next page

| Strategy | Expected number of colposcopy referrals per woman over life-time | Mean life-time risk of cancer (%) | Proportion of cancers detected at local stage (%) | Life-years (undis-counted, from age 9 years) | Total cost per woman (€; dis-counted) | QALE (dis-counted) | Incremental cost (€; dis-counted) | Incremental QALEs (dis-counted) | ICER (Cost (€) per QALY gained) | Status |
|---|---|---|---|---|---|---|---|---|---|---|
| CC-HPV, 25y, 7-7y | 0.920 | 0.158 | 84.44 | 75.90032 | 913.39 | 25.68219 | | | | D |
| CC-HPV(s), 25y, 5-7y | 0.766 | 0.191 | 83.98 | 75.89662 | 913.91 | 25.68075 | | | | D |
| CC-HPV(s), 23y, 5-10y | 0.703 | 0.210 | 83.02 | 75.89514 | 919.51 | 25.68062 | | | | D |
| CC-HPV(s), 23y, 5-7y | 0.729 | 0.200 | 83.71 | 75.89581 | 946.38 | 25.68069 | | | | D |
| CC-HPV, 23y, 7-10y | 1.028 | 0.148 | 83.42 | 75.90099 | 1006.70 | 25.68265 | | | | D |
| CC-HPV, 25y, 5-10y | 1.005 | 0.142 | 84.45 | 75.90248 | 1020.04 | 25.68277 | | | | D |
| SS-HPV(r), 23y, 3-7y | 1.820 | 0.085 | 86.51 | 75.90727 | 1024.07 | 25.68411 | 164.25 | 0.00045 | 363061 | ND |
| CC-HPV, 23y, 7-7y | 1.046 | 0.144 | 83.94 | 75.90159 | 1025.68 | 25.68272 | | | | D |
| CC-HPV, 25y, 5-7y | 1.035 | 0.133 | 85.29 | 75.90330 | 1048.57 | 25.68286 | | | | D |
| CC-HPV, 23y, 5-10y | 1.151 | 0.126 | 83.78 | 75.90396 | 1156.78 | 25.68337 | | | | D |
| CC-HPV(s), 25y, 3-7y | 0.903 | 0.156 | 84.71 | 75.90072 | 1178.19 | 25.68188 | | | | D |
| CC-HPV, 23y, 5-7y | 1.179 | 0.115 | 85.15 | 75.90480 | 1182.07 | 25.68346 | | | | D |
| CC-HPV(s), 23y, 3-7y | 0.889 | 0.145 | 85.93 | 75.90172 | 1247.89 | 25.68224 | | | | D |
| CC-HPV, 25y, 3-7y | 1.214 | 0.117 | 85.66 | 75.90498 | 1316.91 | 25.68331 | | | | D |
| CC-HPV, 23y, 3-7y | 1.397 | 0.093 | 85.63 | 75.90666 | 1483.68 | 25.68396 | | | | D |

Table H.6: Summary statistics, costs, quality-adjusted life-years and ICERs on the cost-efficiency frontier, *50% higher costs*, Sweden. Abbreviations: QALE=Quality-adjusted life expectancy; QALY: Quality-adjusted life-year; ICER=Incremental cost-effectiveness ratio; ND=Not dominated; D=Dominated; ED=Extended dominated.

#### H.7 Life expectancy

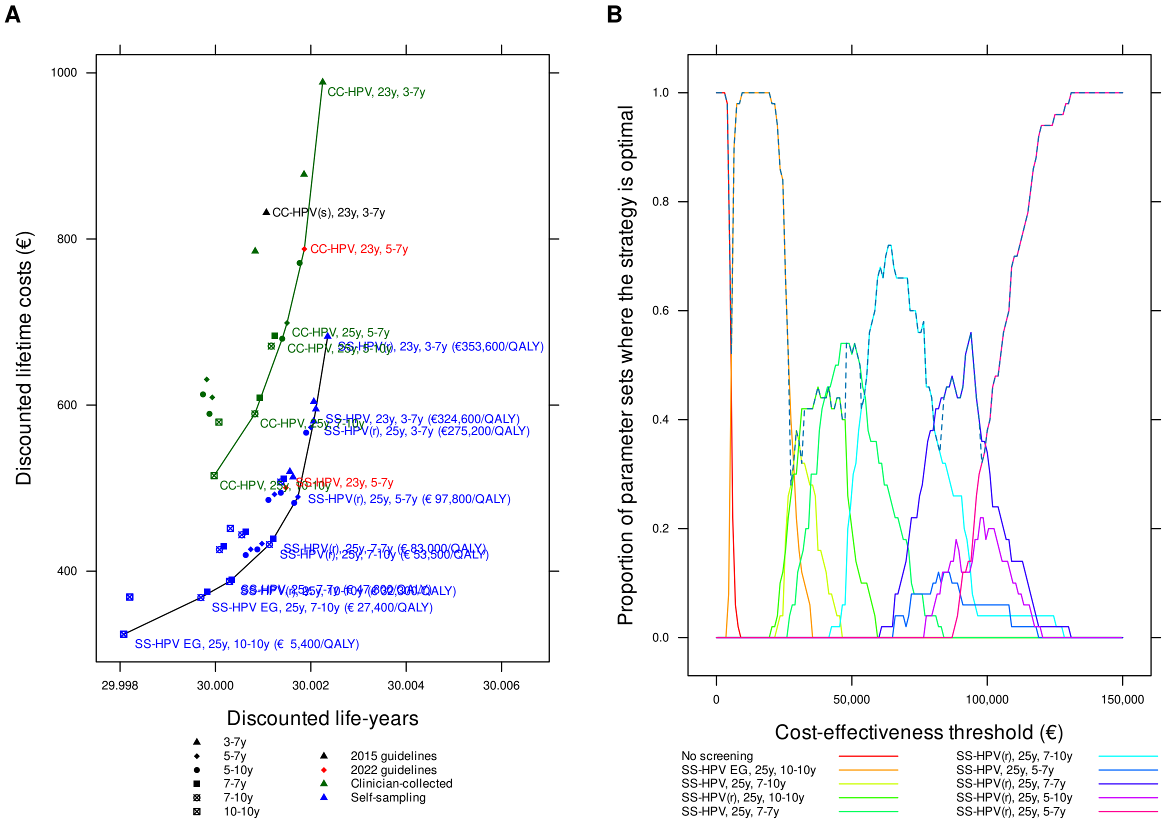

Figure H.7: Cost-effectiveness of cervical cancer screening strategies in unvaccinated cohorts in Sweden: *effectiveness measured using life expectancy*. Panel A: cost-efficiency frontier for clinician-collected tests (green) and all testing (black). Panel B: acceptability curves for different cost-effectiveness thresholds (the dashed line is the cost-effectiveness acceptability frontier).

| Strategy | Expected number of colposcopy referrals per woman over lifetime | Mean lifetime risk of cancer (%) | Proportion of cancers detected at local stage (%) | Life-years (undiscounted, from age 9 years) | Total cost per woman (€; discounted) | QALE (discounted) | Incremental cost (€; discounted) | Incremental QALEs (discounted) | ICER (Cost (€) per QALY gained) | Status |
|---|---|---|---|---|---|---|---|---|---|---|
| No screening | 0.000 | 1.744 | 67.10 | 75.69084 | 145.51 | 25.64434 | | | | ND |
| SS-HPV EG, 25y, 10-10y | 0.626 | 0.277 | 79.46 | 75.88525 | 324.01 | 25.67897 | 178.49 | 0.03463 | 5154 | ND |
| SS-HPV, 25y, 10-10y | 0.626 | 0.278 | 79.44 | 75.88523 | 324.03 | 25.67896 | | | | D |
| SS-HPV EG, 25y, 7-10y | 0.736 | 0.224 | 81.14 | 75.89366 | 368.26 | 25.68071 | | | | ED |
| SS-HPV, 25y, 7-10y | 0.736 | 0.224 | 81.14 | 75.89366 | 368.26 | 25.68071 | | | | D |
| SS-HPV, 23y, 10-10y | 0.710 | 0.274 | 79.08 | 75.88550 | 368.78 | 25.67927 | | | | D |
| SS-HPV EG, 23y, 10-10y | 0.711 | 0.274 | 79.07 | 75.88552 | 368.97 | 25.67927 | | | | D |
| SS-HPV EG, 25y, 7-7y | 0.763 | 0.205 | 82.27 | 75.89489 | 375.25 | 25.68084 | | | | ED |
| SS-HPV(r), 25y, 10-10y | 0.976 | 0.190 | 82.32 | 75.89666 | 387.64 | 25.68153 | 63.63 | 0.00256 | 24809 | ND |
| SS-HPV, 25y, 7-7y | 0.751 | 0.185 | 82.99 | 75.89743 | 389.67 | 25.68146 | | | | D |
| SS-HPV EG, 25y, 5-10y | 0.864 | 0.173 | 83.11 | 75.89866 | 419.41 | 25.68182 | | | | ED |
| SS-HPV EG, 23y, 7-10y | 0.856 | 0.197 | 82.41 | 75.89549 | 425.96 | 25.68131 | | | | D |
| SS-HPV, 25y, 5-10y | 0.824 | 0.163 | 83.28 | 75.89991 | 426.22 | 25.68214 | | | | ED |
| SS-HPV EG, 25y, 5-7y | 0.885 | 0.162 | 83.73 | 75.89952 | 426.34 | 25.68192 | | | | D |
| SS-HPV EG, 23y, 7-7y | 0.867 | 0.192 | 82.75 | 75.89619 | 429.80 | 25.68140 | | | | D |
| SS-HPV(r), 25y, 7-10y | 1.115 | 0.160 | 83.64 | 75.90093 | 432.14 | 25.68248 | 44.50 | 0.00094 | 47104 | ND |
| SS-HPV, 25y, 5-7y | 0.845 | 0.152 | 84.08 | 75.90071 | 433.14 | 25.68223 | | | | D |
| SS-HPV(r), 25y, 7-7y | 1.153 | 0.146 | 84.91 | 75.90172 | 439.16 | 25.68256 | | | | ED |
| SS-HPV, 23y, 7-10y | 0.849 | 0.174 | 82.75 | 75.89785 | 443.82 | 25.68191 | | | | D |
| SS-HPV, 23y, 7-7y | 0.858 | 0.170 | 82.89 | 75.89845 | 447.70 | 25.68199 | | | | D |
| SS-HPV(r), 23y, 10-10y | 1.124 | 0.183 | 81.41 | 75.89652 | 451.45 | 25.68175 | | | | D |
| SS-HPV(r), 25y, 5-10y | 1.277 | 0.127 | 84.87 | 75.90375 | 482.20 | 25.68310 | 50.07 | 0.00062 | 81087 | ND |
| SS-HPV EG, 23y, 5-10y | 0.992 | 0.162 | 82.52 | 75.90044 | 485.76 | 25.68254 | | | | D |
| SS-HPV(r), 25y, 5-7y | 1.313 | 0.120 | 85.54 | 75.90440 | 489.42 | 25.68317 | 7.22 | 0.00007 | 98938 | ND |
| SS-HPV EG, 23y, 5-7y | 1.017 | 0.144 | 84.10 | 75.90159 | 492.49 | 25.68266 | | | | D |
| SS-HPV, 23y, 5-10y | 0.950 | 0.149 | 82.81 | 75.90184 | 494.44 | 25.68286 | | | | D |
| SS-HPV, 23y, 5-7y | 0.970 | 0.135 | 84.39 | 75.90278 | 500.49 | 25.68296 | | | | D |
| SS-HPV(r), 23y, 7-10y | 1.304 | 0.135 | 84.62 | 75.90205 | 507.39 | 25.68293 | | | | D |
| SS-HPV(r), 23y, 7-7y | 1.321 | 0.133 | 84.76 | 75.90252 | 511.46 | 25.68298 | | | | D |
| SS-HPV, 25y, 3-7y | 0.989 | 0.131 | 84.60 | 75.90384 | 513.30 | 25.68296 | | | | D |
| CC-HPV, 25y, 10-10y | 0.786 | 0.202 | 81.66 | 75.89505 | 515.05 | 25.68114 | | | | D |
| SS-HPV EG, 25y, 3-7y | 1.086 | 0.134 | 84.55 | 75.90349 | 519.90 | 25.68288 | | | | D |
| SS-HPV(r), 23y, 5-10y | 1.476 | 0.118 | 84.34 | 75.90464 | 566.76 | 25.68357 | | | | ED |
| SS-HPV(r), 23y, 5-7y | 1.510 | 0.106 | 86.31 | 75.90550 | 573.15 | 25.68366 | 83.73 | 0.00049 | 169740 | ND |
| CC-HPV, 23y, 10-10y | 0.895 | 0.196 | 80.93 | 75.89520 | 579.52 | 25.68143 | | | | D |
| SS-HPV(r), 25y, 3-7y | 1.566 | 0.106 | 86.02 | 75.90596 | 580.71 | 25.68359 | | | | D |
| CC-HPV(s), 25y, 5-10y | 0.739 | 0.198 | 83.74 | 75.89615 | 589.44 | 25.68070 | | | | D |
| CC-HPV, 25y, 7-10y | 0.887 | 0.173 | 83.30 | 75.89943 | 589.51 | 25.68210 | | | | D |
| SS-HPV, 23y, 3-7y | 1.147 | 0.103 | 85.18 | 75.90598 | 595.19 | 25.68373 | | | | ED |

Continued on next page

| Strategy | Expected number of colposcopy referrals per woman over lifetime | Mean lifetime risk of cancer (%) | Proportion of cancers detected at local stage (%) | Life-years (undiscounted, from age 9 years) | Total cost per woman (€; discounted) | QALE (discounted) | Incremental cost (€; discounted) | Incremental QALEs (discounted) | ICER (Cost (€) per QALY gained) | Status |
|---|---|---|---|---|---|---|---|---|---|---|
| SS-HPV EG, 23y, 3-7y | 1.256 | 0.105 | 85.10 | 75.90575 | 604.11 | 25.68368 | | | | D |
| CC-HPV, 25y, 7-7y | 0.920 | 0.158 | 84.44 | 75.90032 | 608.81 | 25.68219 | | | | D |
| CC-HPV(s), 25y, 5-7y | 0.767 | 0.191 | 83.98 | 75.89664 | 609.29 | 25.68076 | | | | D |
| CC-HPV(s), 23y, 5-10y | 0.703 | 0.210 | 83.02 | 75.89514 | 612.87 | 25.68062 | | | | D |
| CC-HPV(s), 23y, 5-7y | 0.730 | 0.200 | 83.71 | 75.89583 | 630.89 | 25.68069 | | | | D |
| CC-HPV, 23y, 7-10y | 1.028 | 0.148 | 83.42 | 75.90099 | 671.00 | 25.68265 | | | | D |
| CC-HPV, 25y, 5-10y | 1.005 | 0.142 | 84.44 | 75.90247 | 679.91 | 25.68276 | | | | D |
| SS-HPV(r), 23y, 3-7y | 1.820 | 0.085 | 86.51 | 75.90727 | 682.63 | 25.68411 | 109.49 | 0.00045 | 242014 | ND |
| CC-HPV, 23y, 7-7y | 1.046 | 0.144 | 83.94 | 75.90159 | 683.65 | 25.68272 | | | | D |
| CC-HPV, 25y, 5-7y | 1.035 | 0.133 | 85.29 | 75.90330 | 698.90 | 25.68286 | | | | D |
| CC-HPV, 23y, 5-10y | 1.151 | 0.126 | 83.79 | 75.90396 | 771.10 | 25.68337 | | | | D |
| CC-HPV(s), 25y, 3-7y | 0.903 | 0.156 | 84.71 | 75.90072 | 785.28 | 25.68188 | | | | D |
| CC-HPV, 23y, 5-7y | 1.179 | 0.115 | 85.14 | 75.90480 | 787.93 | 25.68346 | | | | D |
| CC-HPV(s), 23y, 3-7y | 0.889 | 0.145 | 85.93 | 75.90172 | 831.74 | 25.68224 | | | | D |
| CC-HPV, 25y, 3-7y | 1.214 | 0.117 | 85.66 | 75.90498 | 877.75 | 25.68331 | | | | D |
| CC-HPV, 23y, 3-7y | 1.396 | 0.093 | 85.63 | 75.90667 | 988.78 | 25.68396 | | | | D |

Table H.7: Summary statistics, costs, life-years and ICERs on the cost-efficiency frontier, *effectiveness measured using life expectancy*, Sweden. Abbreviations: ICER=Incremental cost-effectiveness ratio; ND=Not dominated; D=Dominated; ED=Extended dominated.

#### H.8 Assumed lower accuracy for self-sampling

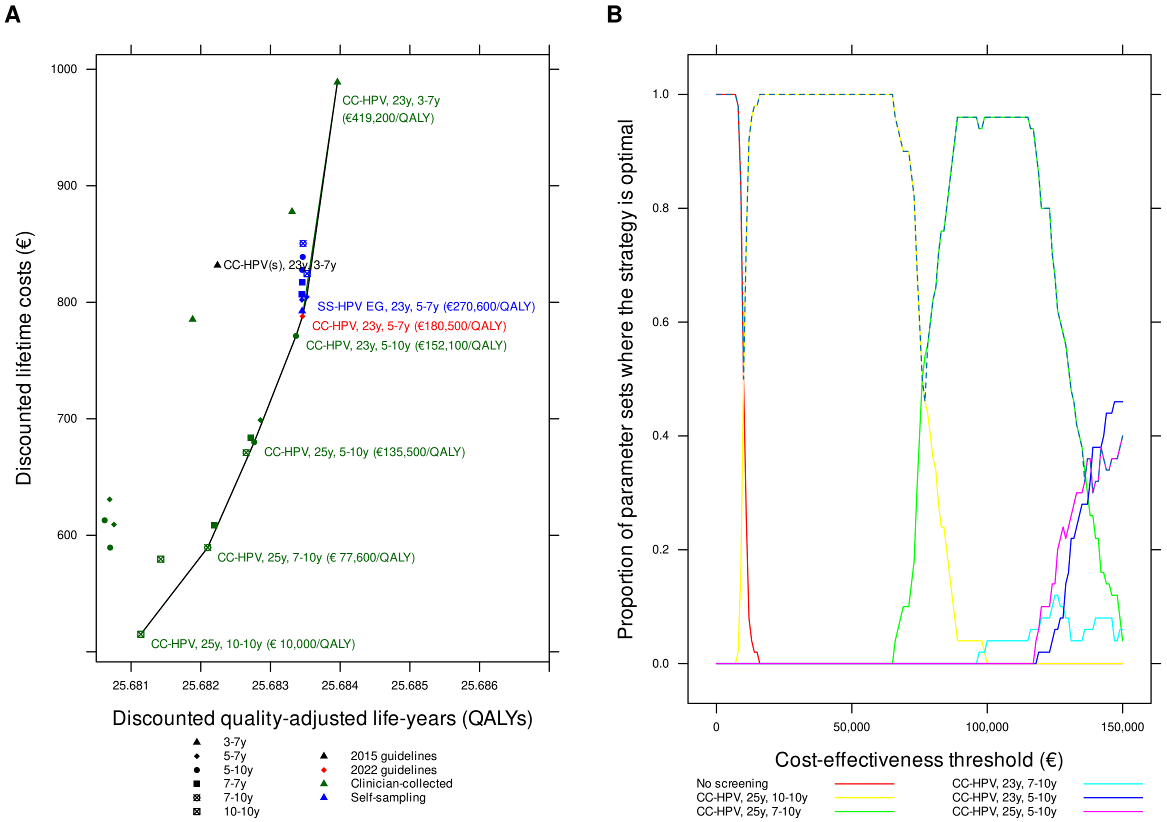

Figure H.8: Cost-effectiveness of cervical cancer screening strategies in unvaccinated cohorts in Sweden: *assumed lower accuracy for self-sampling*. Panel A: cost-efficiency frontier for clinician-collected tests (green) and all testing (black). Panel B: acceptability curves for different cost-effectiveness thresholds (the dashed line is the cost-effectiveness acceptability frontier).

| Strategy | Expected number of colposcopy referrals per woman over lifetime | Mean lifetime risk of cancer (%) | Proportion of cancers detected at local stage (%) | Life-years (undiscounted, from age 9 years) | Total cost per woman (€; discounted) | QALE (discounted) | Incremental cost (€; discounted) | Incremental QALEs (discounted) | ICER (Cost (€) per QALY gained) | Status |
|---|---|---|---|---|---|---|---|---|---|---|
| No screening | 0.000 | 1.744 | 67.10 | 75.69084 | 145.51 | 25.64434 | | | | ND |
| CC-HPV, 25y, 10-10y | 0.786 | 0.202 | 81.66 | 75.89505 | 515.05 | 25.68114 | 369.53 | 0.03680 | 10040 | ND |
| CC-HPV, 23y, 10-10y | 0.895 | 0.196 | 80.93 | 75.89520 | 579.52 | 25.68143 | | | | ED |
| CC-HPV(s), 25y, 5-10y | 0.739 | 0.198 | 83.74 | 75.89615 | 589.44 | 25.68070 | | | | D |
| CC-HPV, 25y, 7-10y | 0.887 | 0.173 | 83.30 | 75.89943 | 589.51 | 25.68210 | 74.46 | 0.00096 | 77594 | ND |
| CC-HPV, 25y, 7-7y | 0.920 | 0.158 | 84.44 | 75.90032 | 608.81 | 25.68219 | | | | ED |
| CC-HPV(s), 25y, 5-7y | 0.767 | 0.191 | 83.98 | 75.89662 | 609.22 | 25.68075 | | | | D |
| CC-HPV(s), 23y, 5-10y | 0.703 | 0.210 | 83.02 | 75.89514 | 612.87 | 25.68062 | | | | D |
| CC-HPV(s), 23y, 5-7y | 0.729 | 0.200 | 83.71 | 75.89582 | 630.82 | 25.68069 | | | | D |
| CC-HPV, 23y, 7-10y | 1.028 | 0.148 | 83.42 | 75.90099 | 671.00 | 25.68265 | | | | ED |
| CC-HPV, 25y, 5-10y | 1.005 | 0.142 | 84.44 | 75.90249 | 679.93 | 25.68277 | 90.43 | 0.00067 | 135489 | ND |
| CC-HPV, 23y, 7-7y | 1.046 | 0.144 | 83.94 | 75.90159 | 683.65 | 25.68272 | | | | D |
| CC-HPV, 25y, 5-7y | 1.035 | 0.133 | 85.29 | 75.90330 | 698.90 | 25.68286 | | | | ED |
| CC-HPV, 23y, 5-10y | 1.151 | 0.126 | 83.79 | 75.90396 | 771.10 | 25.68337 | 91.16 | 0.00060 | 152066 | ND |
| CC-HPV(s), 25y, 3-7y | 0.903 | 0.156 | 84.71 | 75.90072 | 785.28 | 25.68188 | | | | D |
| CC-HPV, 23y, 5-7y | 1.179 | 0.115 | 85.14 | 75.90480 | 787.89 | 25.68346 | 16.80 | 0.00009 | 180504 | ND |
| SS-HPV EG, 25y, 3-7y | 1.198 | 0.115 | 85.07 | 75.90480 | 792.48 | 25.68345 | | | | D |
| SS-HPV EG, 25y, 5-7y | 1.238 | 0.115 | 85.02 | 75.90477 | 802.06 | 25.68345 | | | | D |
| SS-HPV EG, 23y, 5-7y | 1.246 | 0.111 | 85.05 | 75.90507 | 804.59 | 25.68352 | 16.70 | 0.00006 | 270627 | ND |
| SS-HPV EG, 23y, 7-7y | 1.260 | 0.115 | 85.04 | 75.90476 | 806.98 | 25.68345 | | | | D |
| SS-HPV EG, 25y, 7-7y | 1.304 | 0.114 | 85.08 | 75.90475 | 817.15 | 25.68345 | | | | D |
| SS-HPV EG, 23y, 10-10y | 1.332 | 0.110 | 85.00 | 75.90508 | 824.61 | 25.68353 | | | | ED |
| SS-HPV EG, 23y, 5-10y | 1.351 | 0.114 | 85.08 | 75.90477 | 827.79 | 25.68345 | | | | D |
| CC-HPV(s), 23y, 3-7y | 0.889 | 0.145 | 85.93 | 75.90172 | 831.74 | 25.68224 | | | | D |
| SS-HPV EG, 25y, 5-10y | 1.401 | 0.114 | 85.08 | 75.90481 | 838.92 | 25.68346 | | | | D |
| SS-HPV EG, 23y, 7-10y | 1.453 | 0.114 | 84.90 | 75.90484 | 850.45 | 25.68347 | | | | D |
| CC-HPV, 25y, 3-7y | 1.214 | 0.117 | 85.66 | 75.90498 | 877.75 | 25.68331 | | | | D |
| CC-HPV, 23y, 3-7y | 1.396 | 0.093 | 85.63 | 75.90666 | 988.87 | 25.68396 | 184.28 | 0.00044 | 419214 | ND |

Table H.8: Summary statistics, costs, quality-adjusted life-years and ICERs on the cost-efficiency frontier, *assumed lower accuracy for self-sampling*, Sweden. Abbreviations: QALE=Quality-adjusted life expectancy; QALY: Quality-adjusted life-year; ICER=Incremental cost-effectiveness ratio; ND=Not dominated; D=Dominated; ED=Extended dominated.
